# Crypt-top and crypt-bottom colonic epithelial cell population microRNA expression demonstrates cell type-specificity and correlation with endoscopic activity in ulcerative colitis

**DOI:** 10.1101/2022.09.25.22280336

**Authors:** Ruta Inciuraite, Rima Ramonaite, Juozas Kupcinskas, Indre Dalgediene, Ugne Kulokiene, Vytautas Kiudelis, Greta Varkalaite, Aurelija Zvirbliene, Laimas Virginijus Jonaitis, Gediminas Kiudelis, Andre Franke, Stefan Schreiber, Simonas Juzenas, Jurgita Skieceviciene

## Abstract

**Background and Aims:** Colonic epithelial barrier loss and dysfunction are one of the early events in ulcerative colitis (UC) and microRNAs (miRNAs) participate in its regulation. However, cell type-specific profile of miRNAs during inflammation in UC is still unknown. Thus, we aimed to perform miRNA profiling on colon tissue and epithelial cell levels in active and quiescent UC.

**Methods:** Small RNA-sequencing in colon tissue, crypt-bottom (CD44^+^), and crypt-top (CD66a^+^) colonic epithelial cell populations from two independent cohorts of UC patients (active and quiescent, n=74), and healthy individuals (n=50) was performed. Data analysis encompassed differential expression, weighted gene co-expression network (WGCNA), correlation, gene-set enrichment analyses (GSEA).

**Results:** In colon tissue of active and quiescent UC, differentially expressed miRNAs were shown to be potentially involved in intestinal barrier integrity regulation. Consecutive analysis of crypt-bottom and crypt-top colonic epithelial cells revealed distinct miRNA expression patterns in response to UC-caused inflammation. GSEA indicated that differentially expressed epithelial miRNAs are commonly involved in inflammation- and intestinal barrier integrity-related processes (such as signalling of interleukin-4 and interleukin-13), while miRNA differences between cell populations might reflect their function, i.e., crypt-bottom cell miRNA target genes involved in regulation of cell differentiation. Finally, pro-inflammatory miRNA co-expression module correlating with endoscopic UC activity was defined not only in both epithelial cell populations, but also in the colon tissue.

**Conclusions:** miRNA expression patterns are colon epithelial cell population- and UC state-specific and correlate with endoscopic UC activity. Irrespective of the UC stage deregulated epithelial miRNAs are potentially involved in regulation of intestinal barrier integrity.

## Introduction

Colonic epithelial cells and their secreted products primarily form and maintain frontline intestinal barrier function which deteriorates early in ulcerative colitis (UC)^1^. Colonic crypt epithelial cells such as colonocytes, Goblet and epithelial stem cells have been shown to play an important role in UC pathogenesis via reduced epithelial mucus secretion and/or increased barrier permeability^2^. Since one of the aims in UC management is to induce and then to maintain remission, disentangling molecular mechanisms regulating epithelial barrier may help to modulate its permeability and retain long-lasting remission in UC.

Recent studies provide evidence for microRNAs (miRNAs) to be involved in regulation of intestinal epithelial integrity and barrier permeability via interference with protein-coding genes responsible for tight and adherent junctions^3^, also in UC^4^. However, most of the studies have been either based on immortalized cell cultures or bulk tissue experiments, thus the exact cellular context of miRNA deregulation during colonic inflammation remains elusive. Therefore, the search for cell type specific deregulation of miRNAs in UC might uncover novel aspects of underlying regulatory pathways and molecular targets for therapeutic development. Here, we report the comprehensive miRNA expression profiling on colon tissue and colonic crypt-bottom (CD44^+^) and crypt-top (CD66a^+^) cell population levels in active and quiescent UC. By employing small RNA-seq we determine UC activity-specific, as well as epithelial cell population-specific, miRNA expression signatures during colonic inflammation. Additionally, we describe miRNA co-expression network and evaluate its relations to clinical characteristic of UC. Finally, we explore putative biological pathways in which the deregulated miRNAs are involved and provide potential miRNA candidates for further development of UC management.

## Materials and Methods

### Ethics and consent

The approval to perform the study was received from Kaunas Regional Biomedical Research Ethics Committee (No. BE-2-31, 22-03-2018). All subjects have signed a written informed consent form to participate in the study.

### Study samples

Study subject recruitment was conducted at the Department of Gastroenterology, Lithuanian University of Health Sciences (Kaunas, Lithuania) during the period of 2011-2014 (Study group I and III) and 2017-2019 (Study group II). For miRNA sequencing, colon biopsies were collected in two independent groups: the first study group contained 23 patients with active UC, 20 patients with quiescent UC and 33 healthy control individuals (HC), while the second study group was composed of 16 patients with active UC, 15 patients with quiescent UC and 17 HC individuals. Additionally, colon biopsies for qPCR-based gene expression analysis were collected in the third independent group which contained 75 patients with active UC, 50 patients with quiescent UC and 75 HC individuals. Demographic and clinical characteristics of subjects of Study groups I and II are presented in **Table 1**, Study group III – in **Supplementary Table S1**. Tissue samples of patients with active UC and quiescent UC were collected from sigmoid colon. Endoscopic activity was determined using the Mayo endoscopic subscore^5^. Quiescent UC was defined as mucosal healing (endoscopic Mayo subscore ≤ 1) with no clinical symptoms. The variables collected for the individuals of both study groups were such as age, sex, body mass index (BMI), smoking status, disease activity, localization, rectal bleeding, stool frequency, endoscopic Mayo subscore, global assessment score, full Mayo score.

**Table 1.**
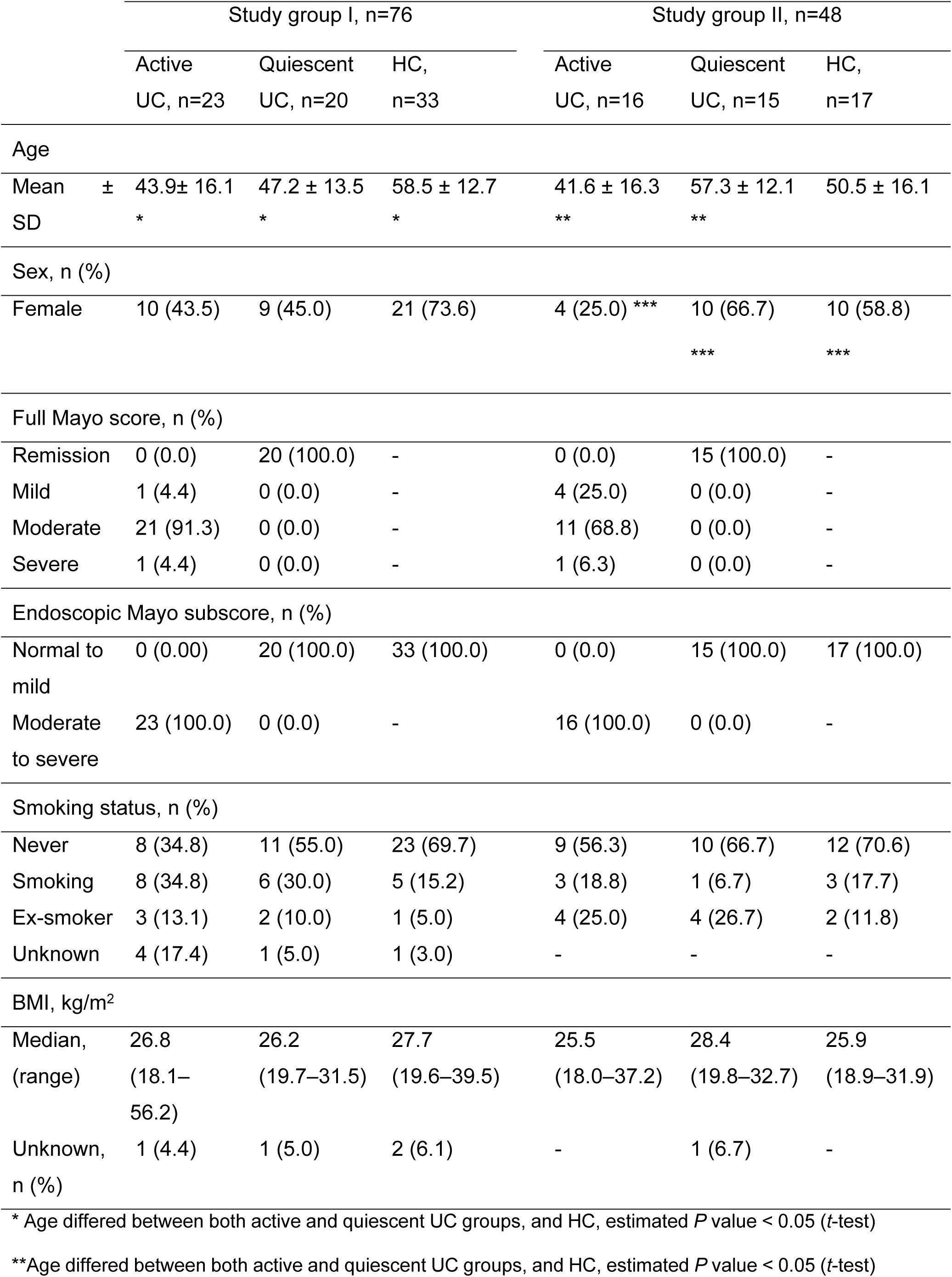
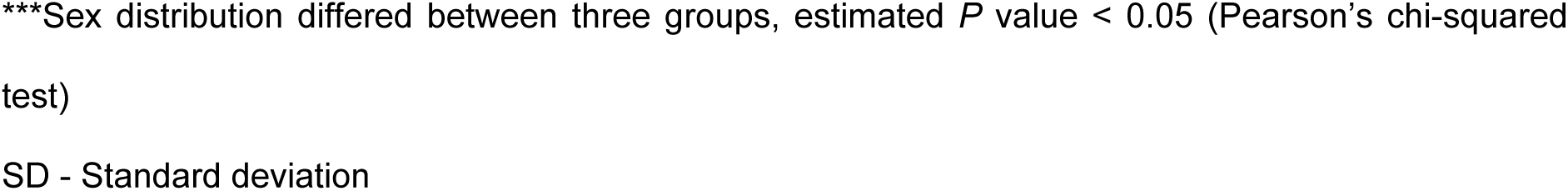
Demographic and clinical characteristics of subjects.

All subjects with active and quiescent UC had a routine colonoscopy performed as a part of their planned examination programme. The diagnosis of UC was based on standard clinical, endoscopic, radiological, and histological criteria^6^. The control individuals included in both study groups consisted of healthy subjects who either underwent colonoscopy due to positive faecal occult blood test or were consulted on functional complaints, but had a normal colonoscopy, uninflamed mucosa during histopathological examination and no previous history of intestinal inflammation. All patients enrolled in the study were of European descent.

### Colon tissue disaggregation

Colon biopsies were mechanically and enzymatically separated into single-cell suspensions. Briefly, four to six biopsies (5–10 mg wet weight each) were washed with PBS solution containing 50 IU/mL penicillin, 50 µg/mL streptomycin, and 0.5 mg/mL gentamicin. Biopsies were minced into small pieces (approx. 1-2 mm^3^) and incubated in 1X trypsin-EDTA solution for 40-45 min at room temperature on an agitator to dissociate single epithelial cells from the lamina propria. After incubation, tissue fragments were gently transferred into PBS and shaken. The isolated cell suspension was filtered and suspended in DMEM:Ham’s F-12 medium (1:1) with 15 mM HEPES (Gibco, USA) buffer for flow cytometry procedures.

### Flow cytometry and fluorescence activated cell sorting

In order to minimize the loss of cell viability, fresh cell suspensions prepared shortly before flow cytometry were used in all cell sorting experiments. Antibody staining was performed in PBS supplemented with 1% heat-inactivated fetal bovine serum. To minimize nonspecific binding of antibodies, cells were first incubated with Human TruStain FcX™ (Fc Receptor Blocking Solution) (BioLegend Inc., USA) for 10 min at a concentration of 3-10 × 10^5^ cells/100 μl. Cells were subsequently stained without washing with antibodies at dilutions recommended by manufacturers. Antibodies used in this study were selected based on previous study^7^ and included: mouse anti-human CD326/EpCAM-FITC (clone VU-1D9, RRID: AB_2534535m Life Technologies, USA), mouse anti-human CD44-APC (clone G44-26, RRID: AB_395868, BD Biosciences, USA), mouse anti-human CD66a-PE (clone 283340, R&D Systems, USA). Cells positive for expression of non-epithelial lineage markers were excluded by staining with mouse anti-human CD45-APC-Cy7 (clone 2D1, RRID: AB_2566375, Biolegend Inc., USA) (see gating strategy in **Supplementary Figure S1a**). After 20 min, stained cells were washed of excess unbound antibodies and resuspended in PBS supplemented with 1% heat-inactivated fetal bovine serum. Flow-cytometry analysis was performed using a CyFlow® Space cell sorter (Sysmex Partec, Germany) (see representative plots of flow cytometry data in **Supplementary Figure S1b-d**). In cell sorting experiments, each population of interest was sorted individually, using a protocol already built-in within the CyFlow® Space flow cytometer software package (FloMax® 2.8), with appropriate adjustments. Data was analysed and visualised using FlowJo™ v10.7 (BD FlowJo, USA). Two cell populations (CD45^-^ /EpCAM^+^/CD44^+^/CD66a^+^ and CD45^-^/EpCAM^+^/CD44^-^/CD66a^+^) were further described based on the literature and single-cell RNA-seq dataset of the human colon, available in GEO database with accession number GSE116222 (see the single-cell RNA-seq data-derived expression plot of three main cell surface markers in **Supplementary Figure S2**). CD45^-^ /EpCAM^+^/CD44^+^/CD66a^-^ population was entitled as crypt-bottom cells (consisting of undifferentiated colonic epithelial cells as well as secretory Goblet and enteroendocrine cells), while CD45^-^/EpCAM^+^/CD44^-^/CD66a^+^ population was entitled as crypt-top cells (consisting of absorptive colonocytes and BEST4^+^/OTOP2^+^ cells). Sorted cell samples were frozen and stored at −70 °C prior to total RNA extraction.

### Total RNA extraction

Total RNA extraction from colon biopsies and sorted cells was performed using standard protocols of commercial miRNeasy Mini Kit (Qiagen, Germany) and Single Cell RNA Purification Kit (Norgen, Canada), respectively. Total RNA concentration was evaluated by NanoDrop2000 spectrophotometer (Thermo Scientific, USA) and Qubit 4 fluorometer (Invitrogen, USA). Total RNA quality was assessed using Agilent 2100 Bioanalyzer (Agilent Biotechnologies, USA).

### Quantitative Reverse Transcription PCR and data analysis

To estimate the expression of *IL-4* and *IL-13* genes in colon tissue of UC patients, total RNA from colon tissue samples was reverse transcribed using a High-Capacity cDNA Reverse Transcription Kit (Applied Biosystems™, USA). Further, expression levels were measured using TaqMan™ Gene Expression Assays (Assay IDs: *IL-4* Hs00174122_m1; *IL-13* Hs00174379_m1) on the 7500 Fast Real-Time PCR System (Applied Biosystems™, USA). The cycle threshold (C_T_) values of *IL-4* and *IL-13* were normalized to the value of *GAPDH* (Assay ID: Hs99999905_m1) reference gene. All the procedures were performed in accordance with the manufacturer’s protocol. Statistical analysis was performed using R studio software (version 4.0.3). Data distribution was determined using the Shapiro-Wilk test, gene expression differences were analysed the two-sided Mann-Whitney U test. The difference between the values was considered significant when P < 0.05.

### Preparation of small RNA libraries and next-generation sequencing

Small RNA libraries from tissue samples were prepared using TruSeq Small RNA Sample Preparation Kit (Illumina, USA) with 1 μg of total RNA input per sample. Small RNA libraries from sorted cells were prepared using NEXTFLEX Small RNA-seq Kit v3 (Bioo Scientific, USA) with up to 50 ng of total RNA input per sample. Procedures were conducted according to the manufacturers’ protocols. The yield of sequencing libraries was assessed using the Agilent 2200 TapeStation system (Agilent Biotechnologies, USA). Subsequently, the TruSeq libraries were pooled with around 24 samples per lane, while the NEXTFLEX libraries were pooled with around 16 samples per lane and then sequenced on HiSeq 4000 (Illumina, USA) next-generation sequencing platform.

### Bioinformatics and statistical analysis of small RNA-seq data

The demultiplexed raw reads (.fastq) were processed with nf-core/smrnaseq v1.0.0 best-practise analysis pipeline using default parameters (Nextflow version v20.01.0)^8^. The pipeline was executed within a Docker container. Briefly, depending on the small RNA-seq library preparation kits, “illumina” or “nextflex” protocol was selected for processing of the libraries generated from tissue and sorted cell samples, respectively. First, Trim Galore (v0.6.3) was used to remove 3’ adapter (5’ - TGGAATTCTCGGGTGCCAAGG - 3’ for both “illumina” and “nextflex” protocols) sequences from the reads and additional 4 nucleotides from both 5’ and 3’ ends of reads (only for “nextflex” protocol). Second, to reduce computational time, the reads with identical sequences were collapsed using seqcluster^9^ while saving the information about the read counts. Third, Bowtie1 v1.2.2^10^ was used to perform the alignment of collapsed reads against mature and hairpin miRNA sequences in miRbase database v22.1^11^. Finally, miRNA annotation was performed using the mirTOP v0.4.23^12^. Further, sample and miRNA QC was performed: samples with initial read count < 1.5 IQR and number of detected miRNAs < 1.5 IQR on log_2_ scale as well as non-expressed (mean raw < 1) and non-variable miRNAs were excluded from further analysis. Subsequently, to perform differential expression analyses of the size factor normalized counts of mature miRNAs between samples, negative binomial generalized linear models implemented in the R package DESeq2^13^ were used including age (scaled and centred) and sex as covariates in the model. The *P*-values resulting from Wald tests were corrected for false discovery rate (FDR) according to Benjamini and Hochberg. The miRNAs with FDR < 0.05 and absolute value of log_2_FC > 1 were considered to be significantly differentially expressed. A multidimensional scaling (MDS) analysis using Euclidean distance was performed on variance stabilizing transformation (VST) normalized miRNA count data. Additionally, Spearman’s rank correlation analysis was performed between sex- and age-adjusted normalized miRNA read counts and endoscopic Mayo subscore. FDR < 0.05 was considered statistically significant. Removal of sex and age effects from normalized data was performed using removeBatchEffect function from the limma R package^14^. Statistical analyses and data processing were performed using R version 4.0.3^15^. Visualization of graphs was performed using the ggplot2 package^16^.

### Gene set enrichment analysis

In order to obtain putative biological functions of differentially expressed miRNAs, gene set enrichment analysis (GSEA) was performed using Reactome pathways^17^ and GO categories^18^. More specifically, luciferase assay-validated miRNA-target interactions (MTIs) were obtained from miRecords^19^, miRTarBase^20^ and TarBase^21^ using the multiMiR package^22^. The retrieved MTIs were then submitted to a hypergeometric test implemented in the enrichPathway (from the ReactomePA package^23^) and enrichGO (from the clusterProfiler package^24^) functions using genes that are expressed in colon crypt-bottom (CD44^+^) and crypt-top (CD66a^+^) cells as a background reference (defined as universe). The universe genes were obtained from single-cell RNA-seq data of the human colon, available in GEO database with accession number GSE116222^25^. The pathways with FDR < 0.05 were considered to be significantly deregulated.

### miRNA co-expression analysis

Samples of study group II were used for weighted gene co-expression network analysis (WGCNA) aiming to identify modules of co-expressed miRNAs using CEMiTool package^26^ (version 1.22.0) for R. Normalized (variance stabilizing transformation) miRNA count table was used to generate the co-expression modules. Filtering based on variance was applied on gene expression table prior to identification of miRNA co-expression modules. Minimal number of miRNAs per submodule as well as minimum size of gene sets for GSEA analysis was set to five, while *P*-value threshold for filtering was set to 0.1. Further, the eigengene value of the co-expression module identified in colonic epithelial cells was also calculated in colon tissue data using the expression table of co-expressed miRNAs and WGCNA package^27^ (version 1.72-1) for R. Next, Spearman’s rank correlation coefficients were calculated between module eigengene values and endoscopic Mayo subscore in both colonic epithelial cell populations as well as in colon tissue. FDR < 0.05 was considered statistically significant. Finally, to assess the performance of the identified module eigengene value for distinguishing between the active and quiescent UC in both colon tissue and distinct colonic epithelial cell populations, analysis of Area Under the Receiver Operating Characteristic curve (AUC-ROC) was applied using pROC package^28^ (version 1.18.4) for R.

## Results

### Differentially expressed miRNAs in active and quiescent UC tissues are involved in regulation of inflammation-related pathways

To identify differentially expressed miRNAs and their putative regulatory processes during chronic colon inflammation, small RNA-seq was performed on inflamed (active) and non-inflamed (quiescent) colonic mucosal biopsies of UC patients and healthy controls (HC) (**Figure 1a**).

**Figure 1:**
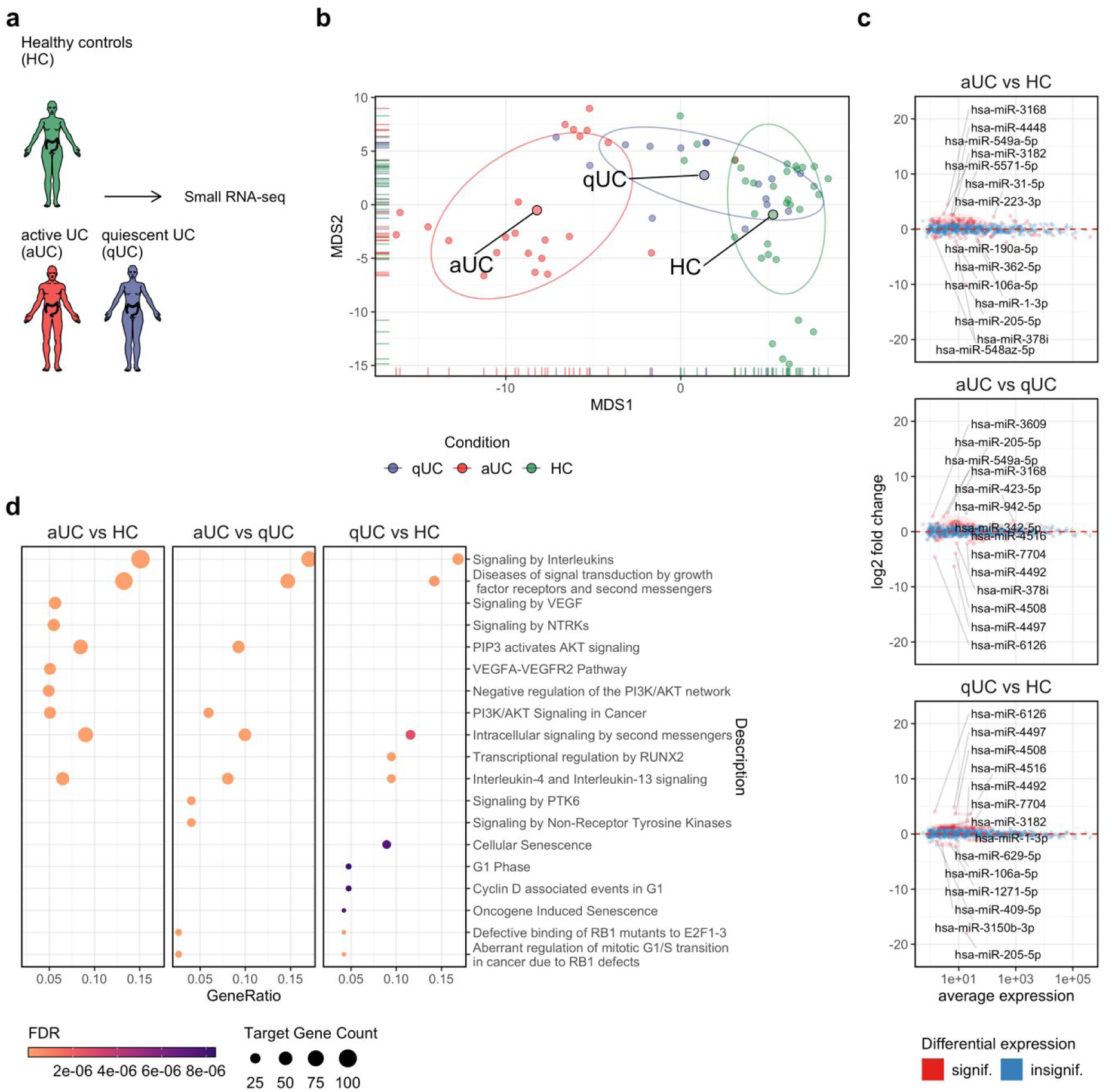
Small RNA-seq defines differentially expressed miRNAs involved in inflammation-associated pathways in active and quiescent UC tissues. **(a)** Design of the initial study phase. **(b)** MDS plot showing the similarity structure of the miRNA transcriptomes in active (aUC) (n=23), quiescent UC (qUC) (n=20) and HC (n=30) tissue samples based on normalized expression values. The dots represent samples coloured by group. The centroid of ellipses corresponds to the group mean; the shapes are defined by covariance within a given group. **(c)** Differentially expressed miRNAs in active (aUC) (n=23) and quiescent UC (qUC) (n=20). The red colour represents significantly (FDR < 0.05) differentially expressed miRNAs with an absolute value of log_2_FC > 1, while the blue colour represents non-differentially expressed miRNAs. **(d)** Top 10 overrepresented pathways in active (aUC) (n=23) and quiescent UC (qUC) (n=20) tissues identified by miRNA set enrichment analysis. Dot size represents the number of miRNA-target gene count in the significantly enriched (FDR < 0.05) Reactome pathways.

After count data normalization and quality control, 573 unique miRNAs were found to be expressed in colon tissue samples. The overall similarity structure (based on MDS analysis; see **Methods**) of colon miRNA transcriptomes revealed two clearly resolved clusters corresponding to active UC and HC tissues, while the third cluster corresponding to quiescent UC overlapped with both active UC and HC clusters, suggesting a shift of miRNA expression from healthy to inflammatory state (**Figure 1b**). To evaluate expression of specific miRNAs in active UC and quiescent UC, differential gene expression analysis was performed. As expected, the most profound miRNA deregulation was observed comparing active UC to HC or to quiescent UC (93 and 59 differentially expressed miRNAs [FDR < 0.05 and |log_2_FC| > 1], respectively). Interestingly, although substantially lower than in active UC, a differential expression of miRNAs (n=32) was also observed in quiescent UC compared to HC (**Figure 1c**; **Supplementary Table S2**). Among the differentially expressed miRNAs, a considerable number (n=13) of molecules, including miR-106-5p, miR-125b-1-3p, miR-205-5p and miR-3182 were deregulated in both active and quiescent UC compared to HC. Contrarily, majority of miRNAs (n=80) including, miR-190a-5p, miR-3168, miR-378i, and miR-223-3p were deregulated only in active UC, while miRNAs (n=19) such as miR-331-3p, miR-409-5p, miR-4497 and miR-629-5p - only in the quiescent UC. In addition, the gradual decrease in miR-1-3p expression was found in all pairwise comparisons (active UC vs quiescent UC, active UC vs HC and quiescent UC vs HC; **Supplementary Table S2**). Thus, besides shared miRNA deregulation patterns, the data also suggests unique and perhaps non-overlapping functions of some miRNAs in active and quiescent UC.

To further determine the biological function of differentially expressed miRNAs in UC pathogenesis, GSEA was performed for each pairwise comparison (active UC vs HC; quiescent UC vs HC; active vs quiescent UC) using validated target genes of significantly deregulated miRNAs and Reactome pathways (**Supplementary Table S3**). Intriguingly, both active UC and quiescent UC compared to HC had over-represented interleukin signalling-related pathways among the top significant ones, such as “*Signaling by Interleukins*” [R-HSA-449147] and “*Interleukin-4 and Interleukin-13 signaling*” [R-HSA-6785807] (**Figure 1d**). To further evaluate this finding, the expression patterns of two main cytokines IL-4 and IL-13 in colon tissue was analysed. The gradual increase in *IL-13* expression was observed between UC patients and HC individuals (2.19-fold [P = 0.031] increase in quiescent UC vs. HC, 2.91-fold [P = 0.0007] increase in active UC vs. quiescent UC, 6.38-fold [P = 2 × 10^−10^] increase in active UC vs. HC), whereas the expression of *IL-4* did not differ between the groups (**Supplementary Figure S3**). *In silico* (enrichment in the interleukin signalling-related target genes), as well as *in vivo* (interleukin expression in colon tissue) analysis confirmed that these pathways remain dysregulated in the colonic mucosa during the disease course of UC.

Taken together, differential miRNA expression and GSEA analyses in UC tissues suggest that dysregulated miRNAs are likely involved in regulation of inflammation-related pathways and that some of the pathways, i.e., interleukin-4 and interleukin-13 signalling, remarkably, remain dysregulated in the colonic mucosa of quiescent UC.

### Sequencing of FACS-sorted colonic epithelial cells shows cell type-specific miRNA expression during colonic inflammation in UC

The role of interleukin-4 and interleukin-13 in mediating permeability of epithelial barrier as well as its relation to the pathogenesis of UC has been previously described^29^. Given that this pathway is deregulated in active UC tissues and remains deregulated in quiescent UC (based on miRNA and mRNA expression levels in our study) and the fact that it affects epithelial barrier function, we further decided to focus on miRNA expression in crypt-bottom (CD44^+^) and crypt-top (CD66a^+^) colonic epithelial cells.

FACS was applied to select and enrich for crypt-bottom and crypt-top colonic epithelial cells from active and quiescent UC patients and healthy controls using CD44^+^ and CD66a^+^ surface markers (**Supplementary Figure S1**). Analysis of FACS enriched cell populations over single-cell analysis was chosen due to limitations of single-cell transcriptome sequencing technology, which is not yet compatible with small RNA-seq. Interestingly, analysis of flow cytometry showed significant (FDR < 0.05) increase in crypt-bottom (CD44^+^) cells in active UC compared to HC (**Figure 2a**), suggesting a potential inflammation-stimulated cell proliferation^30,31^.

**Figure 2:**
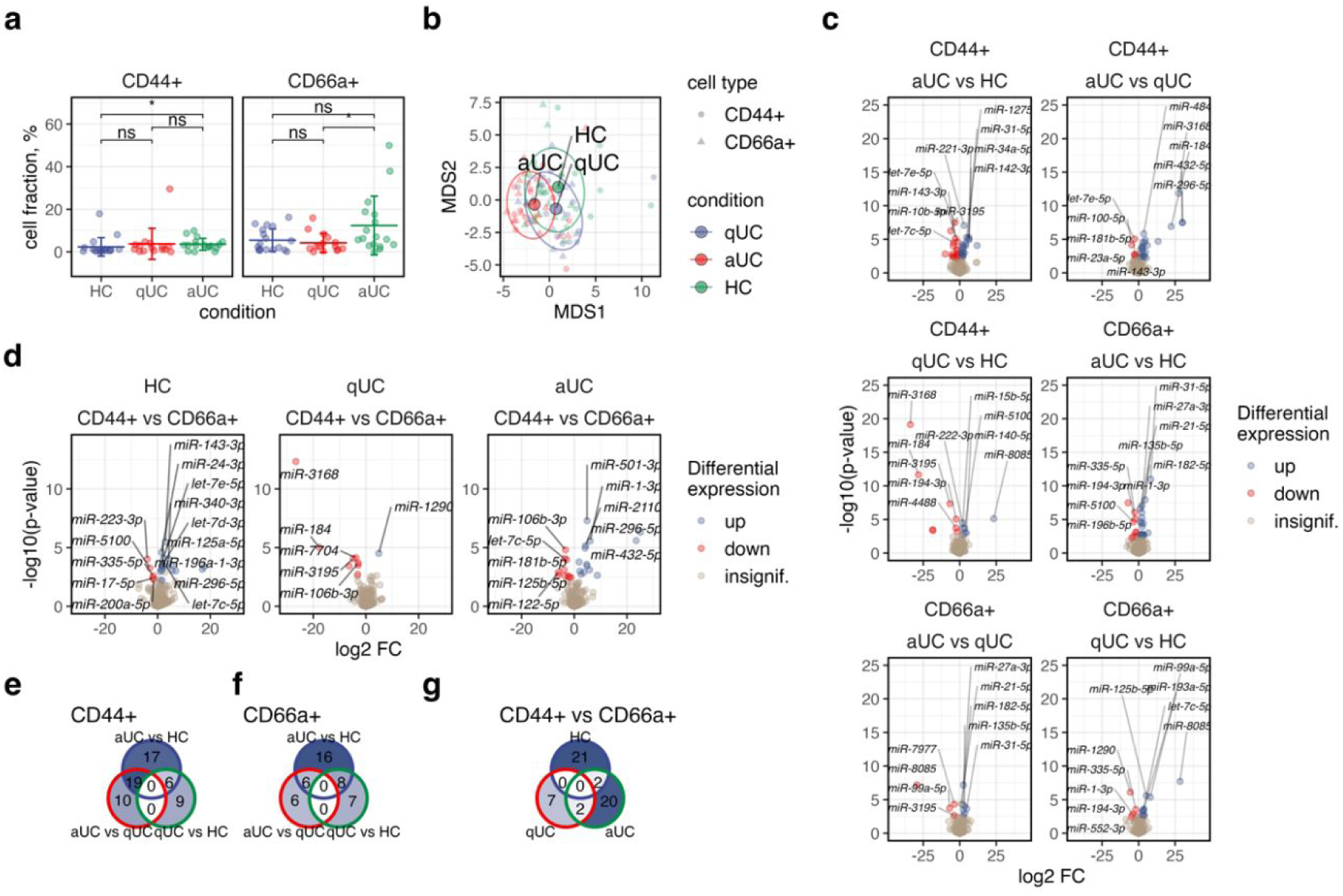
Sequencing data reveals differentially expressed miRNAs in colonic epithelial cells of patients with active and quiescent UC. **(a)** FACS of colonic epithelial cells and distribution of crypt-top CD44^-^CD66^+^ and crypt-bottom CD44^+^CD66^-^ epithelial cell types in inflammatory (aUC) (n=16) and non-inflammatory (qUC and HC) (n=15 and 17) colon tissues. Each dot represents a sample from the patients of the second study group. Mean ±SD of each group is represented by vertical lines. To compare the groups a nonparametric Mann–Whitney U test was performed, p* < 0.05. **(b)** MDS plot showing the similarity structure of the miRNA transcriptomes in crypt-top (CD66a^+^) and crypt-bottom (CD44^+^) colonic epithelial cell populations in active (aUC) (n=16), quiescent UC (qUC) (n=15), and in HC (n=17) based on normalized expression values. The dots represent samples shaped by cell population. Dot colours represent condition. The centroid of ellipses corresponds to the condition group mean; the shapes are defined by covariance within the group. **(c, d)** Volcano plots of differentially expressed miRNAs in crypt-top (CD66a^+^) and crypt-bottom (CD44^+^) colonic epithelial cell populations in active (aUC) (n=16), quiescent UC (qUC) (n=15), and HC (n=17). Colours indicate significantly (FDR < 0.05) differentially expressed miRNAs with an absolute value of log_2_FC > 1 between compared groups. **(e-g)** Venn diagrams representing the numbers of commonly and uniquely differentially expressed miRNAs in **(e)** crypt-bottom (CD44^+^) and **(f)** crypt-top (CD66a^+^) epithelial cell populations in different UC activity, and **(g)** between crypt-bottom and crypt-top cells in the same condition.

Using sequencing, a total number of 436 unique miRNAs were found to be expressed in crypt-bottom (CD44^+^) and crypt-top (CD66a^+^) epithelial cells. Although miRNA transcriptomes of these colonic epithelial cell populations were strongly overlapping (**Figure 2b**), significant changes in expression profiles were observed within and between cell populations in different stages of the disease (**Figure 2c and 2d**; **Supplementary Table S4**). Initially, pairwise comparisons were performed in the same epithelial cell population to identify UC inflammation-associated miRNAs. As in the tissue data, the number of differentially expressed miRNAs (FDR < 0.05 and |log_2_FC| > 1) in both colonic epithelial cell populations were gradually increased depending on the disease activity, i.e., the highest number of differentially expressed miRNAs were observed in active UC compared to HC and to quiescent UC (**Figure 2c**). Among deregulated molecules, none of miRNAs were commonly differentially expressed across all three comparisons (active UC vs HC; quiescent UC vs HC; active UC vs quiescent UC) in both crypt-bottom (CD44^+^) and crypt-top (CD66a^+^) colonic epithelial cells. However, six miRNAs (miR-15b-5p, miR-222-3p, miR-223-3p, miR-194-3p, miR-3195 and miR-574-3p) were identified as commonly differentially expressed in crypt-bottom (CD44^+^) cells and eight miRNAs (let-7c-5p, miR-106b-3p, miR-125b-5p, miR-1-3p, miR-1290, miR-194-3p, miR-335-5p, miR-552-3p) - in crypt-top (CD66a^+^) cells in the active and quiescent UC when compared to HC (**Figure 2e and 2f**). Further, the response of distinct epithelial cell populations to inflammation was determined by performing pairwise comparisons with the separate populations of colonic epithelial cells. Twenty-four miRNAs (such as miR-501-3p, miR-1-3p, miR-296-5p, and miR-122-5p) were identified to be differentially expressed in active UC, nine miRNAs (such as miR-1290, miR-3168, and miR-660-5p) in quiescent UC, and twenty-two miRNAs (such as miR-598-3p, miR-340-3p, and miR-223-3p) - HC when compared crypt-bottom (CD44^+^) and crypt-top (CD66a^+^) colonic epithelial cells (**Figure 2d, Supplementary Table S4**). Notably, the vast majority of identified differentially expressed miRNAs between distinct epithelial cell types in different stages of inflammation were found to be uniquely dysregulated. Only two commonly differentially expressed miRNAs were observed between two comparison groups (miR-106b-3p and miR-1290 in active UC CD44^+^ vs CD66a^+^ and quiescent UC CD44^+^ vs CD66a^+^; miR-296-5p and miR-432-5p active UC CD44^+^ vs CD66a^+^ and HC CD44^+^ vs CD66a^+^) (**Figure 2g**). This suggests that even at different disease activity stages there are cell population-specific responses, in terms of miRNA expression.

### Aberrantly expressed miRNAs in crypt-top (CD66a^+^) and crypt-bottom (CD44^+^) colonic epithelial cells are involved in regulation of inflammation and intestinal epithelial barrier function-related processes

To further evaluate the potential functional role of differentially expressed miRNAs in crypt-bottom (CD44^+^) and crypt-top (CD66a^+^) colonic epithelial cells in UC pathogenesis, GSEA was performed on validated target genes of differentially expressed miRNAs (**Supplementary Table S5**). Similarly, to the colonic tissue data (**Figure 1b**), majority of the overrepresented pathways in both colonic epithelial cell populations in both stages of disease activity overlapped and included signalling pathways such as “*Signaling by Interleukins*” (R-HSA-449147), “*Interleukin-4 and Interleukin-13 signaling*” [R-HSA-6785807], “*Signaling by Receptor Tyrosine Kinases*” [R-HSA-9006934] (**Figure 3a**). This supports that observations in the colon biopsy samples were mainly driven by colonic epithelial cells. Notably, not all comparison groups in crypt-bottom (CD44^+^) cells showed uniform results. The most overrepresented Reactome pathways in crypt-bottom (CD44^+^) cells of quiescent UC patient group differed from those in active UC groups and exclusively included *“Signaling by Nuclear Receptors”* (R-HSA-9006931), *“Extra-nuclear estrogen signaling”* [R-HSA-9009391] pathways which were shown to regulate the permeability of intestinal epithelium and thus be associated with pathogenesis of inflammatory bowel disease^32^ (**Figure 3a**). Finally, to assess which biological processes (from Gene Ontology terms) are overrepresented in crypt-bottom (CD44^+^) compared to crypt-top (CD66a^+^) cells, GSEA of deregulated miRNAs in active UC, quiescent UC, and HC groups was performed. The results revealed that overrepresented processes between the cell populations were mainly related to cell differentiation and motility in both active UC and HC (**Figure 3b**), suggesting that in inflamed and healthy colon mucosa these pathways are differentially regulated between the cell types. In addition, significant enrichment between crypt-bottom (CD44^+^) and crypt-top (CD66a^+^) cells in “*epithelium migration”* [GO:0090132] and *“epithelial cell migration”* [GO:0010631] were uniquely identified only in active UC among the most overrepresented biological processes (**Figure 3b**). Target genes of differentially expressed miRNAs between cell populations in the UC remission group were mainly related to cell migration and were least different between those populations (**Figure 3b**).

**Figure 3:**
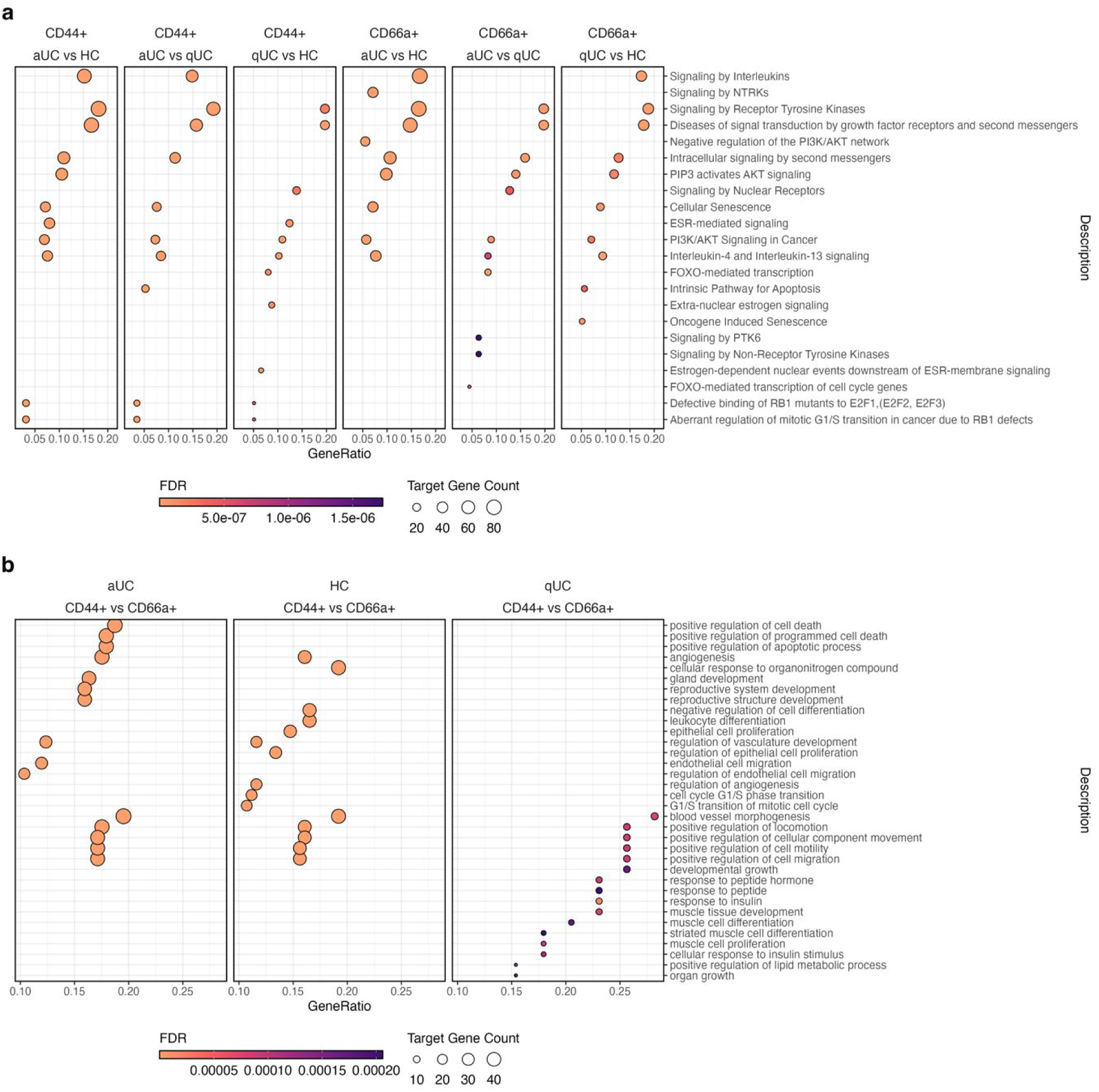
Aberrantly expressed miRNAs of crypt-top (CD66a^+^) and crypt-bottom (CD44^+^) colonic epithelial cells are involved in UC-related processes. Top 10 overrepresented pathways within **(a)** and between **(b)** crypt-top (CD66a^+^) and crypt-bottom (CD44^+^) colonic epithelial cell populations during active (aUC) (n=16), quiescent UC (qUC) (n=15) and in controls (HC) (n=17) identified by miRNA-target gene set enrichment analysis. Dot size represents the number of miRNA gene-target count in the significantly enriched (FDR < 0.05) Reactome pathways **(a)** and GO categories **(b)**.

To summarize, despite the significant overlap of aberrantly expressed miRNAs of both colonic epithelial cell populations in regulatory signalling pathways, GSEA results reveal unique involvement of miRNAs in UC-, inflammation- and intestinal barrier integrity-related processes in different stages of disease activity and/or cell population.

### miRNAs in crypt-top (CD66a^+^) and crypt-bottom (CD44^+^) colonic epithelial cells exhibit co-expression pattern which is related to UC activity

First, to unveil the relation between individual miRNA expression levels and endoscopic Mayo subscore in crypt-top (CD66a^+^) and crypt-bottom (CD44^+^) colonic epithelial cells, Spearman correlation analysis was applied. In crypt-bottom (CD44^+^) and crypt-top (CD66a^+^) colonic epithelial cells, a number (n=34 and n=23, respectively) moderate positive (0.4 < rho < 0.7; FDR < 0.05) and a few (n=6 and n=7, respectively) moderate negative (−0.7 < rho < −0.4; FDR < 0.05) correlations were observed among the normalized miRNA expression levels and endoscopic Mayo subscore (**Supplementary Figure S5; Supplementary Table S6; Supplementary Table S7)**. Noteworthy, analysis resulted not only in a substantial overlapping in disease activity-associated miRNAs between both colonic epithelial cell populations (29 common moderately correlating miRNAs), but also revealed few cell population-unique correlations.

Subsequently, we decided to perform more complex analysis and evaluate if certain colonic epithelial cell miRNAs of both populations are co-expressed together. First, WGCNA performed on all miRNAs of colonic epithelial cell populations uncovered the miRNA co-expression network (**Figure 4a**) and identified two co-expression modules (M1 and M2). Module M1 was comprised of 13 miRNAs (miR-10b-5p, miR-182-5p, miR-146a-5p, miR-196b-5p, miR-222-3p, miR-27a-3p, miR-221-3p, miR-194-3p, miR-183-5p, miR-223-3p, miR-574-5p, miR-135b-5p, miR-31-5p); while module M2 consisted of 11 miRNAs such as let-7b-5p, miR-143-3p, miR-125a-5p, miR-15b-5p, let-7e-5p, miR-5100, miR-181b-5p, miR-1-3p, miR-125b-5p, miR-100-5p, miR-195-5p. Module enrichment analysis based on the evaluation of normalized enrichment score (NES) (**Figure 4b**) further revealed that module M1 was significantly enriched in both crypt-top (CD66a^+^) and crypt-bottom (CD44^+^) epithelial cells of patients with active UC (NES = 1.71 [P_adj._ = 9.7 × 10^−3^] and 1.67 [P_adj._ = 2.9 × 10^−2^], respectively), while in both cell populations of control group individuals it was decreased (NES = −1.79 [P_adj._ = 7.7 × 10^−3^] and −1.74 [P_adj._ = 5.0 × 10^−2^], respectively). Interestingly, the enrichment values for the module M2 were opposite to what was observed for M1, meaning the normalized expression of module M2 in both crypt-top (CD66a^+^) and crypt-bottom (CD44^+^) epithelial cell populations of patients with active UC was significantly decreased (NES = −1.84 [P_adj._ = 9.7 × 10^−3^] and −1.80 [P_adj._ = 2.9 × 10^−2^], respectively), while it was significantly enriched exclusively in crypt-bottom (CD44^+^) cells of patients with quiescent UC (NES = 2.08 [P_adj._ = 3.3 × 10^−4^]).

**Figure 4:**
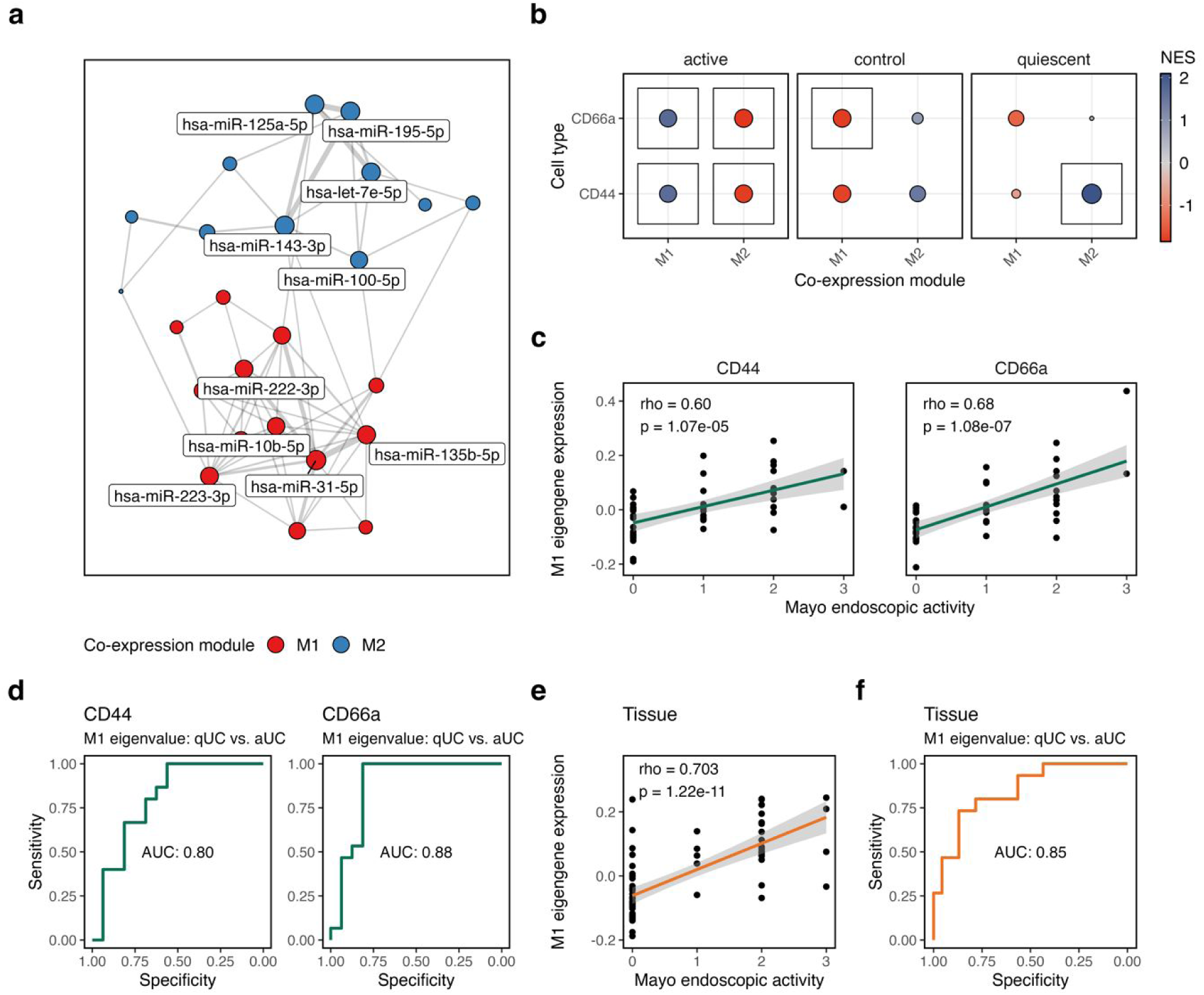
WGCNA in colonic epithelial cell populations reveals miRNA co-expression module which reflects endoscopic activity of UC. **(a)** Network displaying identified co-expression modules (M1 and M2) in crypt-top (CD66a) and crypt-bottom (CD44) colonic epithelial cells of patients with active (aUC) (n=16), quiescent UC (qUC) (n=15) and control individuals (n=17). The color and size of the node represents distinct modules and strength of connectivity, respectively. **(b)** The dot plot showing normalized enrichment score (NES) of the modules M1 and M2 in crypt-top (CD66a) and crypt-bottom (CD44) colonic epithelial cells of patients with active, quiescent UC and control individuals. The color and size of the dot represents the value and absolute value of NES, respectively. The box marks significant value (p_adj,_ < 0.05). Plots **(c)** and **(e)** show the correlation between module M1 eigengene value and Mayo endoscopic activity in crypt-bottom (CD44) and crypt-top (CD66a) colonic epithelial cells as well as colon tissue, respectively. rho – Spearman correlation coefficient. Each dot represents different sample. Plots **(d)** and **(f)** show the AUC-ROC curves reflecting the performance of M1 module eigengene value at distinguishing between active (aUC) and quiescent UC (qUC) in crypt-bottom (CD44) and crypt-top (CD66a) colonic epithelial cells as well as in colon tissue, respectively.

Further, we focused on potentially pro-inflammatory module M1 and explored whether the expression of this module in each of the studied epithelial cell populations is associated with clinical characteristics of UC. Therefore, Spearman correlation analysis was applied to check for the association between endoscopic Mayo subscore and module M1 eigengene value (summarized module expression value). Analysis resulted in the significant moderate positive correlation in both crypt-top (CD66a^+^) and crypt-bottom (CD44^+^) cells (rho = 0.68 [P = 1.08 × 10^−7^] and 0.60 [P = 1.07 × 10^−5^], respectively) (**Figure 4c**). Next, AUC-ROC analysis was employed to assess the performance of the module M1 eigengene value at distinguishing between the active and quiescent UC in each of the studied colonic epithelial cell populations. In the crypt-bottom (CD44^+^) cells, analysis resulted in the AUC value of 80.0% (confidence interval (CI): 63.6% - 96.4%), while in the crypt-top (CD66a^+^) cells M1 eigengene expression value demonstrated even better performance and showed the AUC value of 87.9% (CI: 74.0% - 100.0%) when separating quiescent *vs.* active UC (**Figure 4d**). To evaluate the concordance of the epithelial cell population-derived data with the situation in the colon tissue, we calculated module M1 eigengene values in the tissue and performed analogous analysis as in the crypt-top and crypt-bottom cell populations. Briefly, analysis provided almost identical results to those observed in colonic epithelial cell populations with higher similarity to crypt-top cells. More precisely, in colon tissue, module M1 eigenvalue showed strong positive correlation with endoscopic Mayo subscore (rho = 0.703 [P = 1.22 × 10^−11^]) and produced AUC value of 85.0% (CI: 72.2% - 97.1%).

Generally, the results uncovered potential UC endoscopic activity-related miRNA co-expression patterns that are not only characteristic for both crypt-top and crypt-bottom colonic epithelial cell populations, but also reflect the overall situation in the more heterogenous colon tissue.

## Discussion

Although UC is a well-studied complex disease and huge efforts have been made to explore its molecular mechanisms, the disease pathogenesis still remains largely unclear^33^. Especially, there is a substantial knowledge gap about expression patterns of regulatory non-coding miRNA in UC in a cell type-specific context. Thus, here we provide an overview of the miRNA expression in the unsorted whole colonic mucosa samples of UC patients and present detailed colonic epithelial cell population-specific miRNA expression profiles from active and quiescent UC patients and describe differences in miRNA expression patterns between two distinct - crypt-bottom (CD44^+^) and crypt-top (CD66a^+^) - cell populations. Furthermore, we describe putative biological pathways in which deregulated miRNAs of UC colonic epithelial cell populations might be involved, identify potential inflammatory miRNA co-expression module, determine its associations with endoscopic disease activity and evaluate the performance in distinguishing between active and quiescent stages of UC.

Most importantly, we determined distinct responses in miRNA expression of different colonic epithelial cell populations during UC. Our findings showed that in colon crypt-bottom (CD44^+^) cells (compared to crypt-top [CD66a^+^] cells) inflammation promoted/suppressed the expression of several miRNAs possibly involved in cell proliferation, differentiation and/or permeability of intestinal barrier. For example, let-7c-5p showed considerable down-regulation and miR-501-3p up-regulation in crypt-bottom (CD44^+^) cells during active UC. It has previously been shown that overexpression of let-7c-5p as well as inhibition of miR-501-3p can reduce the proliferation of colorectal cancer cells^34,35^. Thus, deregulation of these miRNAs might be related to the relative increase of the crypt-bottom (CD44^+^) cells in active UC when compared to controls, as it has been shown in our flow cytometry experiment. On the other hand, we observed increased expression of miR-1-3p and decreased expression of miR-125b-5p in crypt-bottom compared to crypt-top cells only during active UC. Both miRNAs were shown to be involved in barrier function dysregulation, where decrease of miR-125b-5p^36^ and increase of miR-1-3p^37^ contribute to disruption of epithelial barrier in colon tissue. This would suggest that epithelial barrier is already impaired in crypt-bottom epithelial cells during active UC; however, it remains unclear if this is a UC specific event or rather a normal cell response to inflammation in the gut.

Furthermore, our analysis showed that not only certain individual miRNAs are associated with the endoscopic Mayo subscore, but they also form an inflammation-related co-expression network, which directly correlates with clinical disease activity. Of two identified miRNA co-expression modules in both crypt-bottom (CD44^+^) and/or crypt-top (CD66a^+^) cells, module M1 was significantly enriched in both epithelial cell populations of active UC patients and was comprised of such miRNAs as miR-31-5p, miR-135b-5p, miR-27a-3p, miR-222-3p, miR-223-3p, etc. In the literature, these small non-coding RNAs are also reported to be upregulated in inflamed, non-inflamed and/or pre-cancerous colon tissues as well as in faeces (specifically, miR-223-3p) of UC patients^38–43^ and are involved in the regulation of inflammatory response (e.g. miR-222-3p targets *SOCS1* and activates STAT3 signalling)^44–46^. Additionally, we observed that all miRNAs belonging to module M1 were involved in Reactome pathways related to interleukin signalling by targeting various validated genes (such as *FOXO3*, *IGF1R, ICAM1, STAT6, STAT1, STAT5A, CCND1*, etc.). Interestingly, when comparing crypt-top and crypt-bottom cell populations-derived module M1 performance and association results to colon tissue, both the correlation coefficient and AUC values in tissue were more like those observed in crypt-top (CD66a^+^) cells. This may be at least partially explained by the cellular composition of the colon mucosa, the most abundant cell type in mucosal layer being absorptive colonocytes^47^. Of note, even though the second identified miRNA co-expression module M2 did not show meaningful associations with clinical characteristics of UC, its exclusive enrichment in crypt-bottom (CD44^+^) colonic epithelial cells of patients with quiescent disease provides novel insights in regulatory mechanisms during UC. For example, among module M2 miRNAs we identified let-7e-5p, the individual expression of which also negatively correlated with the endoscopic Mayo subscore exclusively in crypt-bottom (CD44^+^) colonic epithelial cells. Additionally, in the intestinal epithelium let-7 family member let-7b appears to be among the highest-expressed miRNAs in let-7 group/family (**Supplementary Figure S4**). It has previously been shown that expression of let-7 miRNAs (especially let-7e) is increased and affects maintenance of cell differentiation^48^. Altogether, these observations suggest the relevance of let-7 miRNAs during intestinal inflammation via maintenance of stemness of crypt-bottom (CD44^+^) cells and thereby explain their expression correlation with disease activity, exclusively in undifferentiated colonic epithelial cells.

Finally, we described potential involvement of differentially expressed miRNAs in regulatory biological pathways. Our initial small RNA-seq of colonic mucosa biopsies from active and quiescent UC compared to healthy controls, at first, revealed multiple deregulated miRNAs, which were significantly enriched in inflammation- and intestinal epithelial barrier function-related biological pathways. Interestingly, miRNAs enriched in interleukin biological pathways, such as interleukin-4 and interleukin-13 signalling, were deregulated not only in active, but also in quiescent UC, suggesting lasting derangement of this pathway in mucosa of UC. The interleukin-4 and interleukin-13 pathway is known to differentially regulate epithelial chloride secretion and cause epithelial barrier dysfunction^49^. It has been shown that large amounts of interleukin-13 are produced in colon mucosa of UC patients and thereby impair epithelial barrier function by affecting epithelial apoptosis, tight junctions, and restitution velocity^29,50^. We also confirmed the increased expression of interleukin-13 gene during the course of UC when analysing active and quiescent UC patient colon tissue samples. Contrarily, some studies report decreased mucosal amounts of interleukin-13 in active UC^51^. Nevertheless, the attempts are still being made to adapt the inhibition of interleukin-13-based treatment to induce UC remission (e.g., clinical trials of anti-interleukin-13 monoclonal antibodies (tralokinumab and anrukinzumab)^52^ and preclinical studies of anti-interleukin-Rα2^53^). Our findings on differentially expressed miRNA involvement in interleukin-4 and interleukin-13 regulation even in quiescent UC as well as controversial data in the literature led us to further focus on miRNA expression analysis in less differentiated crypt-bottom (CD44^+^) and fully differentiated absorptive crypt-top (CD66a^+^) colonic epithelial cells, which are responsible for intestinal barrier integrity and permeability^2^. Similarly to the results in colonic biopsies, both crypt-bottom (CD44^+^) and crypt-top (CD66a^+^) epithelial cell populations showed deregulation in miRNAs during UC, the targets of which were significantly enriched in the interleukin-4 and interleukin-13 signalling pathway. Since interleukin-4 and interleukin-13 cytokines are predominantly produced by immune cells^54^, the expected regulatory action of deregulated miRNAs in the colonic epithelial cells would be downstream targets of the pathway, such as *STAT3*, *FOXO3*, *SOCS1*, etc. During active UC in both crypt-bottom (CD44^+^) and crypt-top (CD66a^+^) cells, we found miR-221-3p, miR-182-6p, miR-222-3p and miR-31-5p to be up-regulated, which are known to target *FOXO3* gene^22^. The up-regulation of the aforementioned miRNAs, theoretically, would lead to decreased expression of the *FOXO3* gene, which was already observed in colonic mucosa of UC patients^55^. This in turn, may lead to more severe colonic inflammation during UC^56^. Additionally, both crypt-bottom (CD44^+^) and crypt-top (CD66a^+^) cells of patients with active UC had increased expression of hsa-miR-221-3p and hsa-miR-21-5p, that target *SOCS1* gene^22^. SOCS1 is an important regulator of interleukin-4 signalling, and its forced expression was shown to inhibit interleukin-13 signalling in epithelial cells^57^. In addition to involvement in interleukin signalling pathways, we also detected unique intestinal epithelial barrier function-related processes regulated by disease stage and/or cell population-specific miRNAs. For example, differentially expressed miRNAs, such as miR-222-3p, miR-223-3p, miR-15b-5p, in crypt-bottom (CD44^+^) colonic epithelial cells of quiescent UC could potentially regulate nuclear receptors and extra-nuclear estrogen signalling pathways. These findings fall in accordance with other studies demonstrating the importance of impairment of signal transduction via nuclear receptors in inflammatory bowel disease, as nuclear receptors regulate essential aspects of intestinal barrier functions such as mucus secretion, expression of tight junction proteins and others^32^. Moreover, we observed a few aberrantly expressed miRNAs between crypt-bottom (CD44^+^) and crypt-top (CD66a^+^) cells in active UC, that possibly exert their biological function through regulation of epithelial cell migration, which is known to happen along the crypt-villus axis^58^ and is increased during IBD^59^. Noteworthy, the GSEA results should be treated with caution, since the selection of miRNA targets significantly affects the results^60^. However, currently, there are no methods to solve this issue, since miRNA target prediction as well as its dosage to affect target expression are still rather unsolved problems in the field.

In summary, our study determined crypt-bottom (CD44^+^) and crypt-top (CD66a^+^) colonic epithelial cell-specific miRNA deregulation in UC in cell type- and disease stage-dependent manner. We also revealed cell population-specific miRNA expression patterns and networks as well as their associations with clinical disease activity. Furthermore, we unveiled the potential functional role of differentially expressed miRNAs and observed their possible involvement in biological pathways associated with maintenance of intestinal barrier function in active as well as quiescent UC, in both epithelial cell populations. Together, these observations not only highlight regulatory importance of miRNAs in distinct colonic epithelial cell populations during pathogenesis of UC, but also provide potential miRNA candidates for the development of new treatment strategies to maintain the remission of mucosal inflammation.

## Funding

This work was supported by the Research Council of Lithuania and European Crohn’s and Colitis Organisation (grant numbers S-MIP-20-56 and ECCO Grant 2016, respectively).

## Conflict of Interest

The authors declare no conflict of interest.

## Data Availability

The small RNA-seq data underlying this article are available in Gene Expression Omnibus (GEO) Database and can be accessed with accession numbers GSE185101 (reviewer token: **gxytcgywnvejvit**) and GSE185102 (reviewer token: **ijydywoajxyvxmr**).

## Author contributions

JS, SJ, and JK participated in the concept and design of the study. JK, LVJ, VK, UK, GV and GK participated in patient recruitment and material collection and sample processing. RR, ID, and AZ were responsible for fluorescence-activated cell sorting. AF and SS provided infrastructure for small RNA library preparation and sequencing. RI and SJ were responsible for bioinformatics, statistical analysis, data interpretation and writing the first draft of the manuscript. All authors were members of the writing group and participated in the drafting revision of the manuscript.

---

**Supplementary Figure S1:**
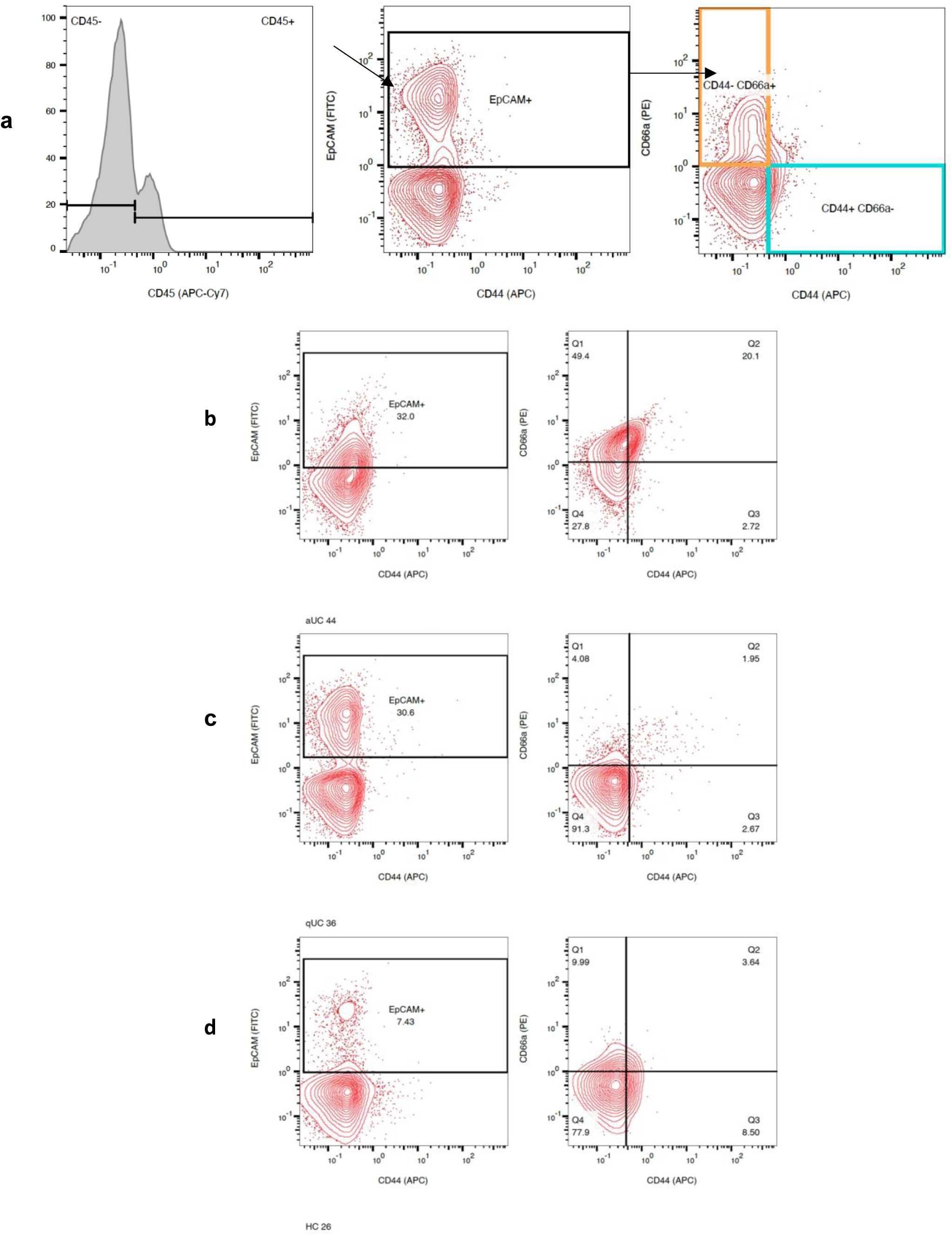
Fluorescence-activated cell sorting (FACS) of crypt-bottom (CD44^+^) and crypt-top (CD66a^+^) colonic epithelial cells from patients with active and quiescent UC and healthy controls (HC). **(a)** Gating strategy for FACS isolation of crypt-top CD44+/CD66a- and crypt-bottom CD44-/CD66a+ epithelial cell populations: selection of epithelial cells by CD45 exclusion, EpCAM inclusion and further isolation of populations by CD44 and CD66a expression. **(b-d)** Representative plots of flow cytometry data showing distribution of CD44+/CD66a- and CD44-/CD66a+ colonic epithelial cell populations during **(b)** active UC (aUC), **(c)** quiescent UC (qUC), and **(d)** in healthy controls (HC).

**Supplementary Figure S2:**
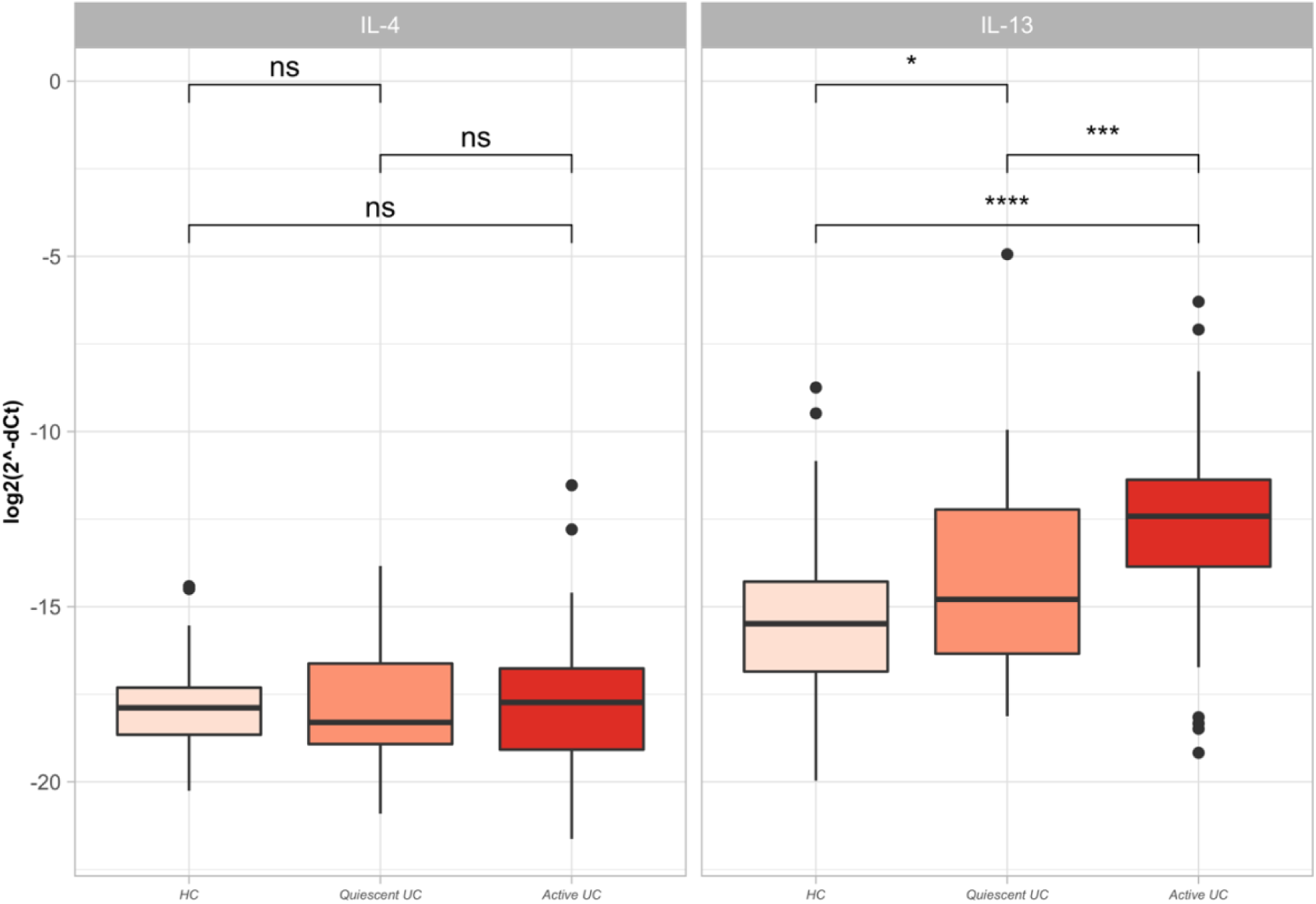
The expression levels of *IL-4* and *IL-13* genes measured by RT-qPCR in colon tissue samples of an independent cohort of HC, active UC, and quiescent UC individuals. Dots represent outliers in each group. Gene expression is represented in logarithmic scale, ΔCt values are inversed in order to show true direction of the expression. Significance levels: * p <= 0.05, *** p <= 0.001, **** p <= 0.0001, ns – not significant.

**Supplementary Figure S3.**
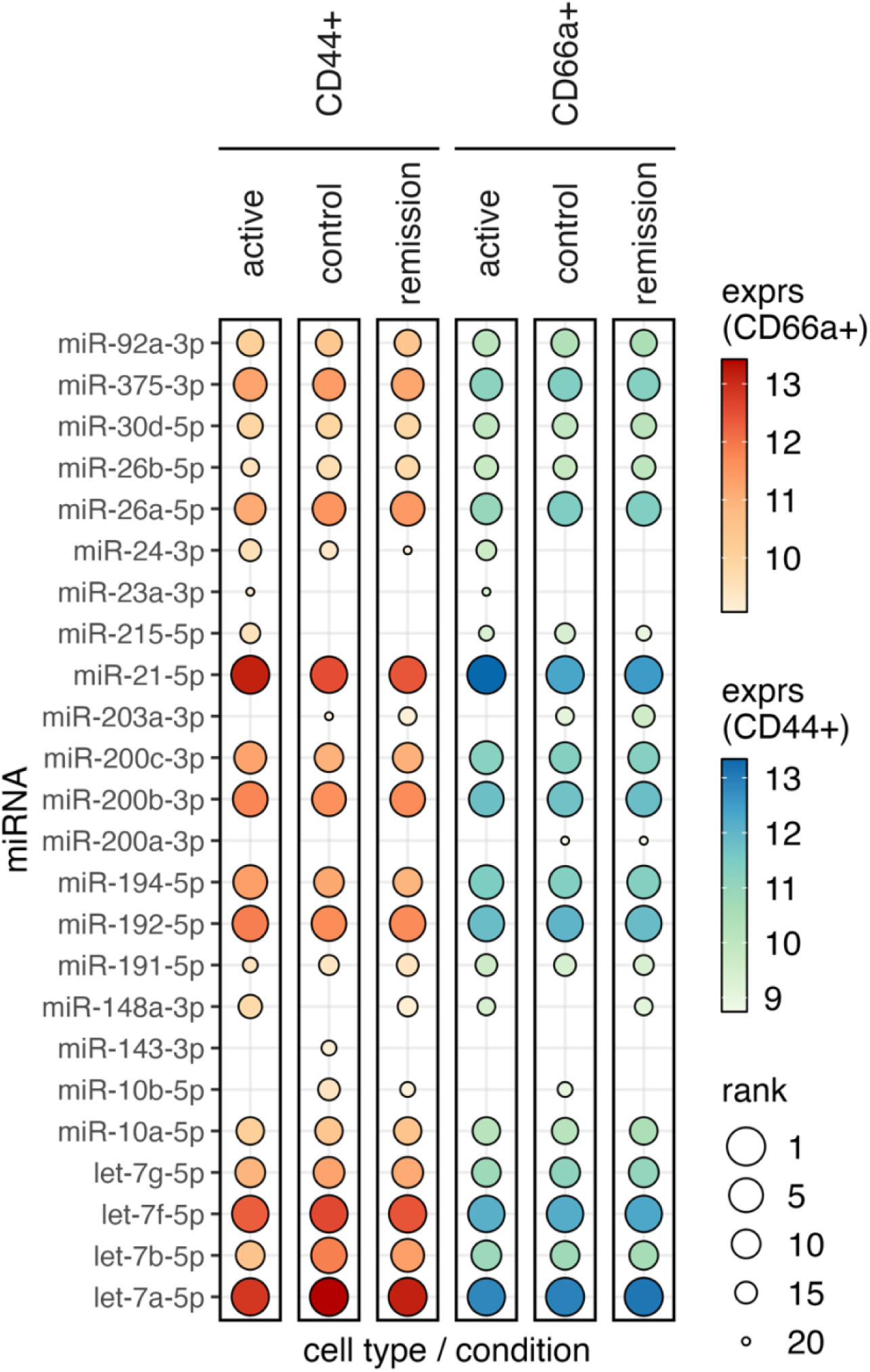
Most abundant (top 20) miRNA expression in colonic epithelial cell CD44+ and CD66a+ populations from UC patients (active and remission) and healthy controls. In each cell population and condition, the median normalized expression levels are represented by color, and rank by dot size. Most abundant miRNAs are ranked in descending order, to be precise, the highest expression value is ranked as 1.

**Supplementary Table S1.** Demographic and clinical characteristics of Study group III subjects recruited for *IL-4* and *IL-13* gene expression analysis in colon tissues.

|  | Study group III, n=200 |  |  |
| --- | --- | --- | --- |
|  | Active<br>UC, n=75 | Quiescent<br>UC, n=50 | HC,<br>n=75 |
| Age |  |  |  |
| Mean ± SD | 46.1 ± 13.7 | 47.3 ± 16.1 | 46.3 ± 11.9 |
| Sex, n (%) |  |  |  |
| Female | 37 (49.3) | 24 (48.0) | 38 (50.7) |
| Full Mayo score, n (%) |  |  |  |
| Remission | 0 (0.0) | 50 (100.0) | - |
| Mild | 3 (4.0) | 0 (0.0) | - |
| Moderate | 70 (93.3) | 0 (0.0) | - |
| Severe | 2 (2.7) | 0 (0.0) | - |
| Endoscopic Mayo subscore, n (%) |  |  |  |
| Normal to mild | 0 (0.0) | 50 (100.0) | 75 (100.0) |
| Moderate to severe | 75 (100.0) | 0 (0.0) | - |
| Smoking status, n (%) |  |  |  |
| Never | 43 (57.3) | 24 (48.0) | 43 (57.3) |
| Smoking | 8 (10.7) | 8 (16.0) | 17 (22.7) |
| Ex-smoker | 22 (29.3) | 16 (32.0) | 12 (16.0) |
| Unknown | 2 (2.7) | 2 (4.0) | 3 (4.0) |
| BMI, kg/m<sup>2</sup> |  |  |  |
| Median, | 24.9 | 25.8 | 24.9 |
| (range) | (16.2–41.5) | (19.5–34.6) | (16.9–49.3) |
| Unknown, n (%) |  | 2 (4.0) | 5 (6.7) |

SD - Standard deviation

**Supplementary Table S2.**
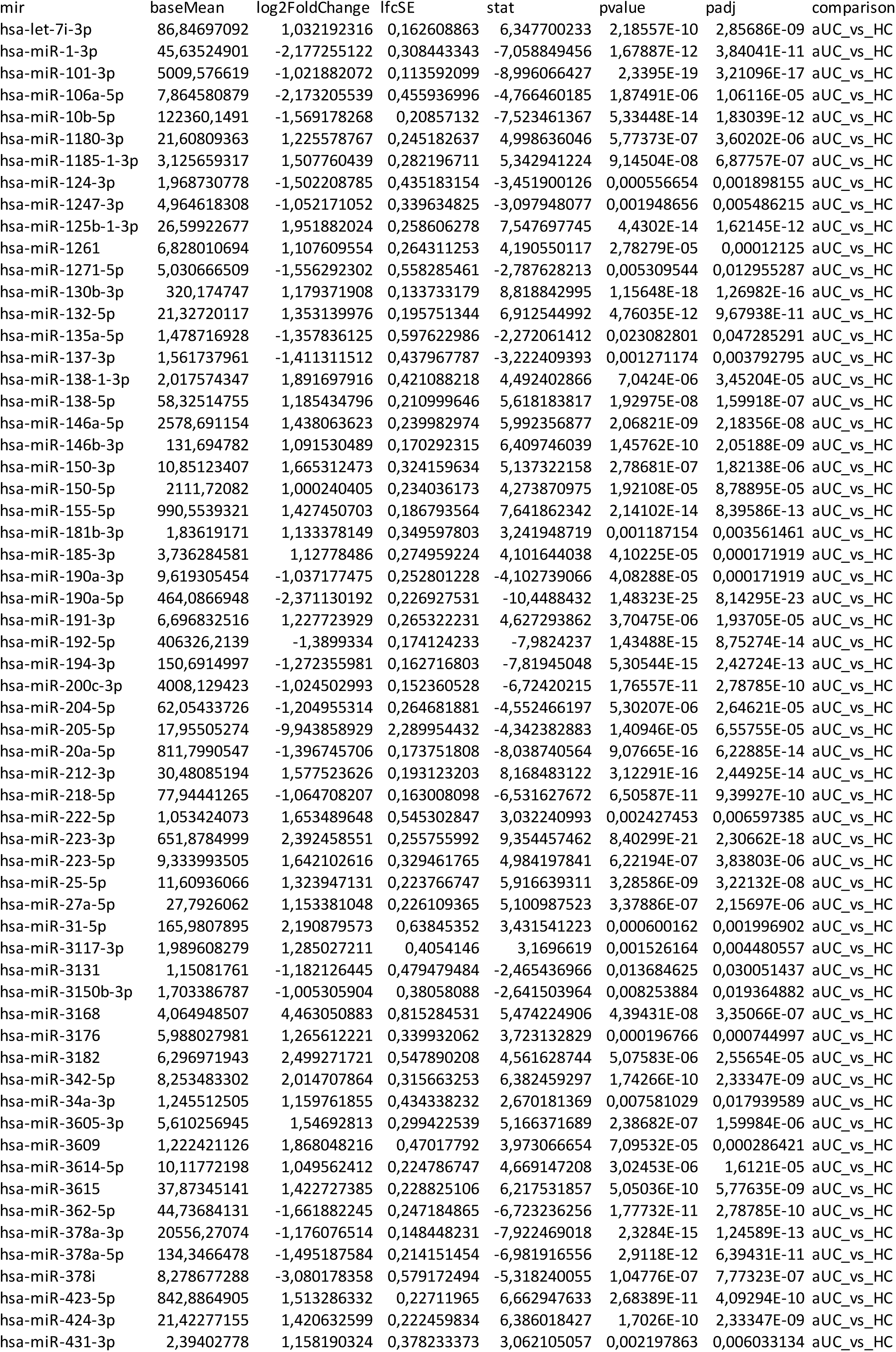
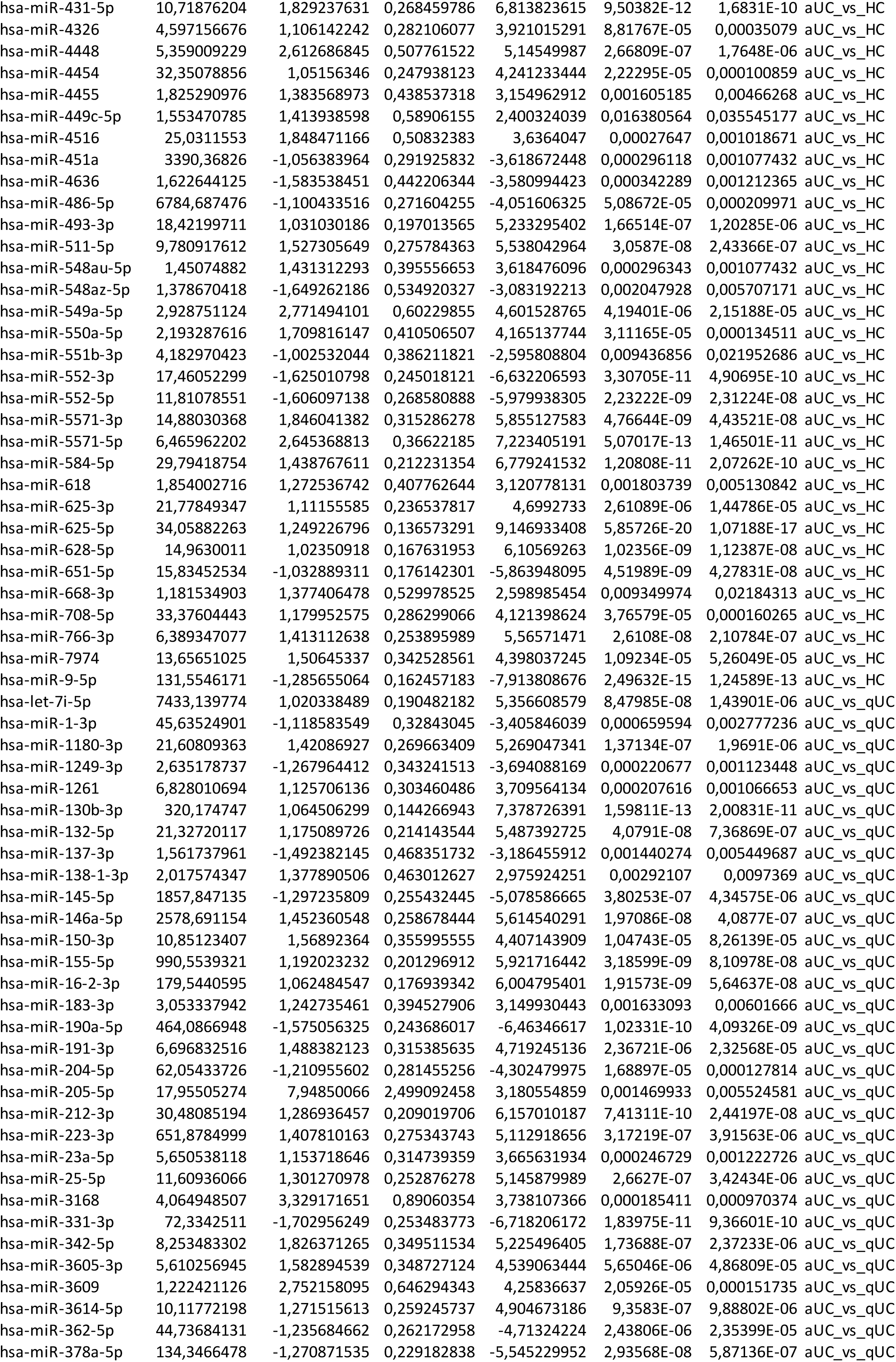
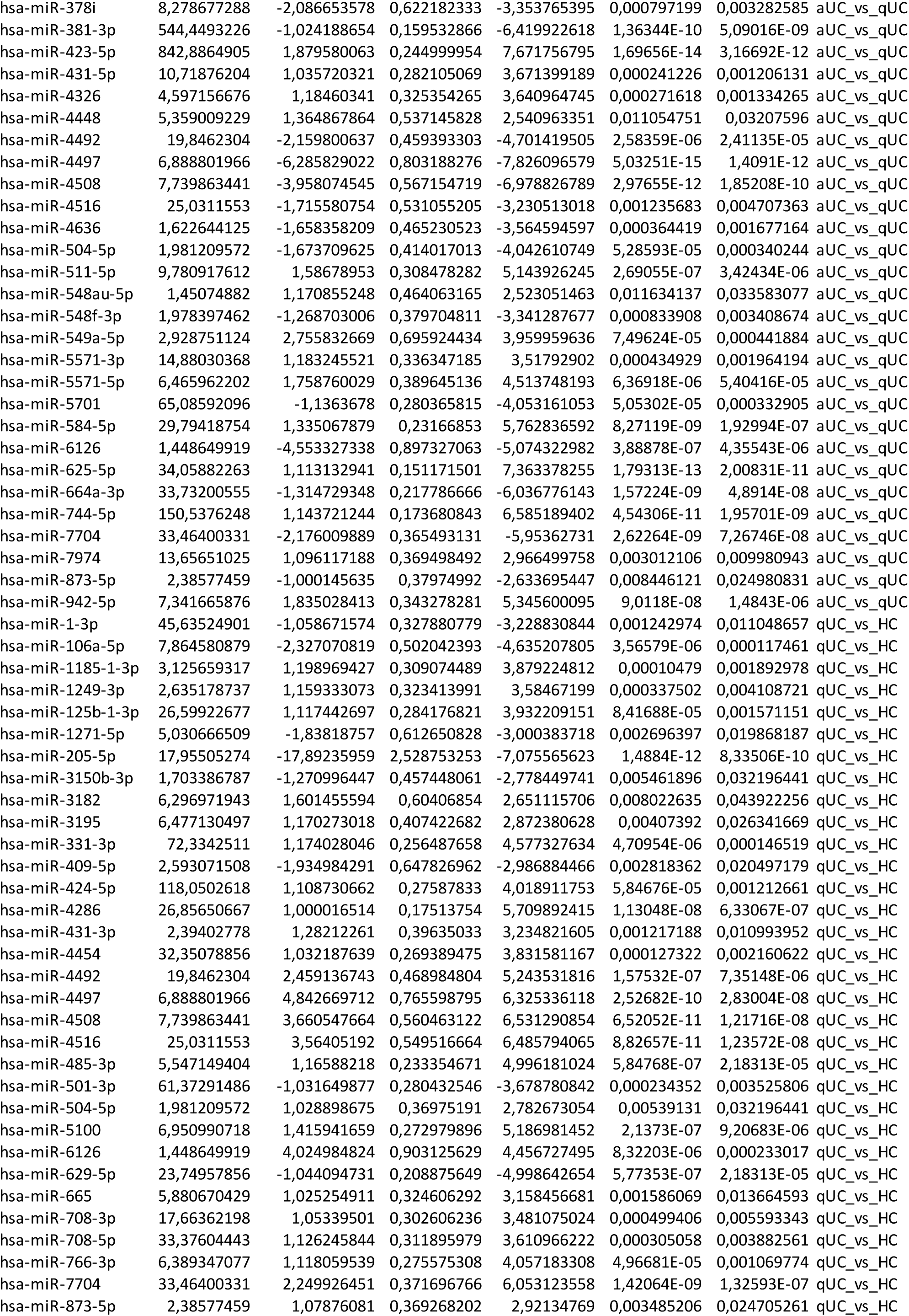
The list of differentially expressed colon tissue miRNAs in active (aUC) and quiescent (qUC) UC.

SD - Standard deviation
|  |  |  |  |  |  |  |  |
| --- | --- | --- | --- | --- | --- | --- | --- |
| mir | baseMean | log2FoldChange | lfcSE | stat | pvalue | padj | comparison |
| hsa-let-7i-3p | 86,84697092 | 1,032192316 | 0,162608863 | 6,347700233 | 2,18557E-10 | 2,85686E-09 | aUC_vs_HC |
| hsa-miR-1-3p | 45,63524901 | -2,177255122 | 0,308443343 | -7,058849456 | 1,67887E-12 | 3,84041E-11 | aUC_vs_HC |
| hsa-miR-101-3p | 5009,576619 | -1,021882072 | 0,113592099 | -8,996066427 | 2,3395E-19 | 3,21096E-17 | aUC_vs_HC |
| hsa-miR-106a-5p | 7,864580879 | -2,173205539 | 0,455936996 | -4,766460185 | 1,87491E-06 | 1,06116E-05 | aUC_vs_HC |
| hsa-miR-10b-5p | 122360,1491 | -1,569178268 | 0,20857132 | -7,523461367 | 5,33448E-14 | 1,83039E-12 | aUC_vs_HC |
| hsa-miR-1180-3p | 21,60809363 | 1,225578767 | 0,245182637 | 4,998636046 | 5,77373E-07 | 3,60202E-06 | aUC_vs_HC |
| hsa-miR-1185-1-3p | 3,125659317 | 1,507760439 | 0,282196711 | 5,342941224 | 9,14504E-08 | 6,87757E-07 | aUC_vs_HC |
| hsa-miR-124-3p | 1,968730778 | -1,502208785 | 0,435183154 | -3,451900126 | 0,000556654 | 0,001898155 | aUC_vs_HC |
| hsa-miR-1247-3p | 4,964618308 | -1,052171052 | 0,339634825 | -3,097948077 | 0,001948656 | 0,005486215 | aUC_vs_HC |
| hsa-miR-125b-1-3p | 26,59922677 | 1,951882024 | 0,258606278 | 7,547697745 | 4,4302E-14 | 1,62145E-12 | aUC_vs_HC |
| hsa-miR-1261 | 6,828010694 | 1,107609554 | 0,264311253 | 4,190550117 | 2,78279E-05 | 0,00012125 | aUC_vs_HC |
| hsa-miR-1271-5p | 5,030666509 | -1,556292302 | 0,558285461 | -2,787628213 | 0,005309544 | 0,012955287 | aUC_vs_HC |
| hsa-miR-130b-3p | 320,174747 | 1,179371908 | 0,133733179 | 8,818842995 | 1,15648E-18 | 1,26982E-16 | aUC_vs_HC |
| hsa-miR-132-5p | 21,32720117 | 1,353139976 | 0,195751344 | 6,912544992 | 4,76035E-12 | 9,67938E-11 | aUC_vs_HC |
| hsa-miR-135a-5p | 1,478716928 | -1,357836125 | 0,597622986 | -2,272061412 | 0,023082801 | 0,047285291 | aUC_vs_HC |
| hsa-miR-137-3p | 1,561737961 | -1,411311512 | 0,437967787 | -3,222409393 | 0,001271174 | 0,003792795 | aUC_vs_HC |
| hsa-miR-138-1-3p | 2,017574347 | 1,891697916 | 0,421088218 | 4,492402866 | 7,0424E-06 | 3,45204E-05 | aUC_vs_HC |
| hsa-miR-138-5p | 58,32514755 | 1,185434796 | 0,210999646 | 5,618183817 | 1,92975E-08 | 1,59918E-07 | aUC_vs_HC |
| hsa-miR-146a-5p | 2578,691154 | 1,438063623 | 0,239982974 | 5,992356877 | 2,06821E-09 | 2,18356E-08 | aUC_vs_HC |
| hsa-miR-146b-3p | 131,694782 | 1,091530489 | 0,170292315 | 6,409746039 | 1,45762E-10 | 2,05188E-09 | aUC_vs_HC |
| hsa-miR-150-3p | 10,85123407 | 1,665312473 | 0,324159634 | 5,137322158 | 2,78681E-07 | 1,82138E-06 | aUC_vs_HC |
| hsa-miR-150-5p | 2111,72082 | 1,000240405 | 0,234036173 | 4,273870975 | 1,92108E-05 | 8,78895E-05 | aUC_vs_HC |
| hsa-miR-155-5p | 990,5539321 | 1,427450703 | 0,186793564 | 7,641862342 | 2,14102E-14 | 8,39586E-13 | aUC_vs_HC |
| hsa-miR-181b-3p | 1,83619171 | 1,133378149 | 0,349597803 | 3,241948719 | 0,001187154 | 0,003561461 | aUC_vs_HC |
| hsa-miR-185-3p | 3,736284581 | 1,12778486 | 0,274959224 | 4,101644038 | 4,10225E-05 | 0,000171919 | aUC_vs_HC |
| hsa-miR-190a-3p | 9,619305454 | -1,037177475 | 0,252801228 | -4,102739066 | 4,08288E-05 | 0,000171919 | aUC_vs_HC |
| hsa-miR-190a-5p | 464,0866948 | -2,371130192 | 0,226927531 | -10,4488432 | 1,48323E-25 | 8,14295E-23 | aUC_vs_HC |
| hsa-miR-191-3p | 6,696832516 | 1,227723929 | 0,265322231 | 4,627293862 | 3,70475E-06 | 1,93705E-05 | aUC_vs_HC |
| hsa-miR-192-5p | 406326,2139 | -1,3899334 | 0,174124233 | -7,9824237 | 1,43488E-15 | 8,75274E-14 | aUC_vs_HC |
| hsa-miR-194-3p | 150,6914997 | -1,272355981 | 0,162716803 | -7,81945048 | 5,30544E-15 | 2,42724E-13 | aUC_vs_HC |
| hsa-miR-200c-3p | 4008,129423 | -1,024502993 | 0,152360528 | -6,72420215 | 1,76557E-11 | 2,78785E-10 | aUC_vs_HC |
| hsa-miR-204-5p | 62,05433726 | -1,204955314 | 0,264681881 | -4,552466197 | 5,30207E-06 | 2,64621E-05 | aUC_vs_HC |
| hsa-miR-205-5p | 17,95505274 | -9,943858929 | 2,289954432 | -4,342382883 | 1,40946E-05 | 6,55755E-05 | aUC_vs_HC |
| hsa-miR-20a-5p | 811,7990547 | -1,396745706 | 0,173751808 | -8,038740564 | 9,07665E-16 | 6,22885E-14 | aUC_vs_HC |
| hsa-miR-212-3p | 30,48085194 | 1,577523626 | 0,193123203 | 8,168483122 | 3,12291E-16 | 2,44925E-14 | aUC_vs_HC |
| hsa-miR-218-5p | 77,94441265 | -1,064708207 | 0,163008098 | -6,531627672 | 6,50587E-11 | 9,39927E-10 | aUC_vs_HC |
| hsa-miR-222-5p | 1,053424073 | 1,653489648 | 0,545302847 | 3,032240993 | 0,002427453 | 0,006597385 | aUC_vs_HC |
| hsa-miR-223-3p | 651,8784999 | 2,392458551 | 0,255755992 | 9,354457462 | 8,40299E-21 | 2,30662E-18 | aUC_vs_HC |
| hsa-miR-223-5p | 9,333993505 | 1,642102616 | 0,329461765 | 4,984197841 | 6,22194E-07 | 3,83803E-06 | aUC_vs_HC |
| hsa-miR-25-5p | 11,60936066 | 1,323947131 | 0,223766747 | 5,916639311 | 3,28586E-09 | 3,22132E-08 | aUC_vs_HC |
| hsa-miR-27a-5p | 27,7926062 | 1,153381048 | 0,226109365 | 5,100987523 | 3,37886E-07 | 2,15697E-06 | aUC_vs_HC |
| hsa-miR-31-5p | 165,9807895 | 2,190879573 | 0,63845352 | 3,431541223 | 0,000600162 | 0,001996902 | aUC_vs_HC |
| hsa-miR-3117-3p | 1,989608279 | 1,285027211 | 0,4054146 | 3,1696619 | 0,001526164 | 0,004480557 | aUC_vs_HC |
| hsa-miR-3131 | 1,15081761 | -1,182126445 | 0,479479484 | -2,465436966 | 0,013684625 | 0,030051437 | aUC_vs_HC |
| hsa-miR-3150b-3p | 1,703386787 | -1,005305904 | 0,38058088 | -2,641503964 | 0,008253884 | 0,019364882 | aUC_vs_HC |
| hsa-miR-3168 | 4,064948507 | 4,463050883 | 0,815284531 | 5,474224906 | 4,39431E-08 | 3,35066E-07 | aUC_vs_HC |
| hsa-miR-3176 | 5,988027981 | 1,265612221 | 0,339932062 | 3,723132829 | 0,000196766 | 0,000744997 | aUC_vs_HC |
| hsa-miR-3182 | 6,296971943 | 2,499271721 | 0,547890208 | 4,561628744 | 5,07583E-06 | 2,55654E-05 | aUC_vs_HC |
| hsa-miR-342-5p | 8,253483302 | 2,014707864 | 0,315663253 | 6,382459297 | 1,74266E-10 | 2,33347E-09 | aUC_vs_HC |
| hsa-miR-34a-3p | 1,245512505 | 1,159761855 | 0,434338232 | 2,670181369 | 0,007581029 | 0,017939589 | aUC_vs_HC |
| hsa-miR-3605-3p | 5,610256945 | 1,54692813 | 0,299422539 | 5,166371689 | 2,38682E-07 | 1,59984E-06 | aUC_vs_HC |
| hsa-miR-3609 | 1,222421126 | 1,868048216 | 0,47017792 | 3,973066654 | 7,09532E-05 | 0,000286421 | aUC_vs_HC |
| hsa-miR-3614-5p | 10,11772198 | 1,049562412 | 0,224786747 | 4,669147208 | 3,02453E-06 | 1,6121E-05 | aUC_vs_HC |
| hsa-miR-3615 | 37,87345141 | 1,422727385 | 0,228825106 | 6,217531857 | 5,05036E-10 | 5,77635E-09 | aUC_vs_HC |
| hsa-miR-362-5p | 44,73684131 | -1,661882245 | 0,247184865 | -6,723236256 | 1,77732E-11 | 2,78785E-10 | aUC_vs_HC |
| hsa-miR-378a-3p | 20556,27074 | -1,176076514 | 0,148448231 | -7,922469018 | 2,3284E-15 | 1,24589E-13 | aUC_vs_HC |
| hsa-miR-378a-5p | 134,3466478 | -1,495187584 | 0,214151454 | -6,981916556 | 2,9118E-12 | 6,39431E-11 | aUC_vs_HC |
| hsa-miR-378i | 8,278677288 | -3,080178358 | 0,579172494 | -5,318240055 | 1,04776E-07 | 7,77323E-07 | aUC_vs_HC |
| hsa-miR-423-5p | 842,8864905 | 1,513286332 | 0,22711965 | 6,662947633 | 2,68389E-11 | 4,09294E-10 | aUC_vs_HC |
| hsa-miR-424-3p | 21,42277155 | 1,420632599 | 0,222459834 | 6,386018427 | 1,7026E-10 | 2,33347E-09 | aUC_vs_HC |
| hsa-miR-431-3p | 2,39402778 | 1,158190324 | 0,378233373 | 3,062105057 | 0,002197863 | 0,006033134 | aUC_vs_HC |

|  |  |  |  |  |  |  |  |  |
| --- | --- | --- | --- | --- | --- | --- | --- | --- |
| 1 |  |  |  |  |  |  |  |  |
| 2 | hsa-miR-431-5p | 10,71876204 | 1,829237631 | 0,268459786 | 6,813823615 | 9,50382E-12 | 1,6831E-10 | aUC_vs_HC |
| 3 | hsa-miR-4326 | 4,597156676 | 1,106142242 | 0,282106077 | 3,921015291 | 8,81767E-05 | 0,00035079 | aUC_vs_HC |
| 4 | hsa-miR-4448 | 5,359009229 | 2,612686845 | 0,507761522 | 5,14549987 | 2,66809E-07 | 1,7648E-06 | aUC_vs_HC |
| 5 | hsa-miR-4454 | 32,35078856 | 1,05156346 | 0,247938123 | 4,241233444 | 2,22295E-05 | 0,000100859 | aUC_vs_HC |
| 6 | hsa-miR-4455 | 1,825290976 | 1,383568973 | 0,438537318 | 3,154962912 | 0,001605185 | 0,00466268 | aUC_vs_HC |
| 7 | hsa-miR-449c-5p | 1,553470785 | 1,413938598 | 0,58906155 | 2,400324039 | 0,016380564 | 0,035545177 | aUC_vs_HC |
| 8 | hsa-miR-4516 | 25,0311553 | 1,848471166 | 0,50832383 | 3,6364047 | 0,00027647 | 0,001018671 | aUC_vs_HC |
| 9 | hsa-miR-451a | 3390,36826 | -1,056383964 | 0,291925832 | -3,618672448 | 0,000296118 | 0,001077432 | aUC_vs_HC |
| 10 | hsa-miR-4636 | 1,622644125 | -1,583538451 | 0,442206344 | -3,580994423 | 0,000342289 | 0,001212365 | aUC_vs_HC |
| 11 | hsa-miR-486-5p | 6784,687476 | -1,100433516 | 0,271604255 | -4,051606325 | 5,08672E-05 | 0,000209971 | aUC_vs_HC |
| 12 | hsa-miR-493-3p | 18,42199711 | 1,031030186 | 0,197013565 | 5,233295402 | 1,66514E-07 | 1,20285E-06 | aUC_vs_HC |
| 13 | hsa-miR-511-5p | 9,780917612 | 1,527305649 | 0,275784363 | 5,538042964 | 3,0587E-08 | 2,43366E-07 | aUC_vs_HC |
| 14 | hsa-miR-548au-5p | 1,45074882 | 1,431312293 | 0,395556653 | 3,618476096 | 0,000296343 | 0,001077432 | aUC_vs_HC |
| 15 | hsa-miR-548az-5p | 1,378670418 | -1,649262186 | 0,534920327 | -3,083192213 | 0,002047928 | 0,005707171 | aUC_vs_HC |
| 16 | hsa-miR-549a-5p | 2,928751124 | 2,771494101 | 0,60229855 | 4,601528765 | 4,19401E-06 | 2,15188E-05 | aUC_vs_HC |
| 17 | hsa-miR-550a-5p | 2,193287616 | 1,709816147 | 0,410506507 | 4,165137744 | 3,11165E-05 | 0,000134511 | aUC_vs_HC |
| 18 | hsa-miR-551b-3p | 4,182970423 | -1,002532044 | 0,386211821 | -2,595808804 | 0,009436856 | 0,021952686 | aUC_vs_HC |
| 19 | hsa-miR-552-3p | 17,46052299 | -1,625010798 | 0,245018121 | -6,632206593 | 3,30705E-11 | 4,90695E-10 | aUC_vs_HC |
| 20 | hsa-miR-552-5p | 11,81078551 | -1,606097138 | 0,268580888 | -5,979938305 | 2,23222E-09 | 2,31224E-08 | aUC_vs_HC |
| 21 | hsa-miR-5571-3p | 14,88030368 | 1,846041382 | 0,315286278 | 5,855127583 | 4,76644E-09 | 4,43521E-08 | aUC_vs_HC |
| 22 | hsa-miR-5571-5p | 6,465962202 | 2,645368813 | 0,36622185 | 7,223405191 | 5,07017E-13 | 1,46501E-11 | aUC_vs_HC |
| 23 | hsa-miR-584-5p | 29,79418754 | 1,438767611 | 0,212231354 | 6,779241532 | 1,20808E-11 | 2,07262E-10 | aUC_vs_HC |
| 24 | hsa-miR-618 | 1,854002716 | 1,272536742 | 0,407762644 | 3,120778131 | 0,001803739 | 0,005130842 | aUC_vs_HC |
| 25 | hsa-miR-625-3p | 21,77849347 | 1,11155585 | 0,236537817 | 4,6992733 | 2,61089E-06 | 1,44786E-05 | aUC_vs_HC |
| 26 | hsa-miR-625-5p | 34,05882263 | 1,249226796 | 0,136573291 | 9,146933408 | 5,85726E-20 | 1,07188E-17 | aUC_vs_HC |
| 27 | hsa-miR-628-5p | 14,9630011 | 1,02350918 | 0,167631953 | 6,10569263 | 1,02356E-09 | 1,12387E-08 | aUC_vs_HC |
| 28 | hsa-miR-651-5p | 15,83452534 | -1,032889311 | 0,176142301 | -5,863948095 | 4,51989E-09 | 4,27831E-08 | aUC_vs_HC |
| 29 | hsa-miR-668-3p | 1,181534903 | 1,377406478 | 0,529978525 | 2,598985454 | 0,009349974 | 0,02184313 | aUC_vs_HC |
| 30 | hsa-miR-708-5p | 33,37604443 | 1,179952575 | 0,286299066 | 4,121398624 | 3,76579E-05 | 0,000160265 | aUC_vs_HC |
| 31 | hsa-miR-766-3p | 6,389347077 | 1,413112638 | 0,253895989 | 5,56571471 | 2,6108E-08 | 2,10784E-07 | aUC_vs_HC |
| 32 | hsa-miR-7974 | 13,65651025 | 1,50645337 | 0,342528561 | 4,398037245 | 1,09234E-05 | 5,26049E-05 | aUC_vs_HC |
| 33 | hsa-miR-9-5p | 131,5546171 | -1,285655064 | 0,162457183 | -7,913808676 | 2,49632E-15 | 1,24589E-13 | aUC_vs_HC |
| 34 | hsa-let-7i-5p | 7433,139774 | 1,020338489 | 0,190482182 | 5,356608579 | 8,47985E-08 | 1,43901E-06 | aUC_vs_qUC |
| 35 | hsa-miR-1-3p | 45,63524901 | -1,118583549 | 0,32843045 | -3,405846039 | 0,000659594 | 0,002777236 | aUC_vs_qUC |
| 36 | hsa-miR-1180-3p | 21,60809363 | 1,42086927 | 0,269663409 | 5,269047341 | 1,37134E-07 | 1,9691E-06 | aUC_vs_qUC |
| 37 | hsa-miR-1249-3p | 2,635178737 | -1,267964412 | 0,343241513 | -3,694088169 | 0,000220677 | 0,001123448 | aUC_vs_qUC |
| 38 | hsa-miR-1261 | 6,828010694 | 1,125706136 | 0,303460486 | 3,709564134 | 0,000207616 | 0,001066653 | aUC_vs_qUC |
| 39 | hsa-miR-130b-3p | 320,174747 | 1,064506299 | 0,144266943 | 7,378726391 | 1,59811E-13 | 2,00831E-11 | aUC_vs_qUC |
| 40 | hsa-miR-132-5p | 21,32720117 | 1,175089726 | 0,214143544 | 5,487392725 | 4,0791E-08 | 7,36869E-07 | aUC_vs_qUC |
| 41 | hsa-miR-137-3p | 1,561737961 | -1,492382145 | 0,468351732 | -3,186455912 | 0,001440274 | 0,005449687 | aUC_vs_qUC |
| 42 | hsa-miR-138-1-3p | 2,017574347 | 1,377890506 | 0,463012627 | 2,975924251 | 0,00292107 | 0,0097369 | aUC_vs_qUC |
| 43 | hsa-miR-145-5p | 1857,847135 | -1,297235809 | 0,255432445 | -5,078586665 | 3,80253E-07 | 4,34575E-06 | aUC_vs_qUC |
| 44 | hsa-miR-146a-5p | 2578,691154 | 1,452360548 | 0,258678444 | 5,614540291 | 1,97086E-08 | 4,0877E-07 | aUC_vs_qUC |
| 45 | hsa-miR-150-3p | 10,85123407 | 1,56892364 | 0,355995555 | 4,407143909 | 1,04743E-05 | 8,26139E-05 | aUC_vs_qUC |
| 46 | hsa-miR-155-5p | 990,5539321 | 1,192023232 | 0,201296912 | 5,921716442 | 3,18599E-09 | 8,10978E-08 | aUC_vs_qUC |
| 47 | hsa-miR-16-2-3p | 179,5440595 | 1,062484547 | 0,176939342 | 6,004795401 | 1,91573E-09 | 5,64637E-08 | aUC_vs_qUC |
| 48 | hsa-miR-183-3p | 3,053337942 | 1,242735461 | 0,394527906 | 3,149930443 | 0,001633093 | 0,00601666 | aUC_vs_qUC |
| 49 | hsa-miR-190a-5p | 464,0866948 | -1,575056325 | 0,243686017 | -6,46346617 | 1,02331E-10 | 4,09326E-09 | aUC_vs_qUC |
| 50 | hsa-miR-191-3p | 6,696832516 | 1,488382123 | 0,315385635 | 4,719245136 | 2,36721E-06 | 2,32568E-05 | aUC_vs_qUC |
| 51 | hsa-miR-204-5p | 62,05433726 | -1,210955602 | 0,281455256 | -4,302479975 | 1,68897E-05 | 0,000127814 | aUC_vs_qUC |
| 52 | hsa-miR-205-5p | 17,95505274 | 7,94850066 | 2,499092458 | 3,180554859 | 0,001469933 | 0,005524581 | aUC_vs_qUC |
| 53 | hsa-miR-212-3p | 30,48085194 | 1,286936457 | 0,209019706 | 6,157010187 | 7,41311E-10 | 2,44197E-08 | aUC_vs_qUC |
| 54 | hsa-miR-223-3p | 651,8784999 | 1,407810163 | 0,275343743 | 5,112918656 | 3,17219E-07 | 3,91563E-06 | aUC_vs_qUC |
| 55 | hsa-miR-23a-5p | 5,650538118 | 1,153718646 | 0,314739359 | 3,665631934 | 0,000246729 | 0,001222726 | aUC_vs_qUC |
| 56 | hsa-miR-25-5p | 11,60936066 | 1,301270978 | 0,252876278 | 5,145879989 | 2,6627E-07 | 3,42434E-06 | aUC_vs_qUC |
| 57 | hsa-miR-3168 | 4,064948507 | 3,329171651 | 0,89060354 | 3,738107366 | 0,000185411 | 0,000970374 | aUC_vs_qUC |
| 58 | hsa-miR-331-3p | 72,3342511 | -1,702956249 | 0,253483773 | -6,718206172 | 1,83975E-11 | 9,36601E-10 | aUC_vs_qUC |
| 59 | hsa-miR-342-5p | 8,253483302 | 1,826371265 | 0,349511534 | 5,225496405 | 1,73688E-07 | 2,37233E-06 | aUC_vs_qUC |
| 60 | hsa-miR-3605-3p | 5,610256945 | 1,582894539 | 0,348727124 | 4,539063444 | 5,65046E-06 | 4,86809E-05 | aUC_vs_qUC |
|  | hsa-miR-3609 | 1,222421126 | 2,752158095 | 0,646294343 | 4,25836637 | 2,05926E-05 | 0,000151735 | aUC_vs_qUC |
|  | hsa-miR-3614-5p | 10,11772198 | 1,271515613 | 0,259245737 | 4,904673186 | 9,3583E-07 | 9,88802E-06 | aUC_vs_qUC |
|  | hsa-miR-362-5p | 44,73684131 | -1,235684662 | 0,262172958 | -4,71324224 | 2,43806E-06 | 2,35399E-05 | aUC_vs_qUC |
|  | hsa-miR-378a-5p | 134,3466478 | -1,270871535 | 0,229182838 | -5,545229952 | 2,93568E-08 | 5,87136E-07 | aUC_vs_qUC |

|  |  |  |  |  |  |  |  |  |
| --- | --- | --- | --- | --- | --- | --- | --- | --- |
| 2 | hsa-miR-378i | 8,278677288 | -2,086653578 | 0,622182333 | -3,353765395 | 0,000797199 | 0,003282585 | aUC_vs_qUC |
| 3 | hsa-miR-381-3p | 544,4493226 | -1,024188654 | 0,159532866 | -6,419922618 | 1,36344E-10 | 5,09016E-09 | aUC_vs_qUC |
| 4 | hsa-miR-423-5p | 842,8864905 | 1,879580063 | 0,244999954 | 7,671756795 | 1,69656E-14 | 3,16692E-12 | aUC_vs_qUC |
| 5 | hsa-miR-431-5p | 10,71876204 | 1,035720321 | 0,282105069 | 3,671399189 | 0,000241226 | 0,001206131 | aUC_vs_qUC |
| 6 | hsa-miR-4326 | 4,597156676 | 1,18460341 | 0,325354265 | 3,640964745 | 0,000271618 | 0,001334265 | aUC_vs_qUC |
| 7 | hsa-miR-4448 | 5,359009229 | 1,364867864 | 0,537145828 | 2,540963351 | 0,011054751 | 0,03207596 | aUC_vs_qUC |
| 8 | hsa-miR-4492 | 19,8462304 | -2,159800637 | 0,459393303 | -4,701419505 | 2,58359E-06 | 2,41135E-05 | aUC_vs_qUC |
| 9 | hsa-miR-4497 | 6,888801966 | -6,285829022 | 0,803188276 | -7,826096579 | 5,03251E-15 | 1,4091E-12 | aUC_vs_qUC |
| 10 | hsa-miR-4508 | 7,739863441 | -3,958074545 | 0,567154719 | -6,978826789 | 2,97655E-12 | 1,85208E-10 | aUC_vs_qUC |
| 11 | hsa-miR-4516 | 25,0311553 | -1,715580754 | 0,531055205 | -3,230513018 | 0,001235683 | 0,004707363 | aUC_vs_qUC |
| 12 | hsa-miR-4636 | 1,622644125 | -1,658358209 | 0,465230523 | -3,564594597 | 0,000364419 | 0,001677164 | aUC_vs_qUC |
| 13 | hsa-miR-504-5p | 1,981209572 | -1,673709625 | 0,414017013 | -4,042610749 | 5,28593E-05 | 0,000340244 | aUC_vs_qUC |
| 14 | hsa-miR-511-5p | 9,780917612 | 1,58678953 | 0,308478282 | 5,143926245 | 2,69055E-07 | 3,42434E-06 | aUC_vs_qUC |
| 15 | hsa-miR-548au-5p | 1,45074882 | 1,170855248 | 0,464063165 | 2,523051463 | 0,011634137 | 0,033583077 | aUC_vs_qUC |
| 16 | hsa-miR-548f-3p | 1,978397462 | -1,268703006 | 0,379704811 | -3,341287677 | 0,000833908 | 0,003408674 | aUC_vs_qUC |
| 17 | hsa-miR-549a-5p | 2,928751124 | 2,755832669 | 0,695924434 | 3,959959636 | 7,49624E-05 | 0,000441884 | aUC_vs_qUC |
| 18 | hsa-miR-5571-3p | 14,88030368 | 1,183245521 | 0,336347185 | 3,51792902 | 0,000434929 | 0,001964194 | aUC_vs_qUC |
| 19 | hsa-miR-5571-5p | 6,465962202 | 1,758760029 | 0,389645136 | 4,513748193 | 6,36918E-06 | 5,40416E-05 | aUC_vs_qUC |
| 20 | hsa-miR-5701 | 65,08592096 | -1,1363678 | 0,280365815 | -4,053161053 | 5,05302E-05 | 0,000332905 | aUC_vs_qUC |
| 21 | hsa-miR-584-5p | 29,79418754 | 1,335067879 | 0,23166853 | 5,762836592 | 8,27119E-09 | 1,92994E-07 | aUC_vs_qUC |
| 22 | hsa-miR-6126 | 1,448649919 | -4,553327338 | 0,897327063 | -5,074322982 | 3,88878E-07 | 4,35543E-06 | aUC_vs_qUC |
| 23 | hsa-miR-625-5p | 34,05882263 | 1,113132941 | 0,151171501 | 7,363378255 | 1,79313E-13 | 2,00831E-11 | aUC_vs_qUC |
| 24 | hsa-miR-664a-3p | 33,73200555 | -1,314729348 | 0,217786666 | -6,036776143 | 1,57224E-09 | 4,8914E-08 | aUC_vs_qUC |
| 25 | hsa-miR-744-5p | 150,5376248 | 1,143721244 | 0,173680843 | 6,585189402 | 4,54306E-11 | 1,95701E-09 | aUC_vs_qUC |
| 26 | hsa-miR-7704 | 33,46400331 | -2,176009889 | 0,365493131 | -5,95362731 | 2,62264E-09 | 7,26746E-08 | aUC_vs_qUC |
| 27 | hsa-miR-7974 | 13,65651025 | 1,096117188 | 0,369498492 | 2,966499758 | 0,003012106 | 0,009980943 | aUC_vs_qUC |
| 28 | hsa-miR-873-5p | 2,38577459 | -1,000145635 | 0,37974992 | -2,633695447 | 0,008446121 | 0,024980831 | aUC_vs_qUC |
| 29 | hsa-miR-942-5p | 7,341665876 | 1,835028413 | 0,343278281 | 5,345600095 | 9,0118E-08 | 1,4843E-06 | aUC_vs_qUC |
| 30 | hsa-miR-1-3p | 45,63524901 | -1,058671574 | 0,327880779 | -3,228830844 | 0,001242974 | 0,011048657 | qUC_vs_HC |
| 31 | hsa-miR-106a-5p | 7,864580879 | -2,327070819 | 0,502042393 | -4,635207805 | 3,56579E-06 | 0,000117461 | qUC_vs_HC |
| 32 | hsa-miR-1185-1-3p | 3,125659317 | 1,198969427 | 0,309074489 | 3,879224812 | 0,00010479 | 0,001892978 | qUC_vs_HC |
| 33 | hsa-miR-1249-3p | 2,635178737 | 1,159333073 | 0,323413991 | 3,58467199 | 0,000337502 | 0,004108721 | qUC_vs_HC |
| 34 | hsa-miR-125b-1-3p | 26,59922677 | 1,117442697 | 0,284176821 | 3,932209151 | 8,41688E-05 | 0,001571151 | qUC_vs_HC |
| 35 | hsa-miR-1271-5p | 5,030666509 | -1,83818757 | 0,612650828 | -3,000383718 | 0,002696397 | 0,019868187 | qUC_vs_HC |
| 36 | hsa-miR-205-5p | 17,95505274 | -17,89235959 | 2,528753253 | -7,075565623 | 1,4884E-12 | 8,33506E-10 | qUC_vs_HC |
| 37 | hsa-miR-3150b-3p | 1,703386787 | -1,270996447 | 0,457448061 | -2,778449741 | 0,005461896 | 0,032196441 | qUC_vs_HC |
| 38 | hsa-miR-3182 | 6,296971943 | 1,601455594 | 0,60406854 | 2,651115706 | 0,008022635 | 0,043922256 | qUC_vs_HC |
| 39 | hsa-miR-3195 | 6,477130497 | 1,170273018 | 0,407422682 | 2,872380628 | 0,00407392 | 0,026341669 | qUC_vs_HC |
| 40 | hsa-miR-331-3p | 72,3342511 | 1,174028046 | 0,256487658 | 4,577327634 | 4,70954E-06 | 0,000146519 | qUC_vs_HC |
| 41 | hsa-miR-409-5p | 2,593071508 | -1,934984291 | 0,647826962 | -2,986884466 | 0,002818362 | 0,020497179 | qUC_vs_HC |
| 42 | hsa-miR-424-5p | 118,0502618 | 1,108730662 | 0,27587833 | 4,018911753 | 5,84676E-05 | 0,001212661 | qUC_vs_HC |
| 43 | hsa-miR-4286 | 26,85650667 | 1,000016514 | 0,17513754 | 5,709892415 | 1,13048E-08 | 6,33067E-07 | qUC_vs_HC |
| 44 | hsa-miR-431-3p | 2,39402778 | 1,28212261 | 0,39635033 | 3,234821605 | 0,001217188 | 0,010993952 | qUC_vs_HC |
| 45 | hsa-miR-4454 | 32,35078856 | 1,032187639 | 0,269389475 | 3,831581167 | 0,000127322 | 0,002160622 | qUC_vs_HC |
| 46 | hsa-miR-4492 | 19,8462304 | 2,459136743 | 0,468984804 | 5,243531816 | 1,57532E-07 | 7,35148E-06 | qUC_vs_HC |
| 47 | hsa-miR-4497 | 6,888801966 | 4,842669712 | 0,765598795 | 6,325336118 | 2,52682E-10 | 2,83004E-08 | qUC_vs_HC |
| 48 | hsa-miR-4508 | 7,739863441 | 3,660547664 | 0,560463122 | 6,531290854 | 6,52052E-11 | 1,21716E-08 | qUC_vs_HC |
| 49 | hsa-miR-4516 | 25,0311553 | 3,56405192 | 0,549516664 | 6,485794065 | 8,82657E-11 | 1,23572E-08 | qUC_vs_HC |
| 50 | hsa-miR-485-3p | 5,547149404 | 1,16588218 | 0,233354671 | 4,996181024 | 5,84768E-07 | 2,18313E-05 | qUC_vs_HC |
| 51 | hsa-miR-501-3p | 61,37291486 | -1,031649877 | 0,280432546 | -3,678780842 | 0,000234352 | 0,003525806 | qUC_vs_HC |
| 52 | hsa-miR-504-5p | 1,981209572 | 1,028898675 | 0,36975191 | 2,782673054 | 0,00539131 | 0,032196441 | qUC_vs_HC |
| 53 | hsa-miR-5100 | 6,950990718 | 1,415941659 | 0,272979896 | 5,186981452 | 2,1373E-07 | 9,20683E-06 | qUC_vs_HC |
| 54 | hsa-miR-6126 | 1,448649919 | 4,024984824 | 0,903125629 | 4,456727495 | 8,32203E-06 | 0,000233017 | qUC_vs_HC |
| 55 | hsa-miR-629-5p | 23,74957856 | -1,044094731 | 0,208875649 | -4,998642654 | 5,77353E-07 | 2,18313E-05 | qUC_vs_HC |
| 56 | hsa-miR-665 | 5,880670429 | 1,025254911 | 0,324606292 | 3,158456681 | 0,001586069 | 0,013664593 | qUC_vs_HC |
| 57 | hsa-miR-708-3p | 17,66362198 | 1,05339501 | 0,302606236 | 3,481075024 | 0,000499406 | 0,005593343 | qUC_vs_HC |
| 58 | hsa-miR-708-5p | 33,37604443 | 1,126245844 | 0,311895979 | 3,610966222 | 0,000305058 | 0,003882561 | qUC_vs_HC |
| 59 | hsa-miR-766-3p | 6,389347077 | 1,118059539 | 0,275575308 | 4,057183308 | 4,96681E-05 | 0,001069774 | qUC_vs_HC |
| 60 | hsa-miR-7704 | 33,46400331 | 2,249926451 | 0,371696766 | 6,053123558 | 1,42064E-09 | 1,32593E-07 | qUC_vs_HC |
| 61 | hsa-miR-873-5p | 2,38577459 | 1,07876081 | 0,369268202 | 2,92134769 | 0,003485206 | 0,024705261 | qUC_vs_HC |

**Supplemenatry Table S3.**
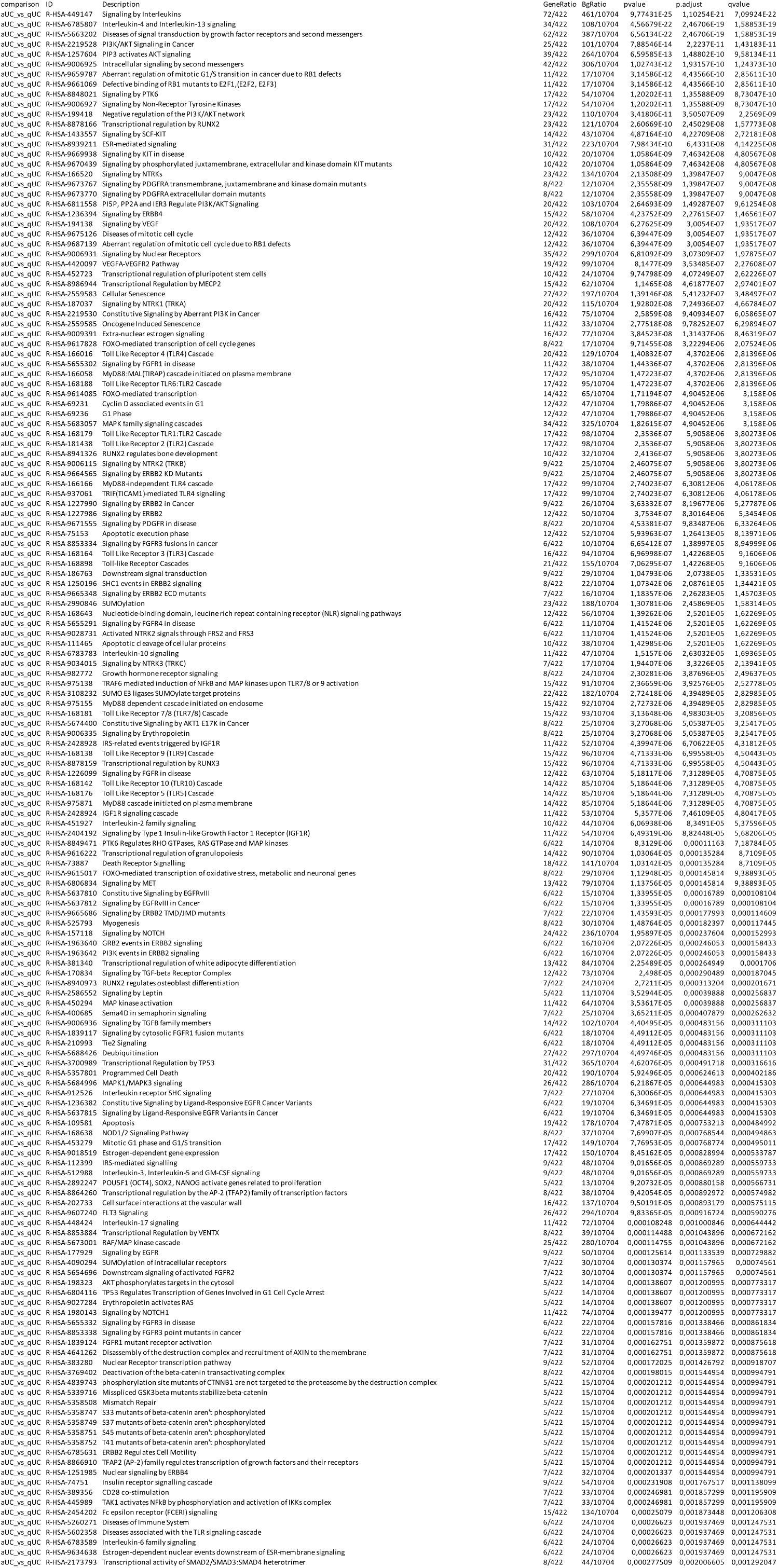
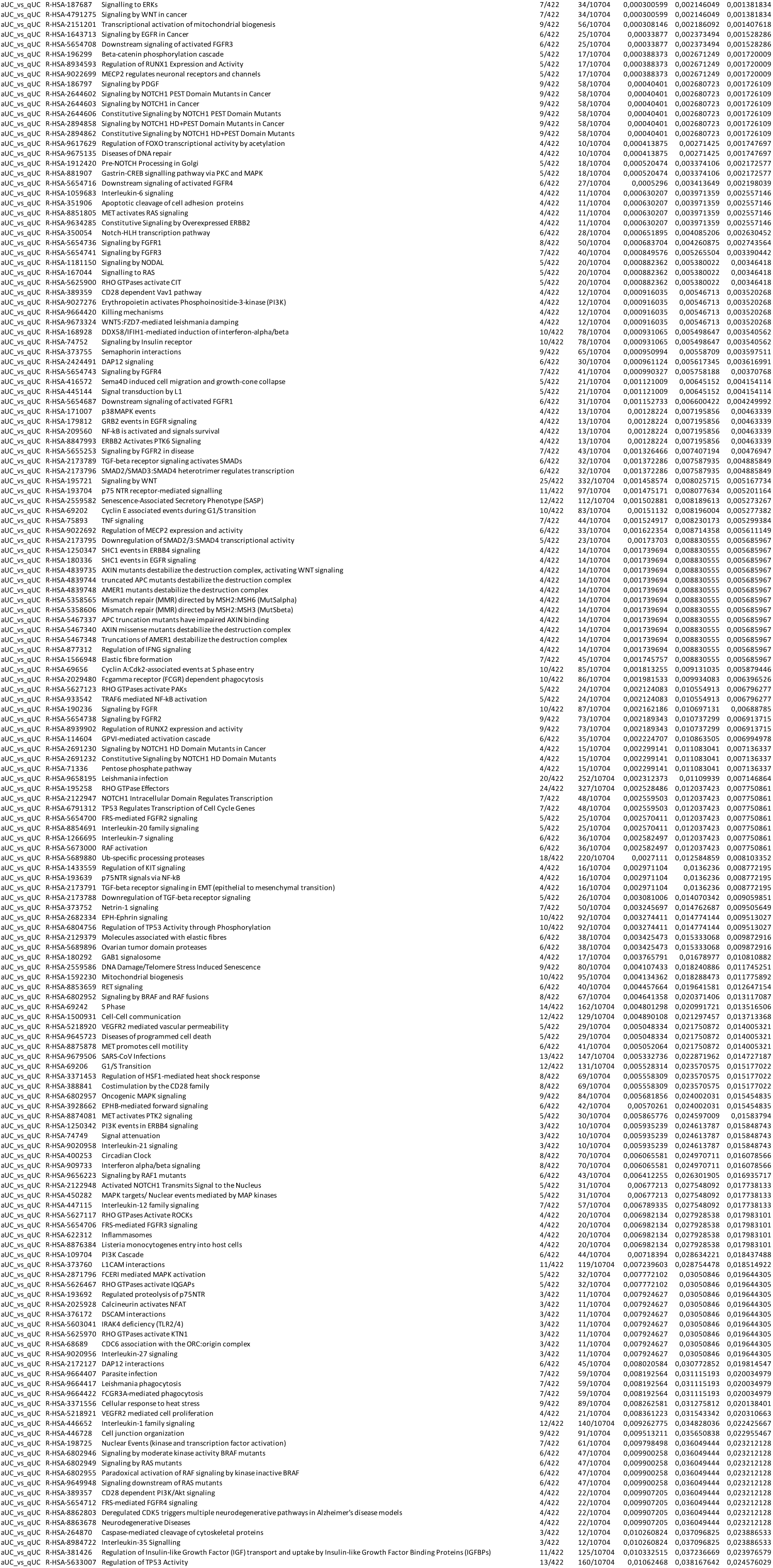
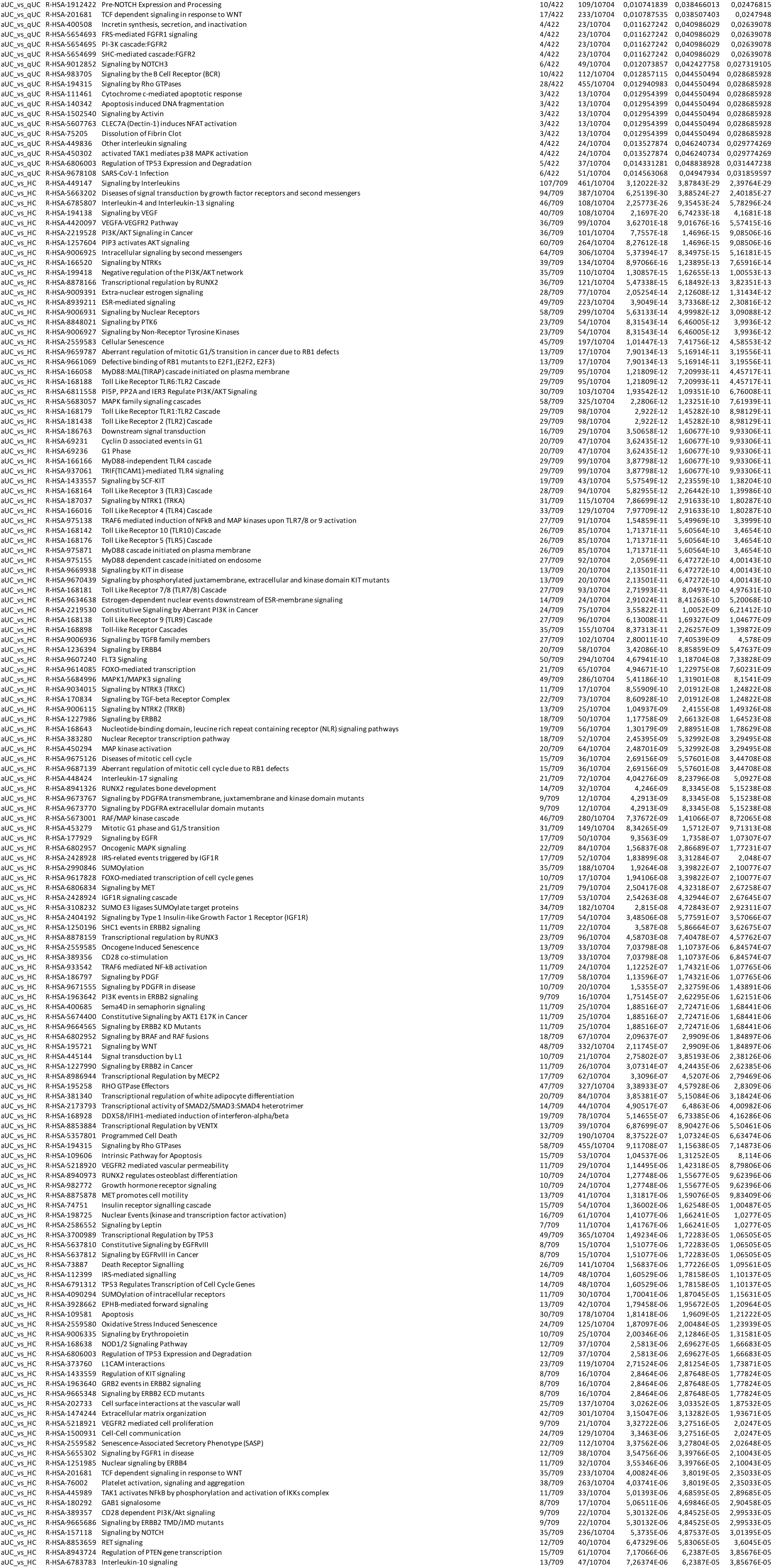
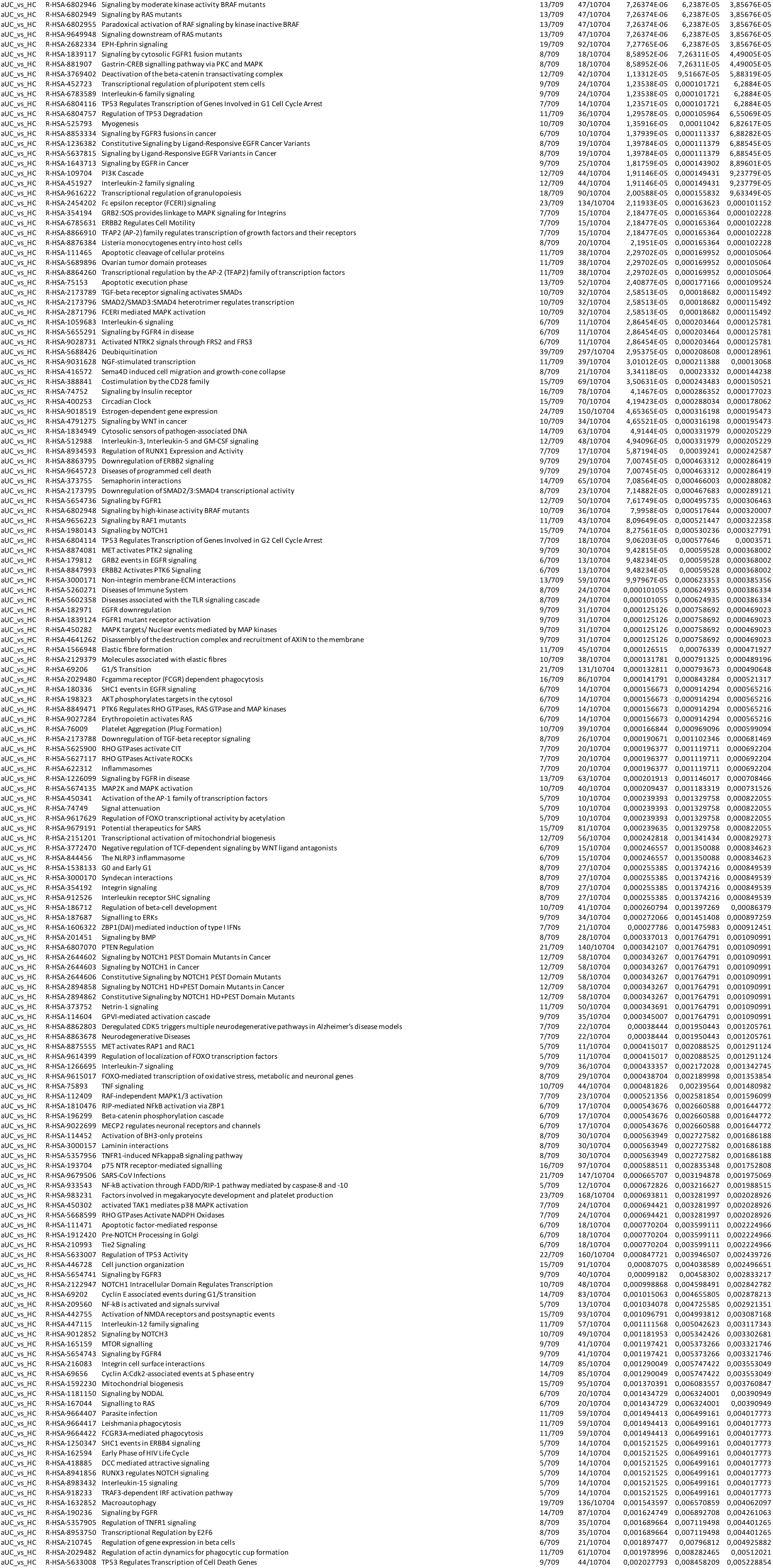
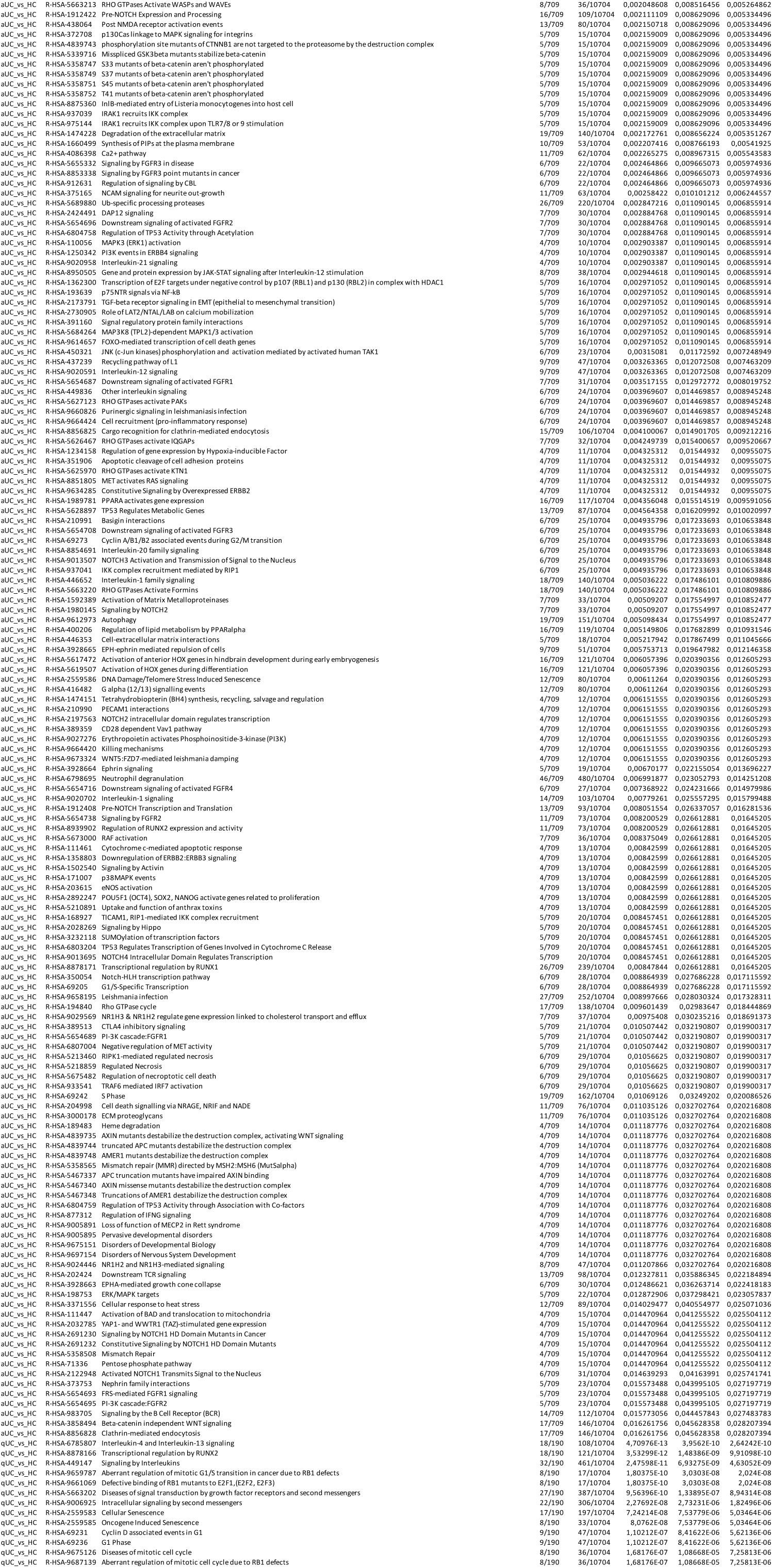
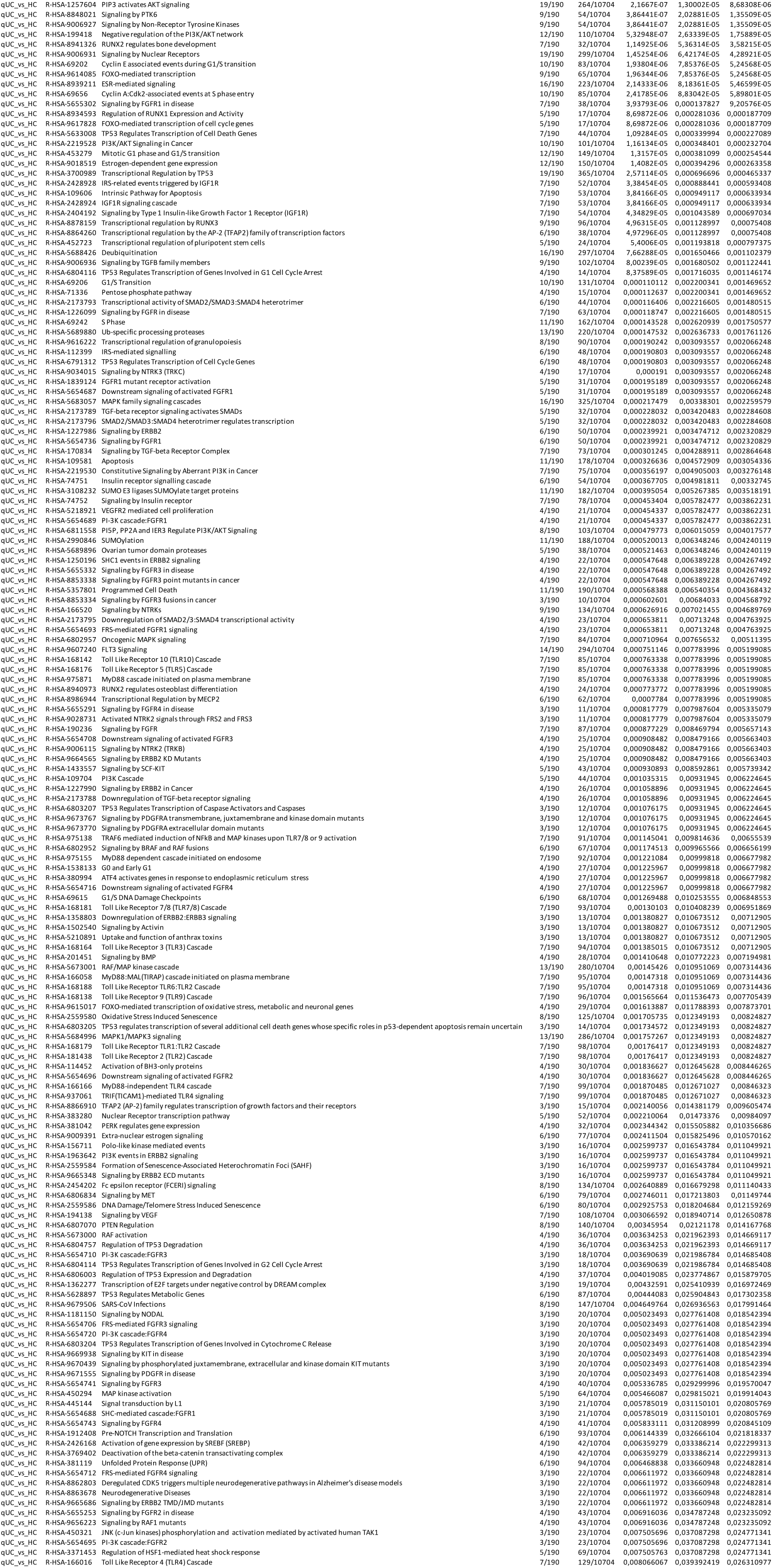
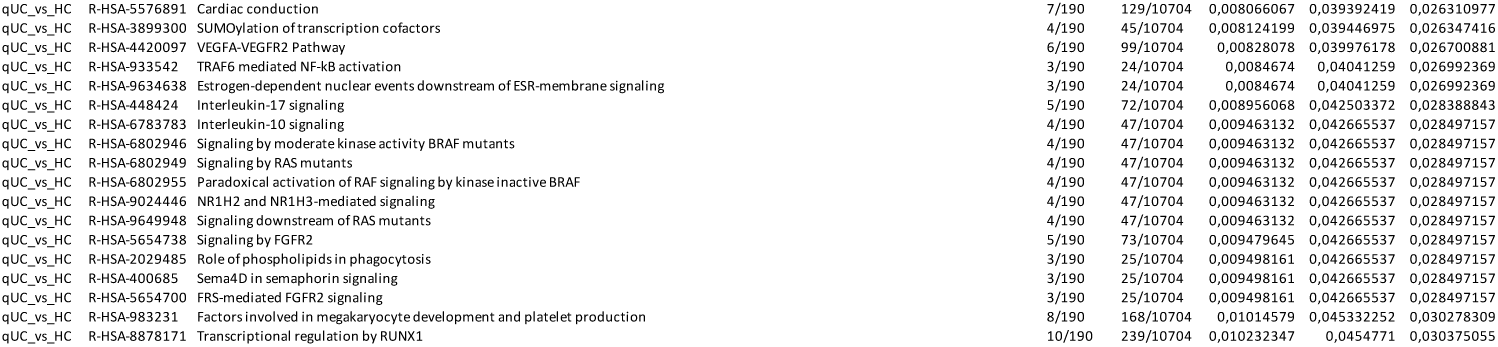
The list of Reactome pathways significantly enriched with differentially expressed colon tissue miRNAs in active (aUC) and quiescent (qUC)

**Supplementary Table S4.**
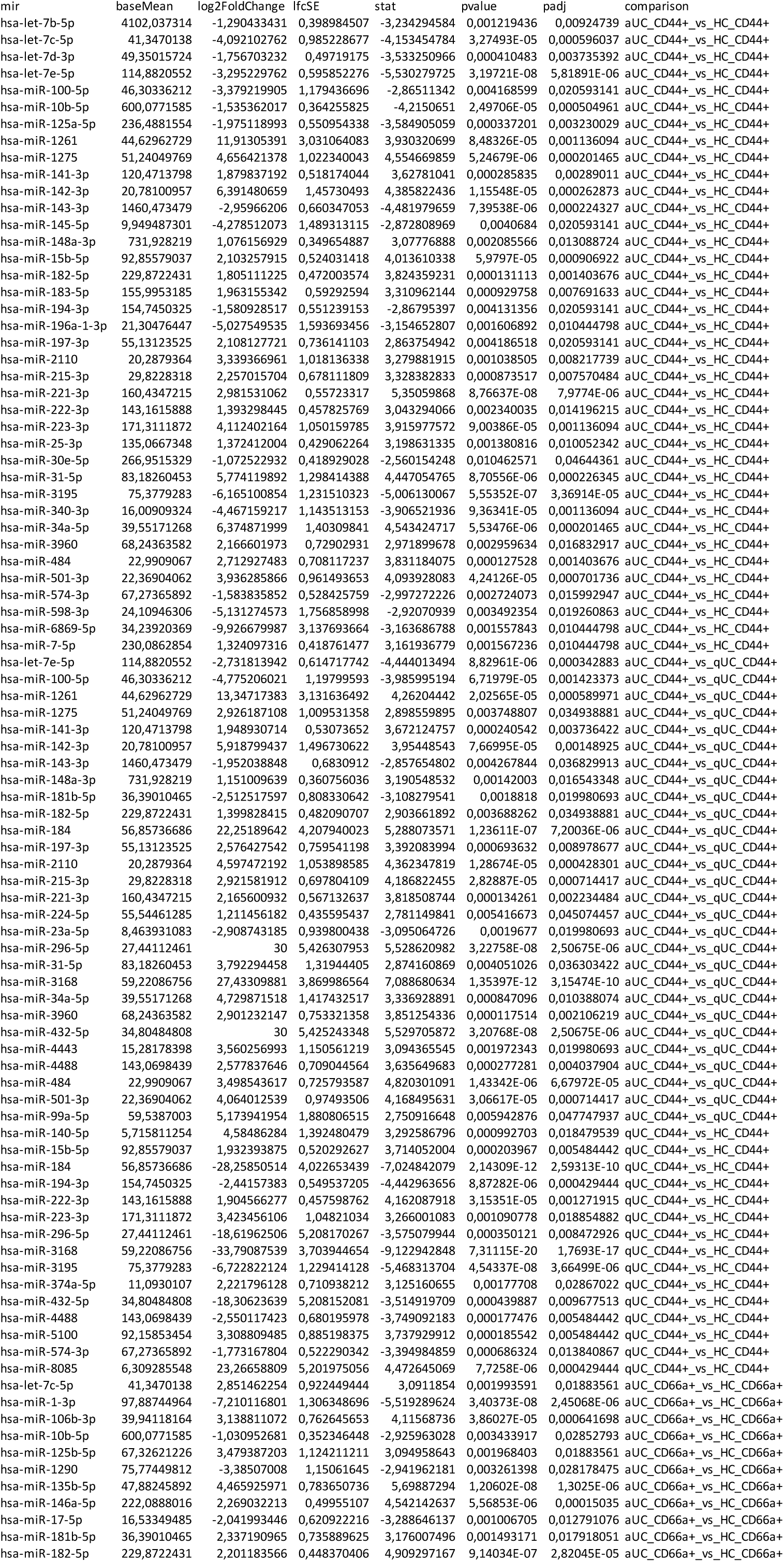
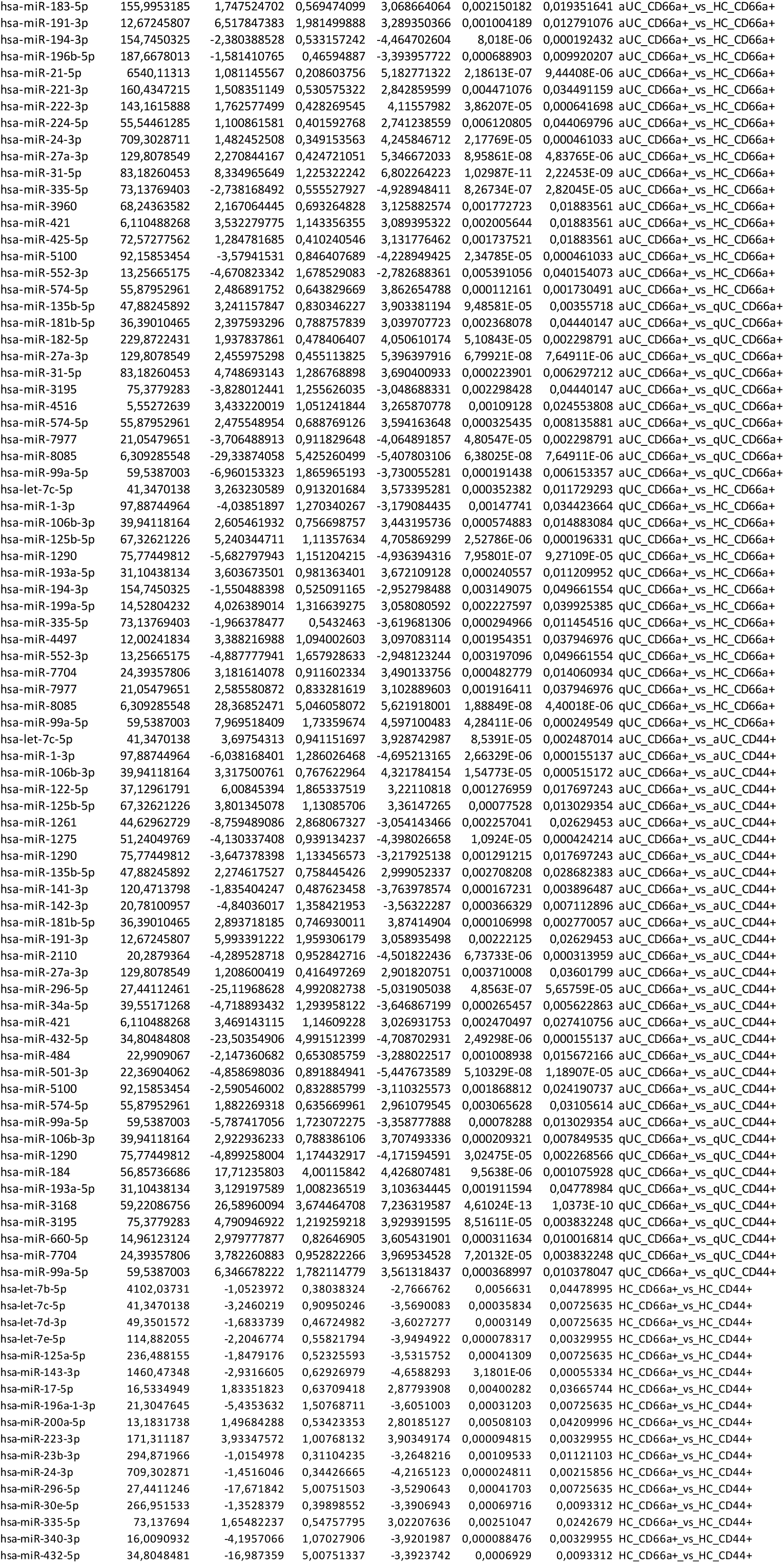
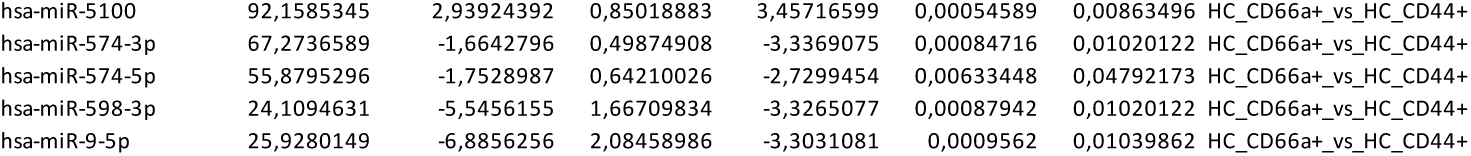
The list of differentially expressed miRNAs in colonic epithelial cell populations of active (aUC) and quiescent (qUC) UC patients, and Healthy controls (HC)

**Supplementary Table S5.**
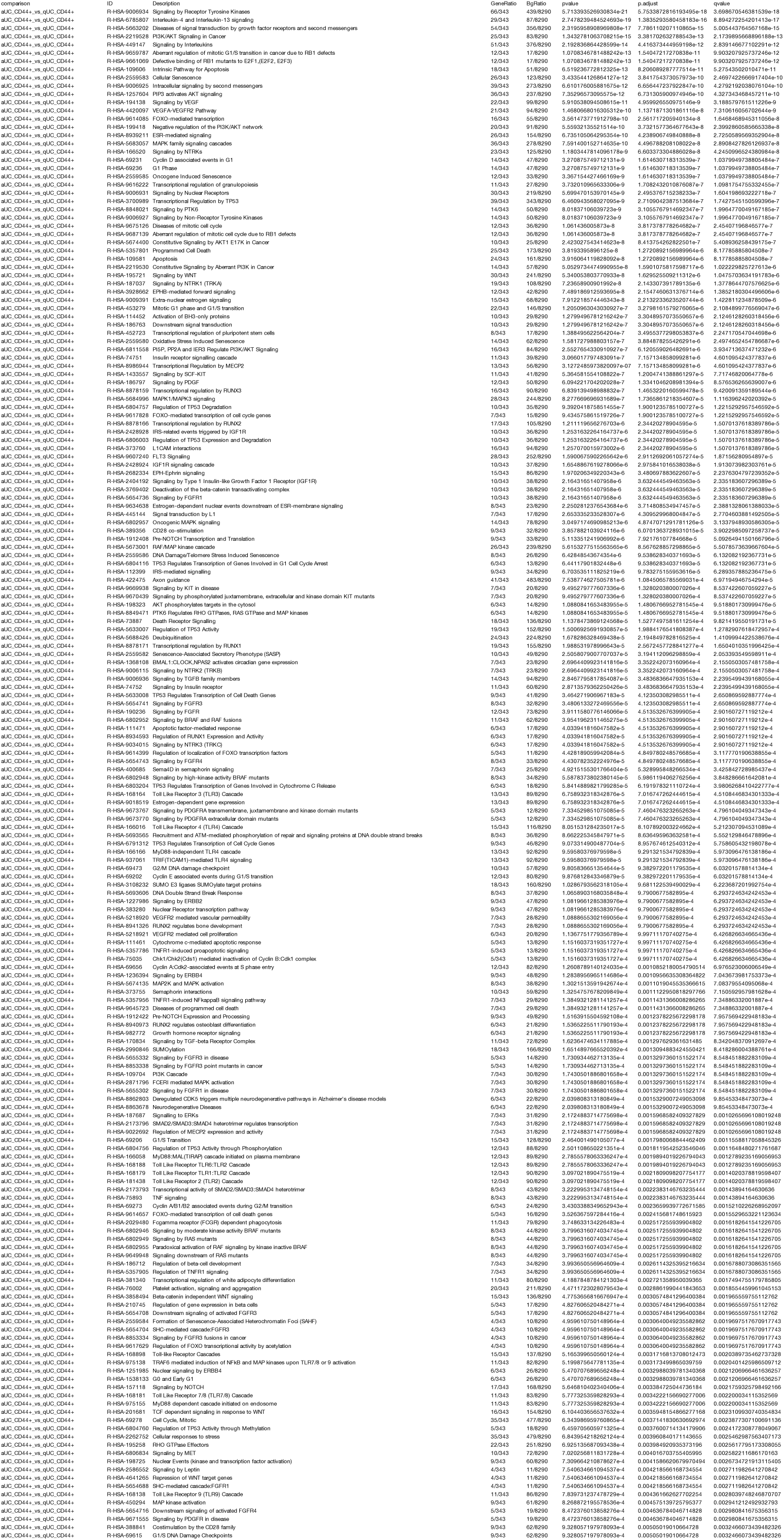
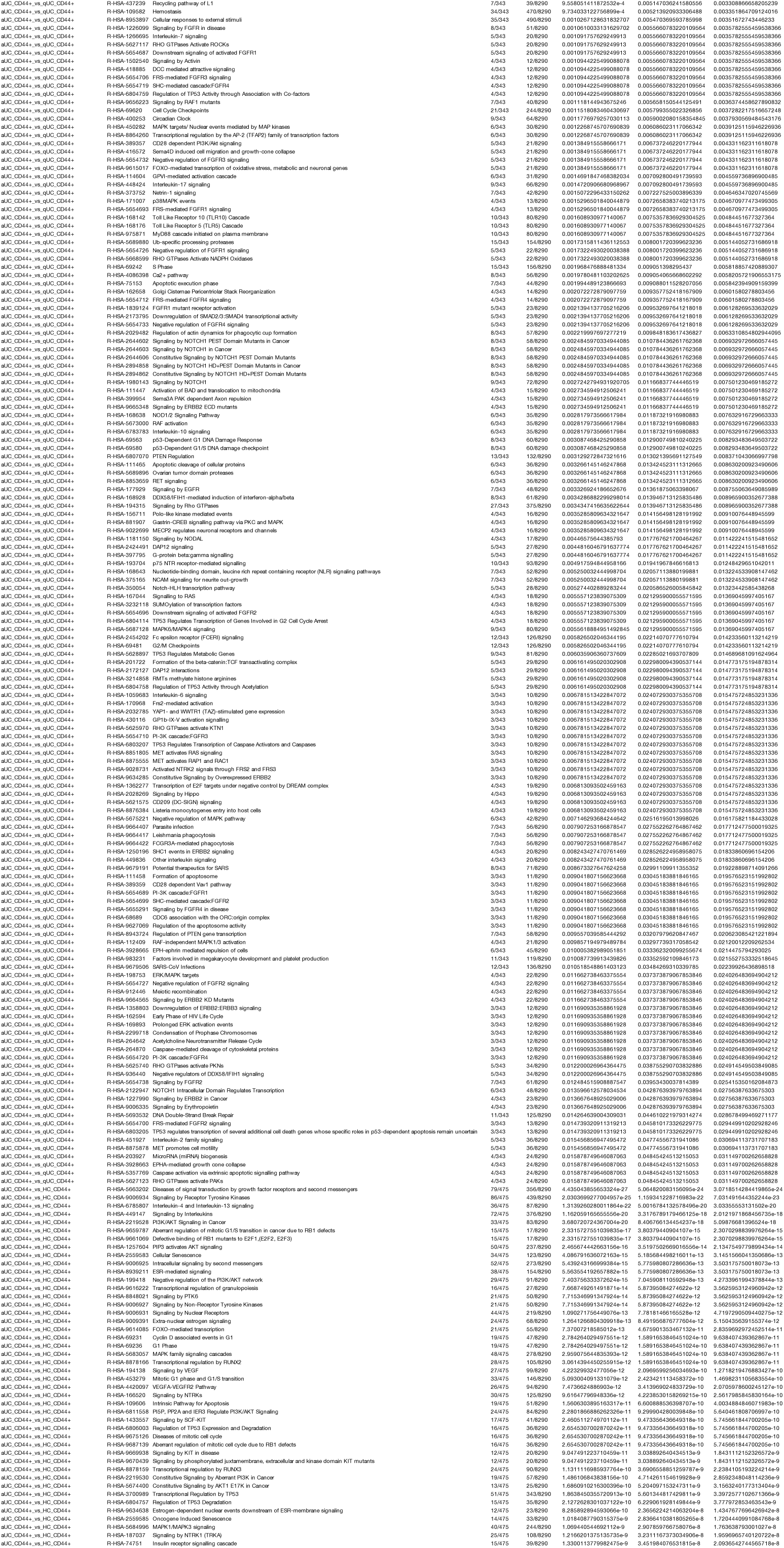
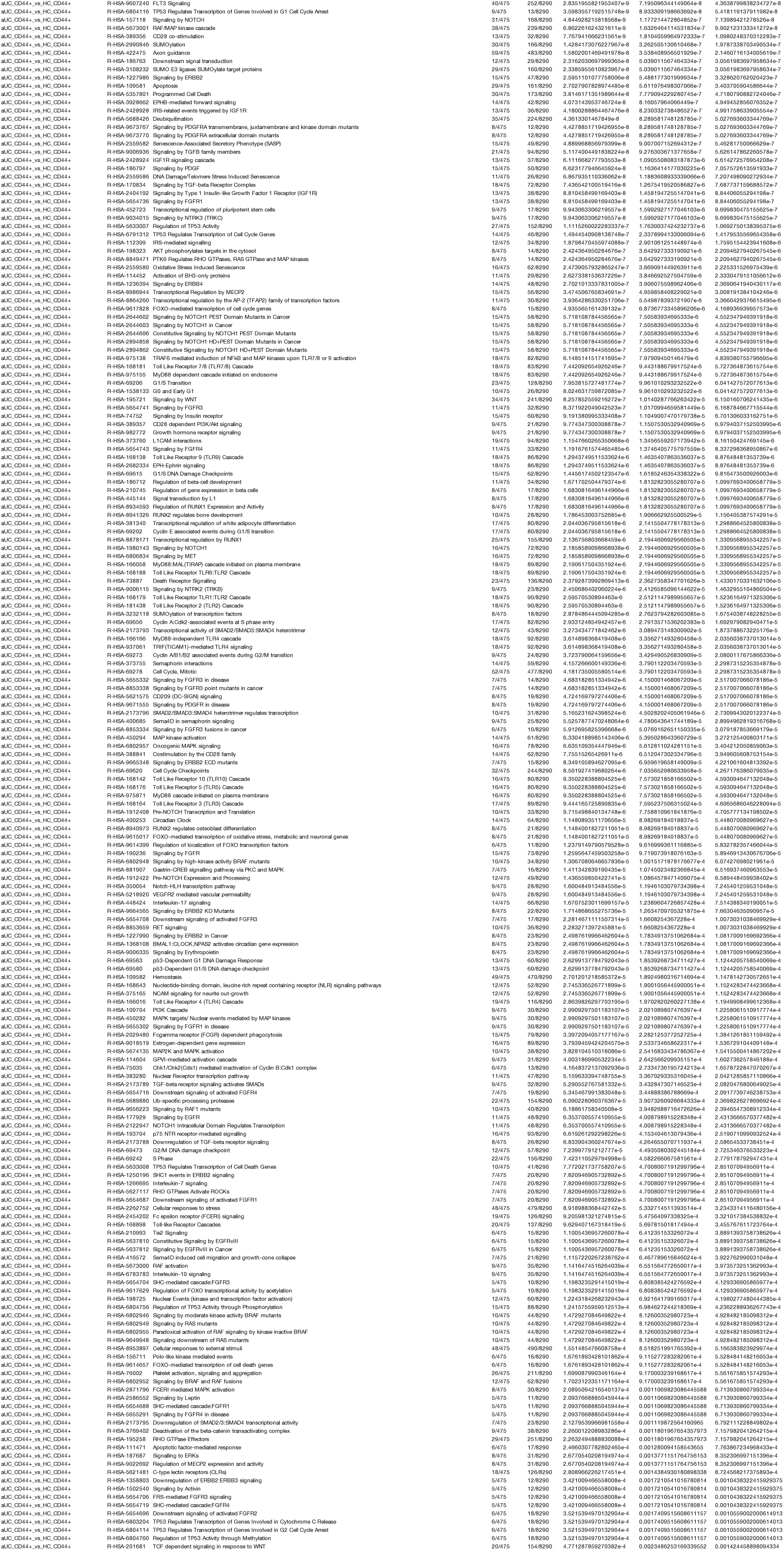
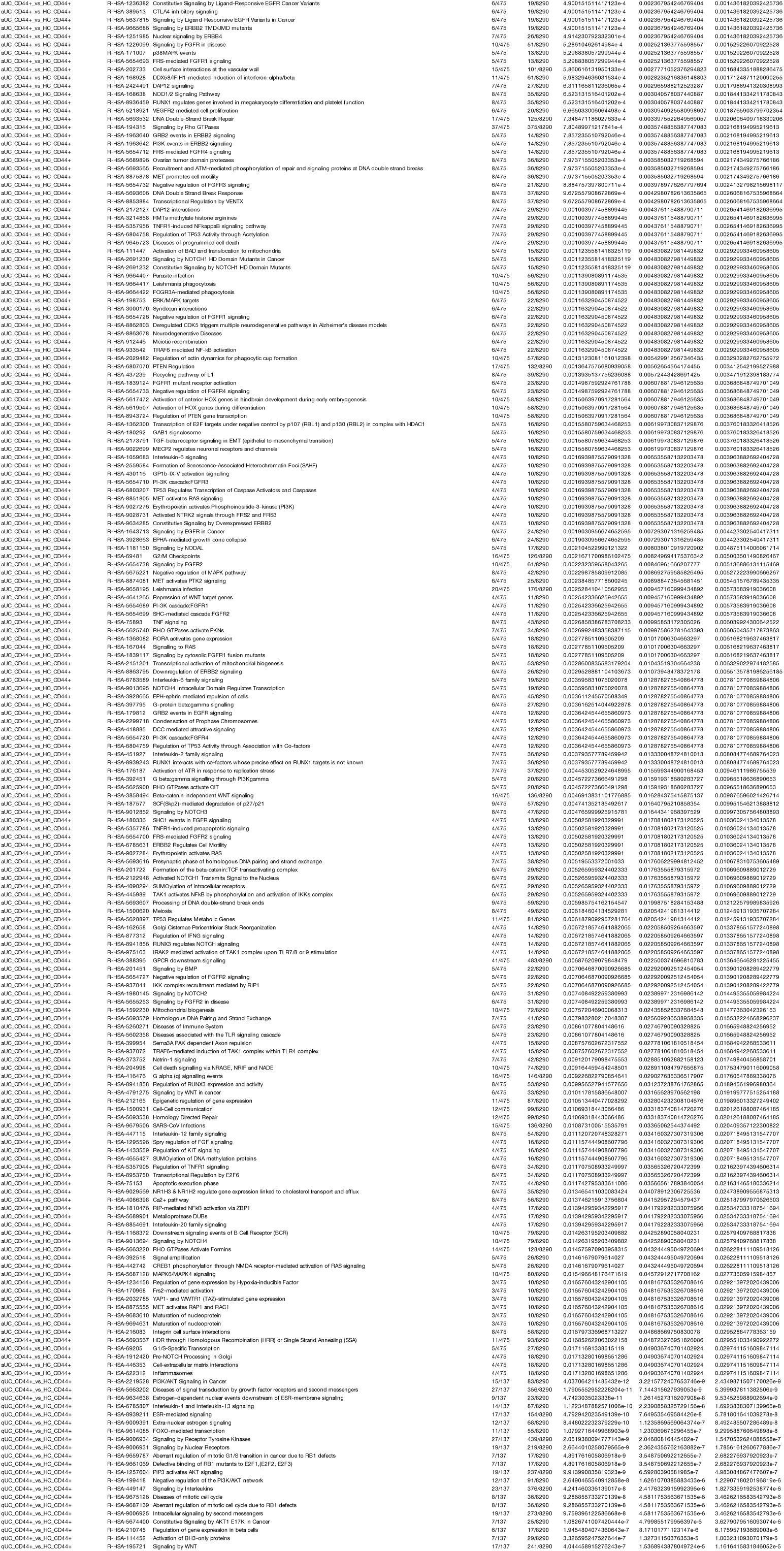
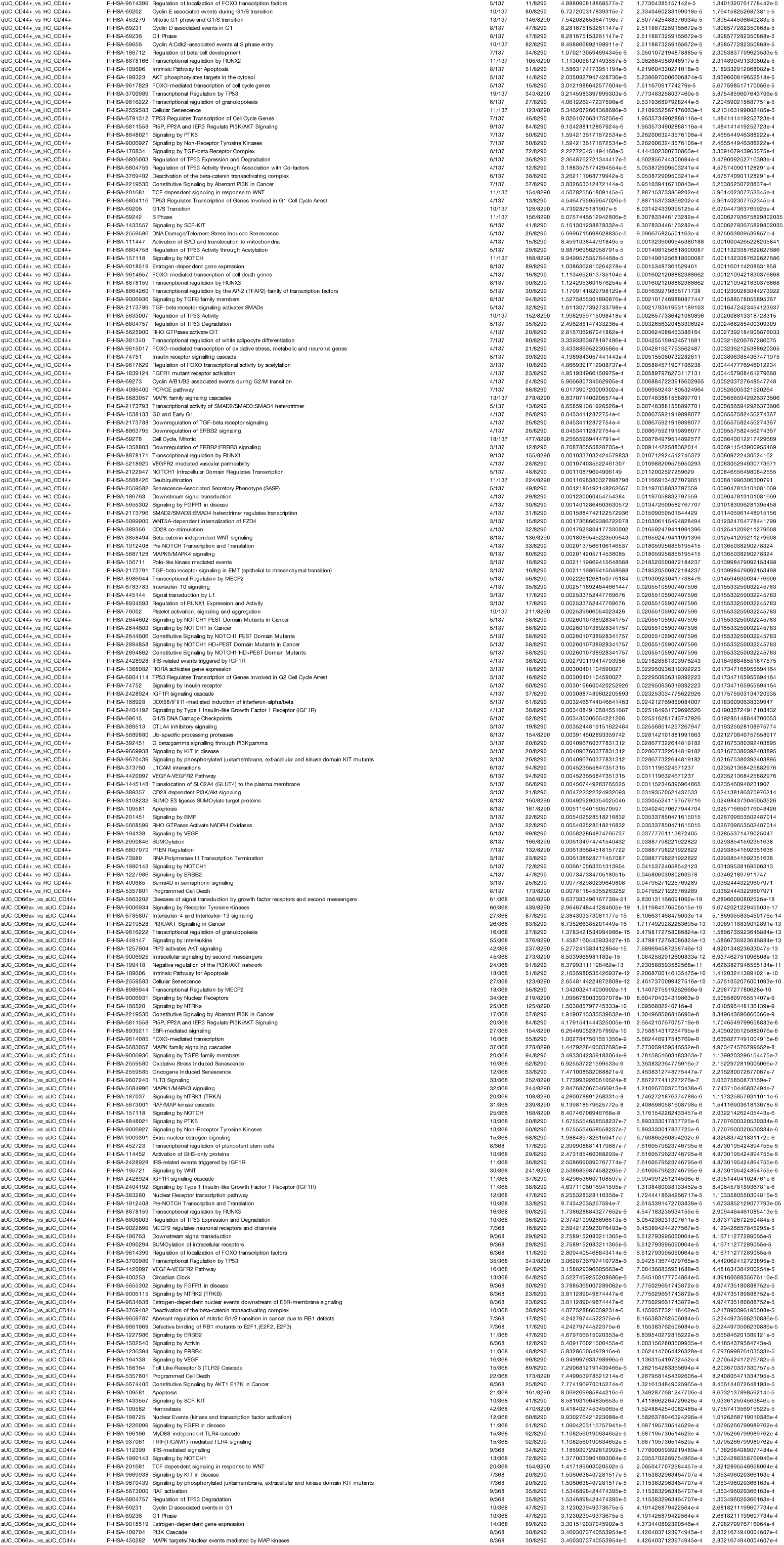
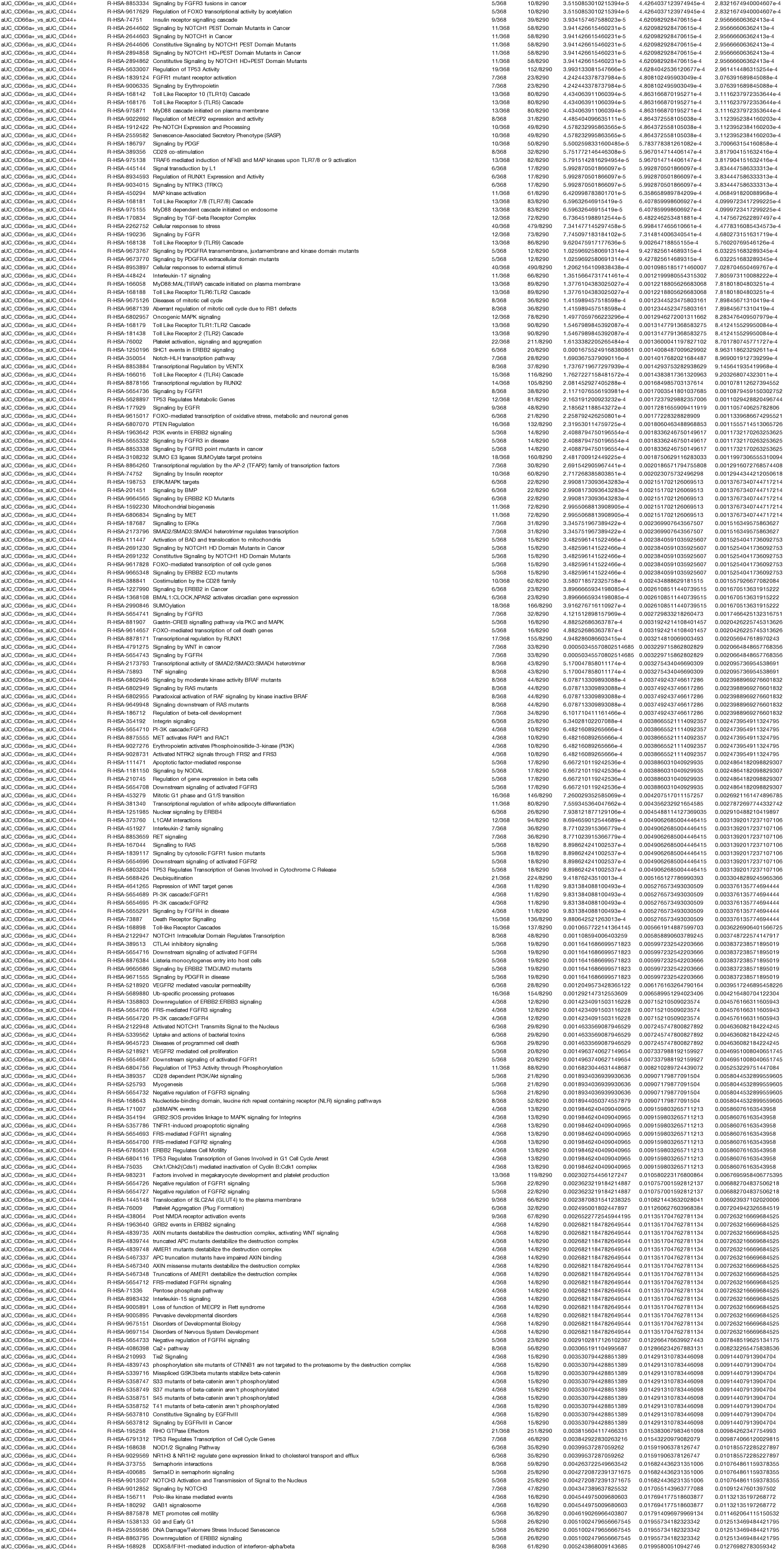
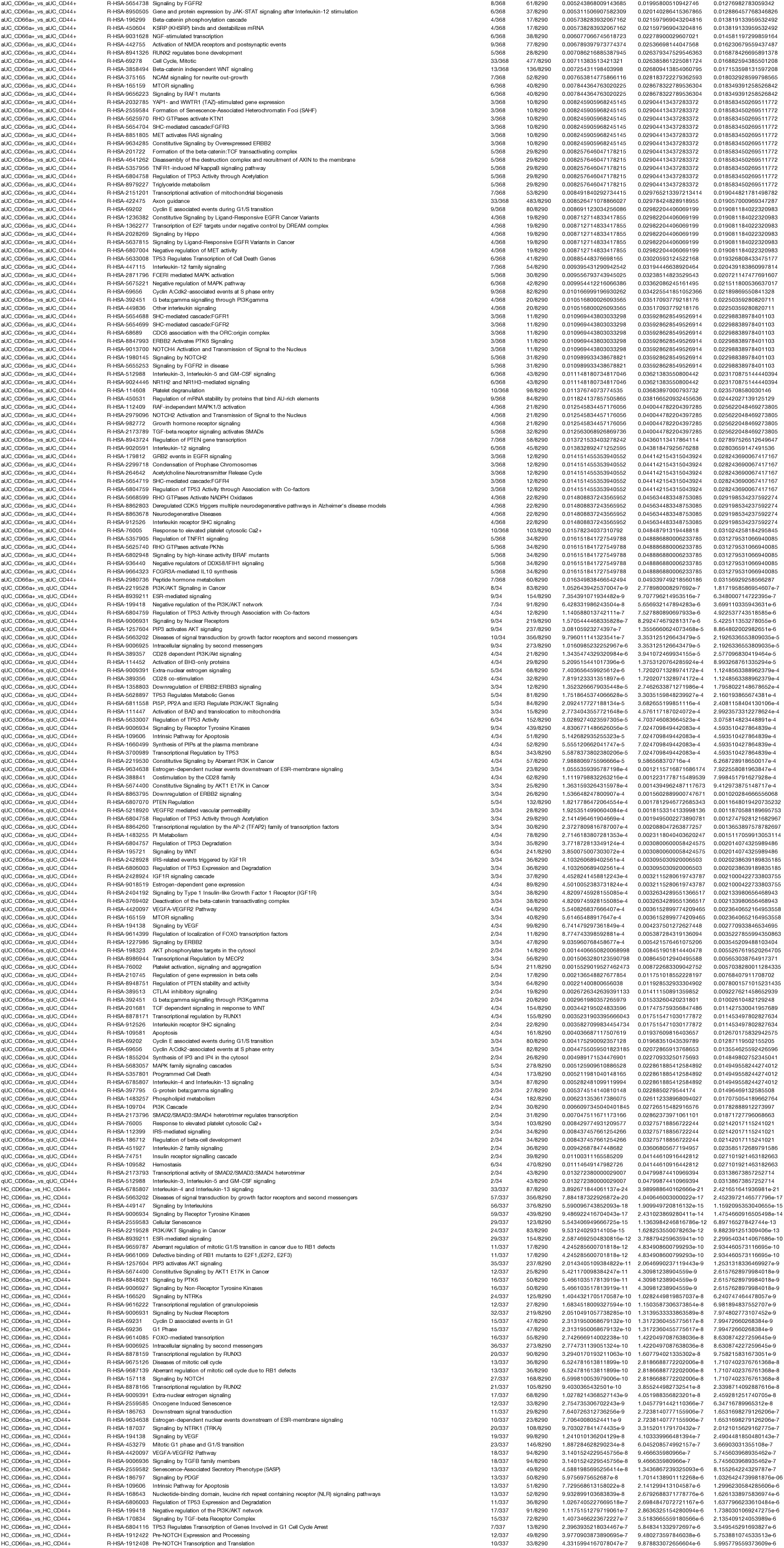
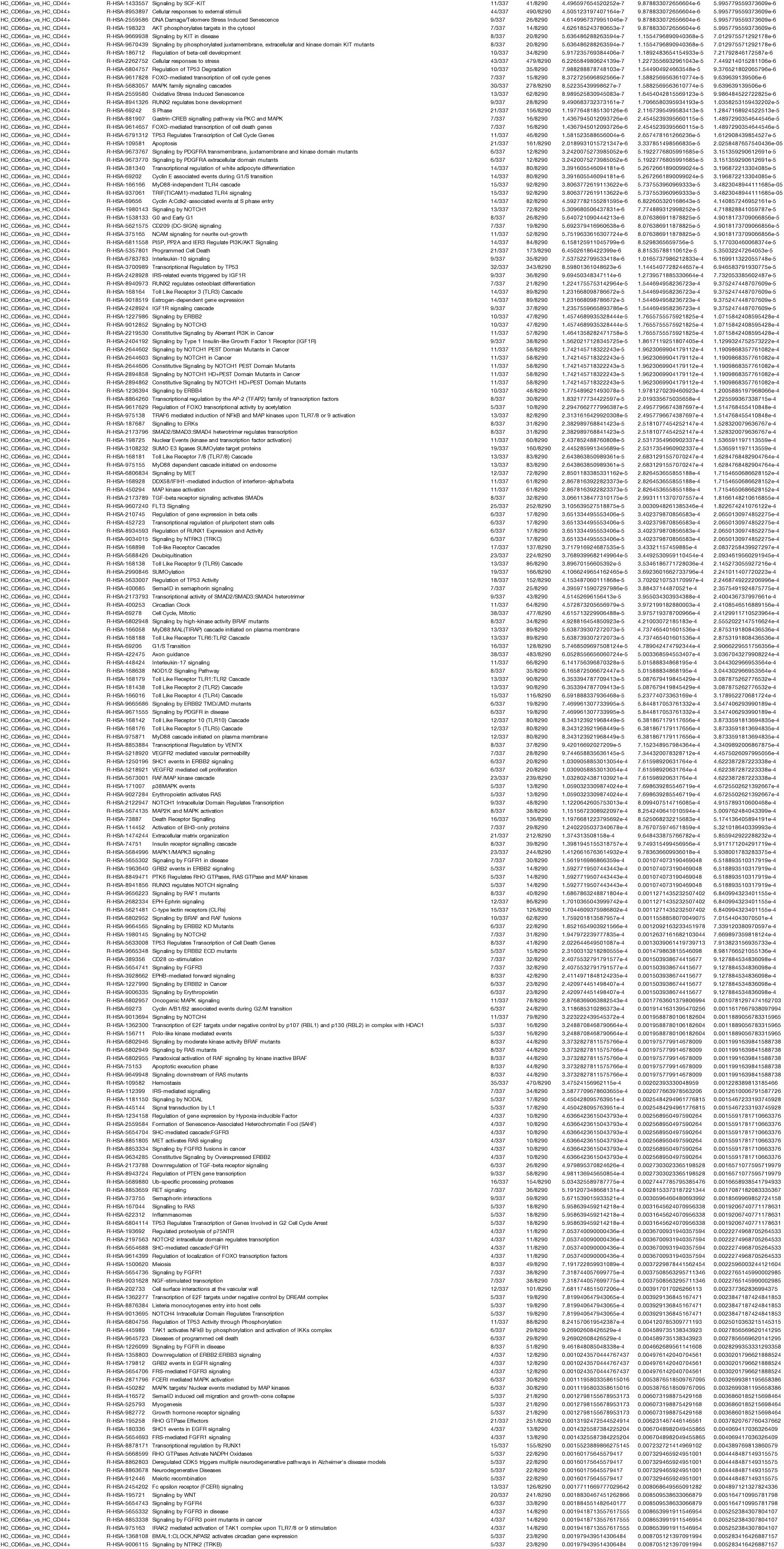
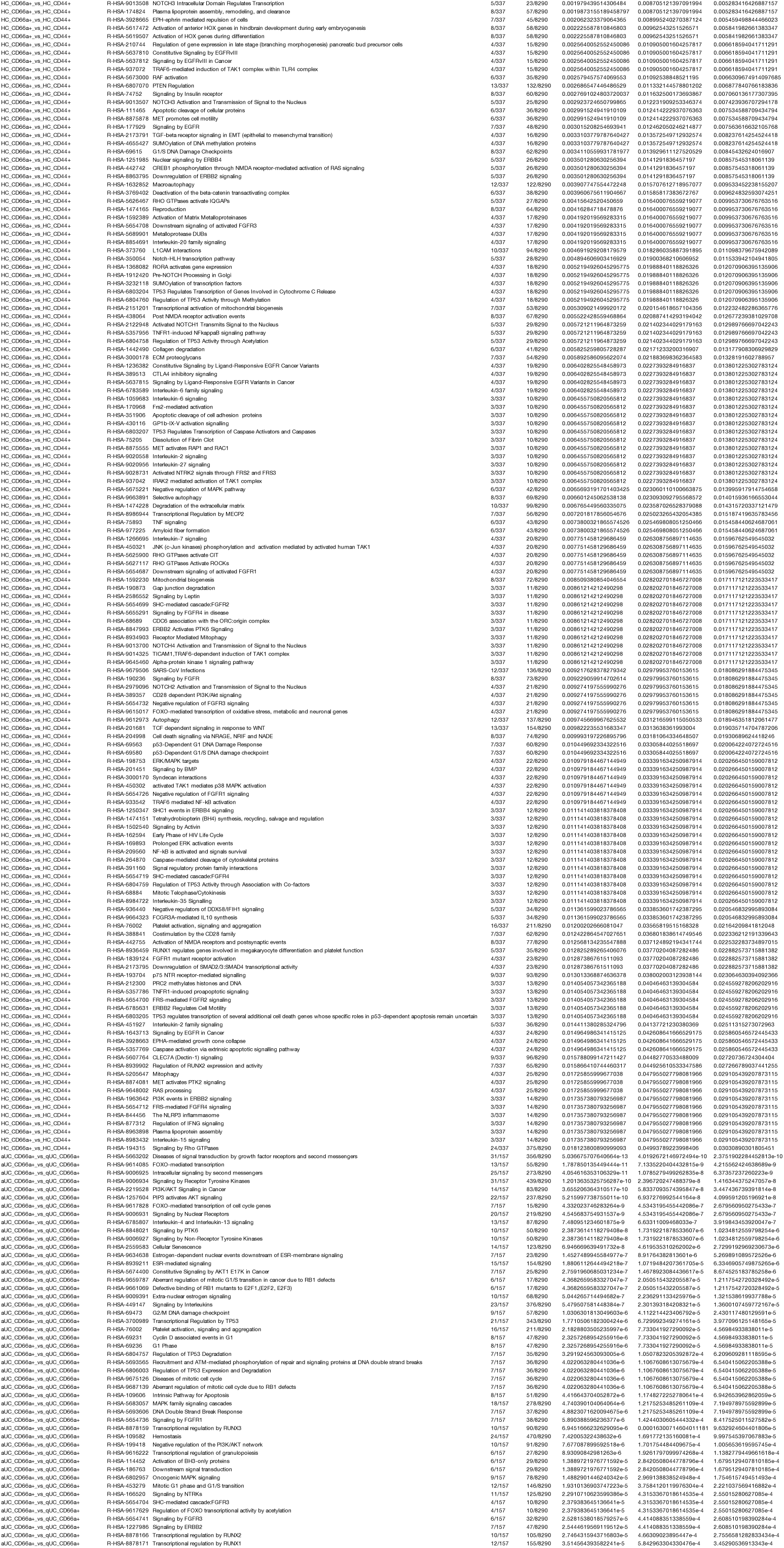
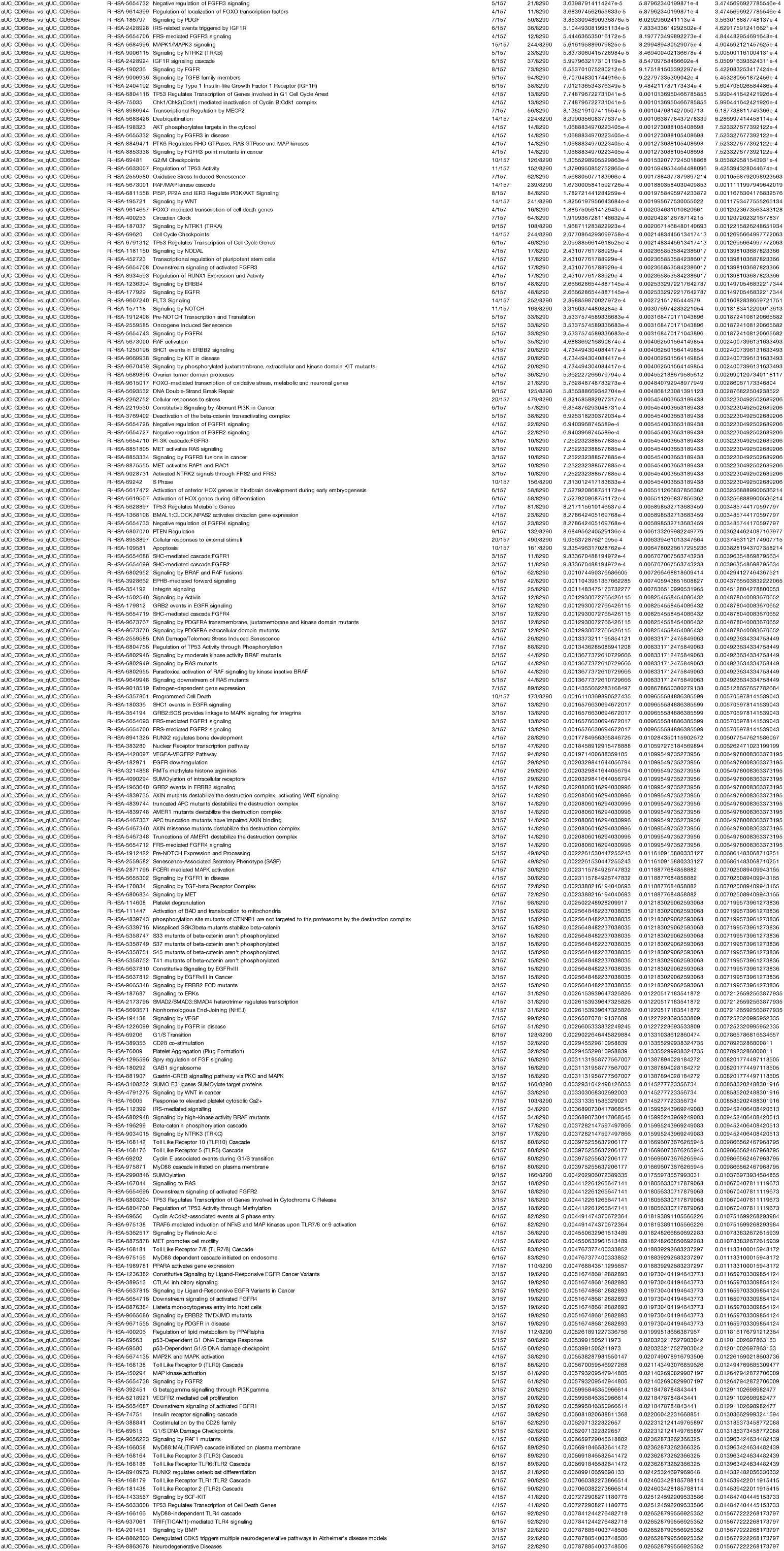
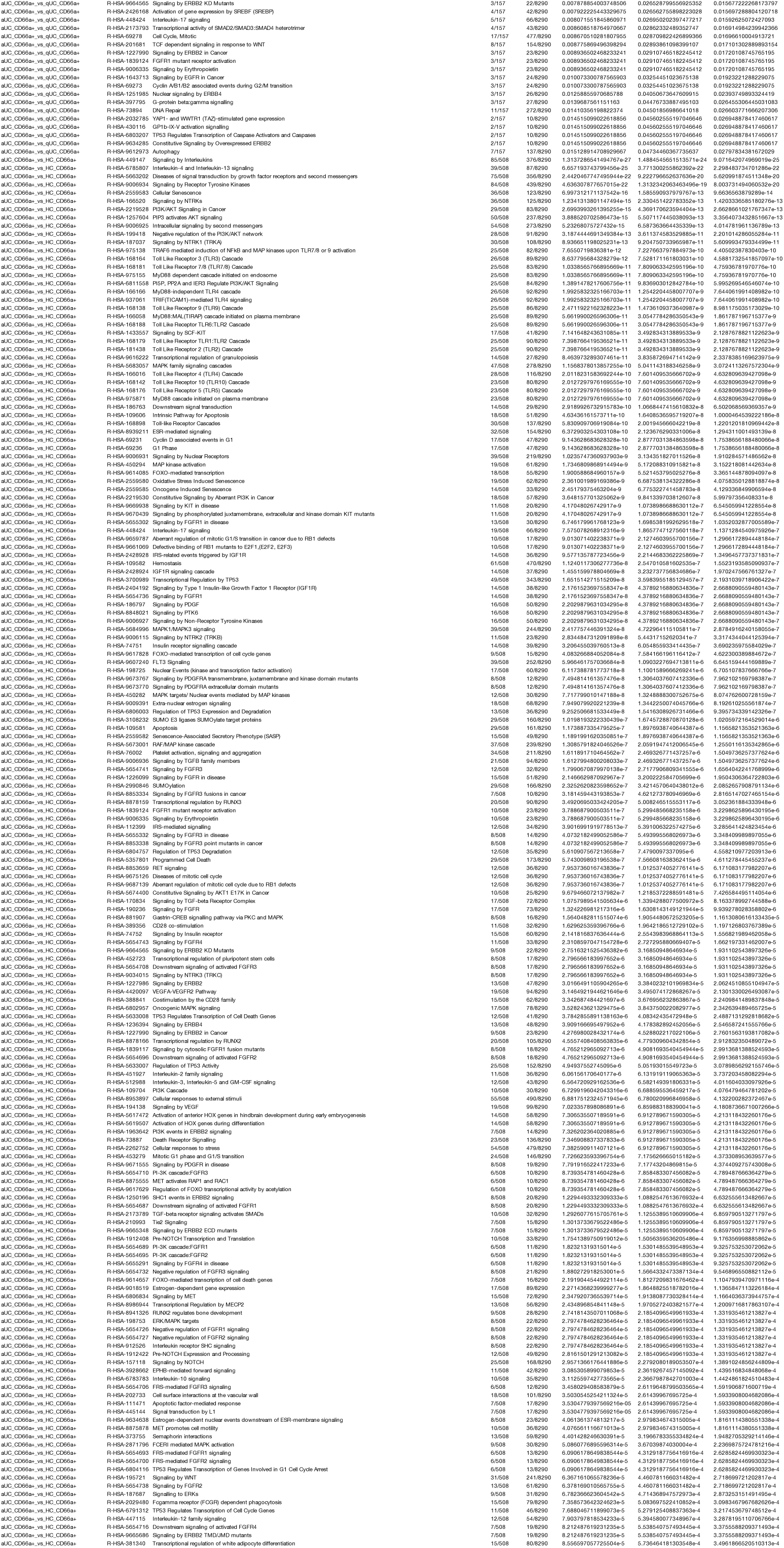
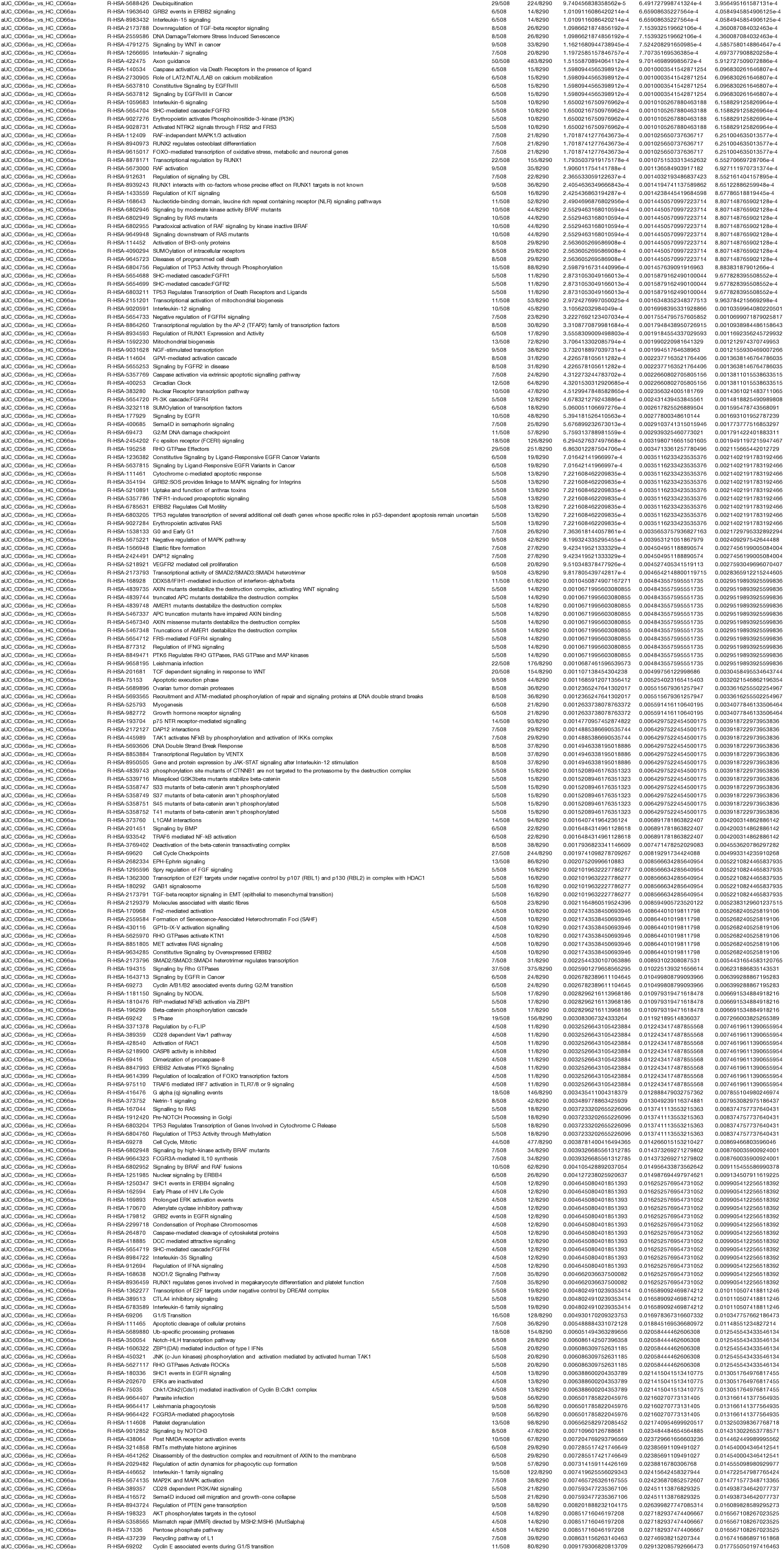
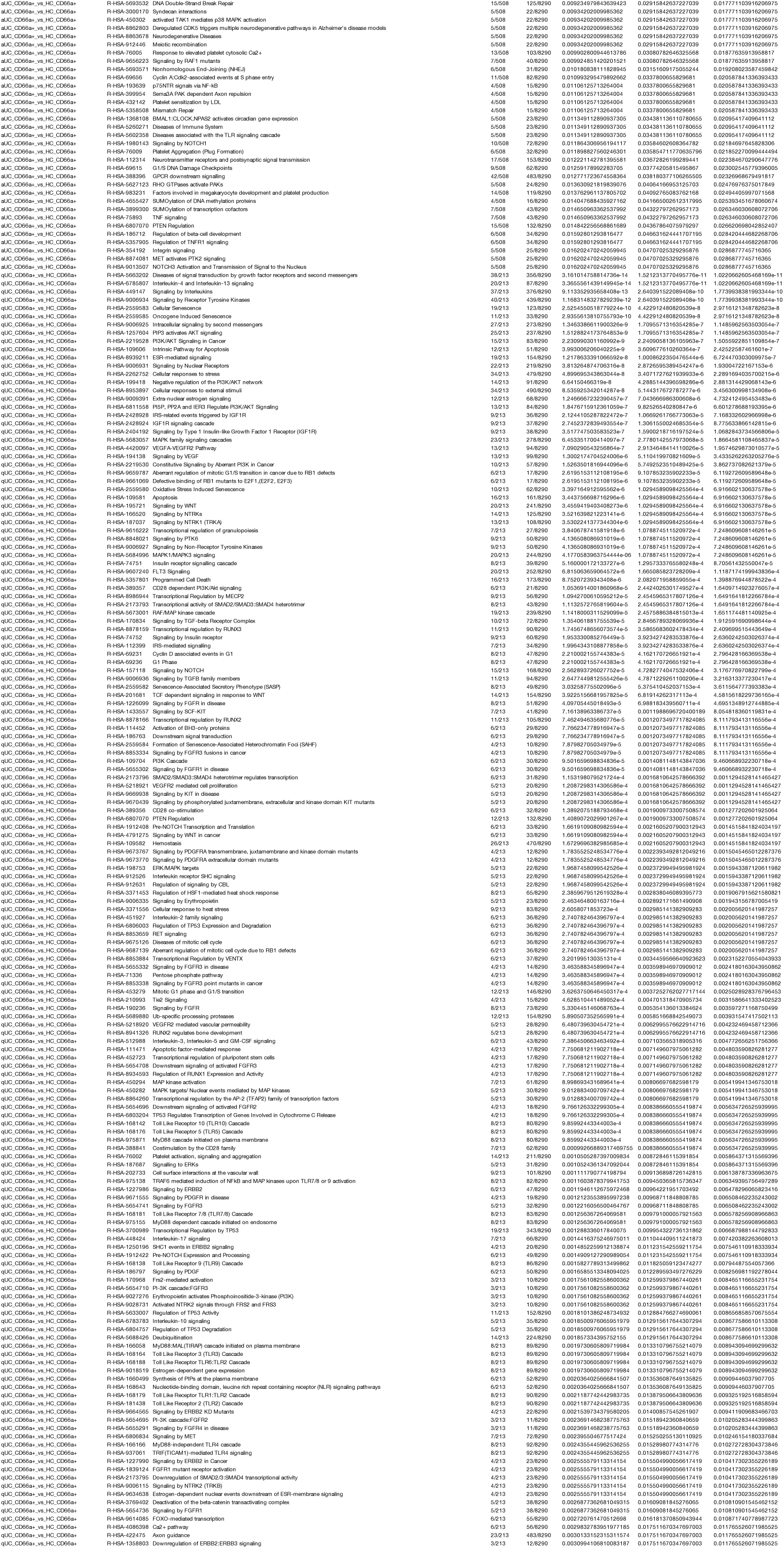
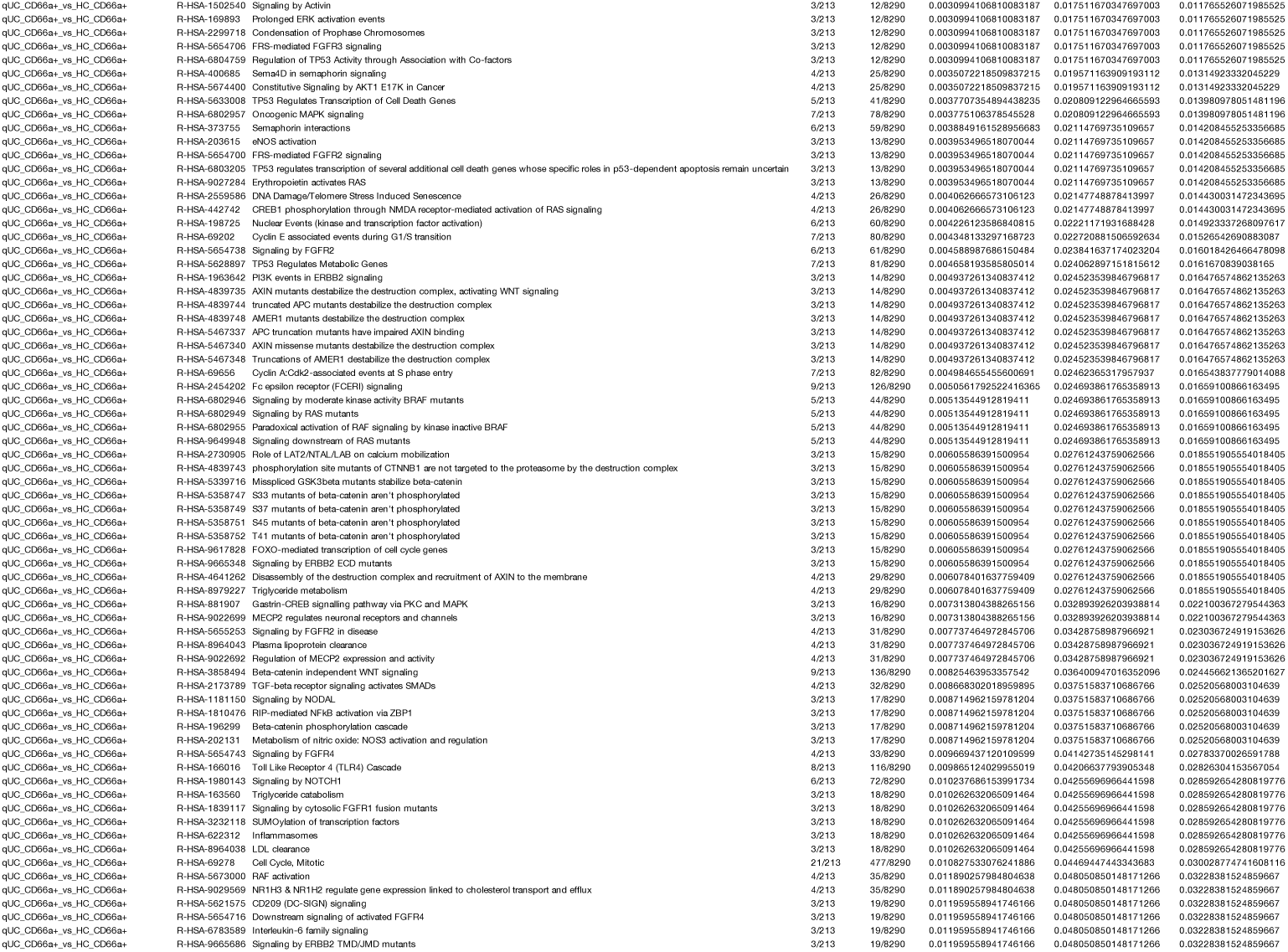
The list of Reactome pathways significantly enriched with differentially expressed miRNAs in colonic epithelial cell populations of active (aUC) and quiescent (qUC) UC patients, healthy controls (HC)

**Supplementary Table S6.**
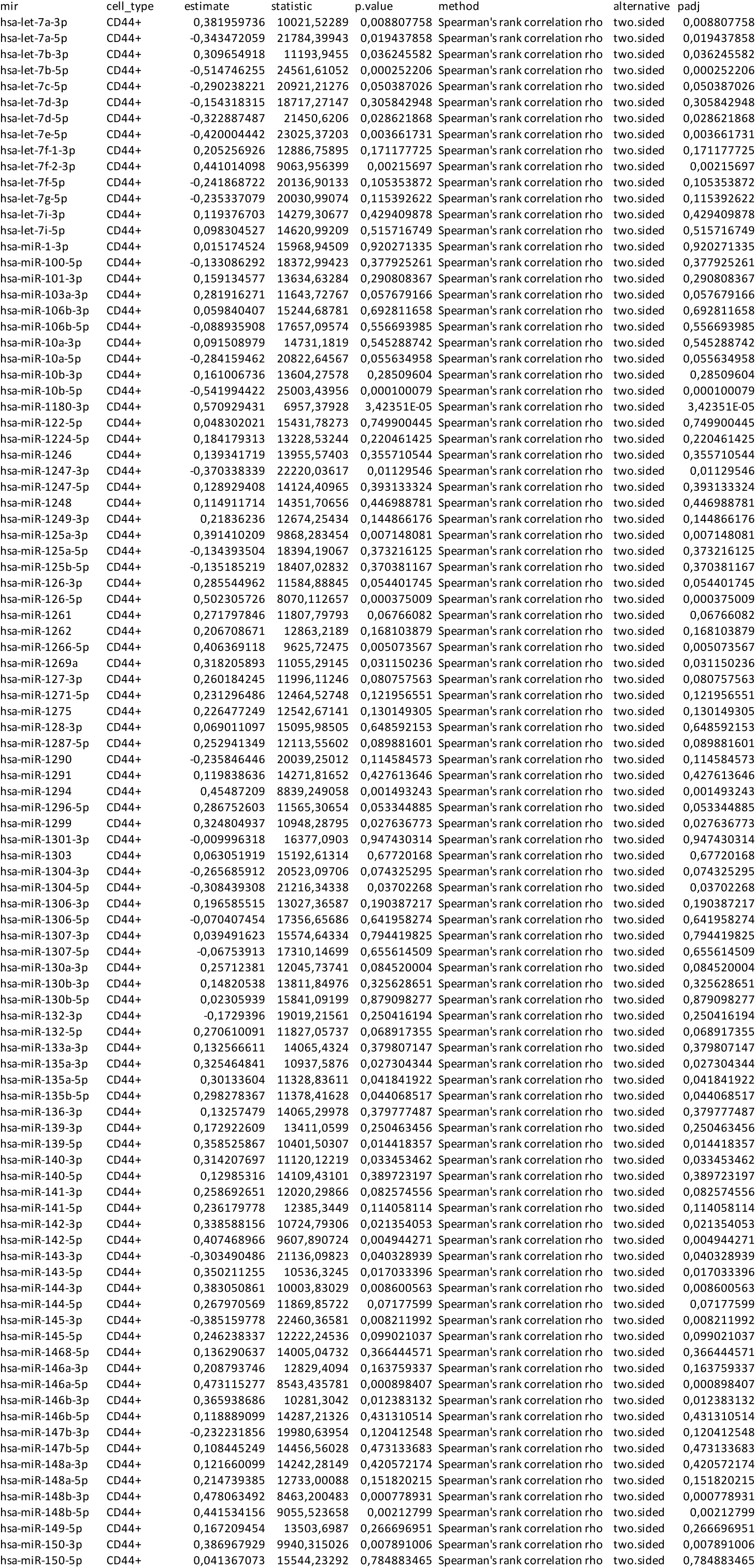
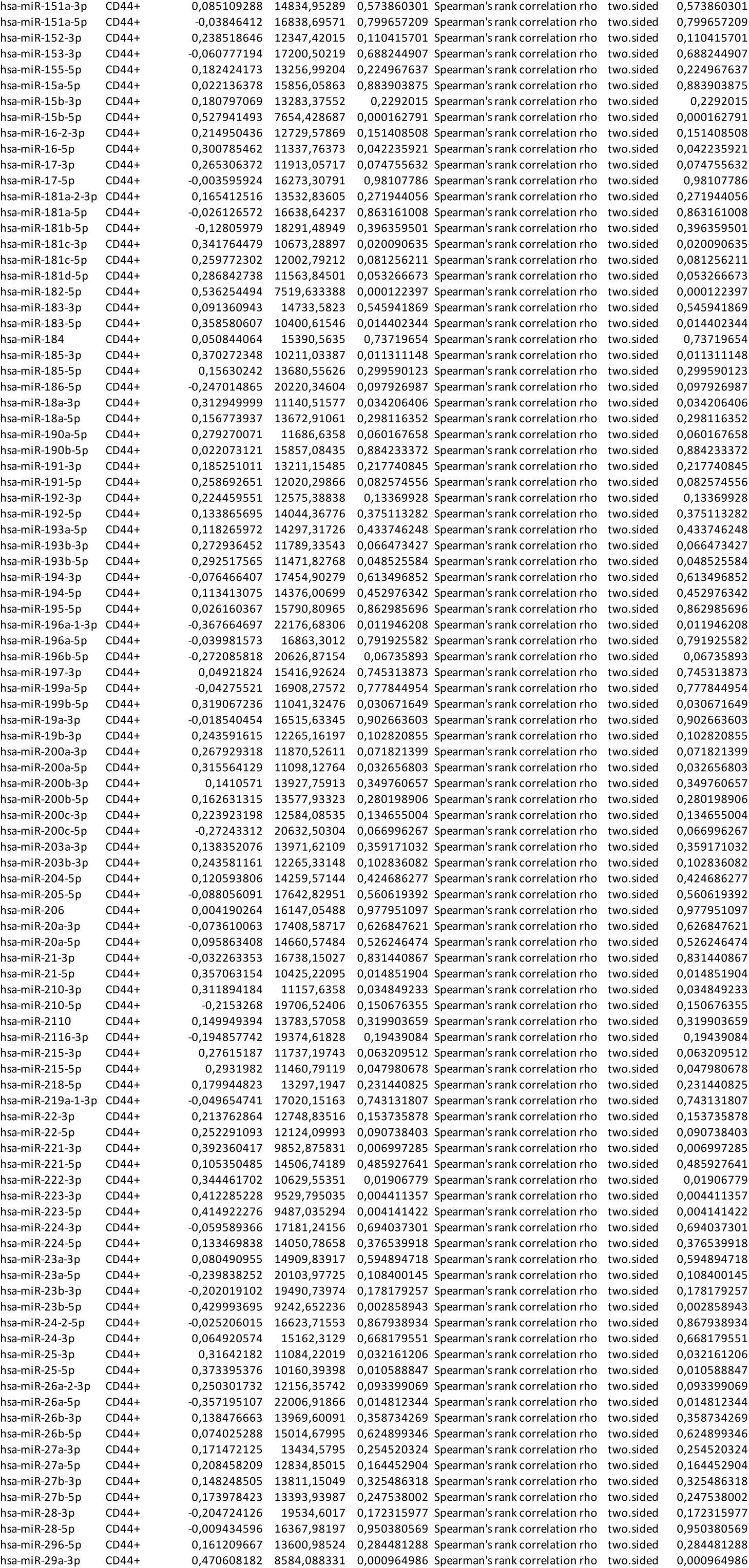
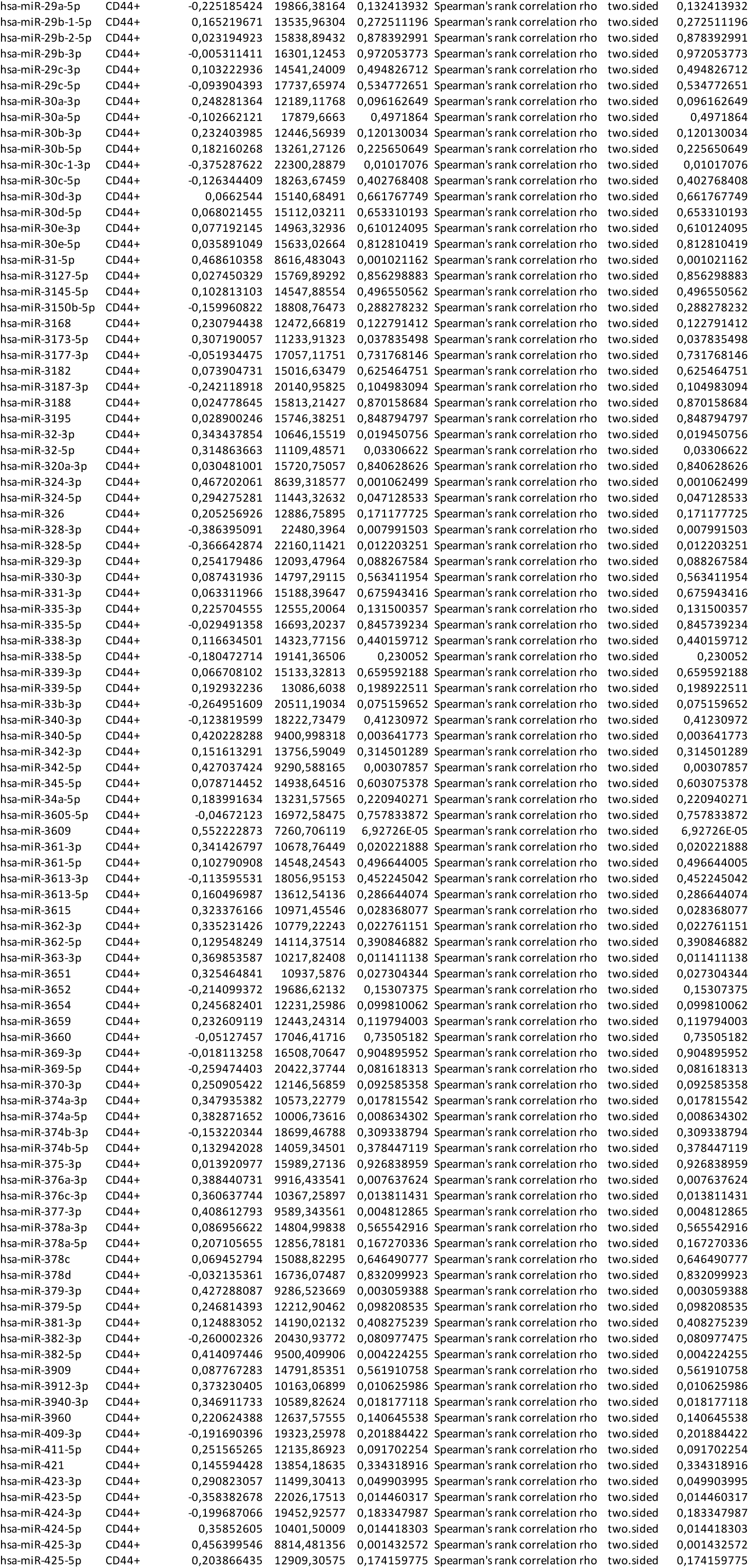
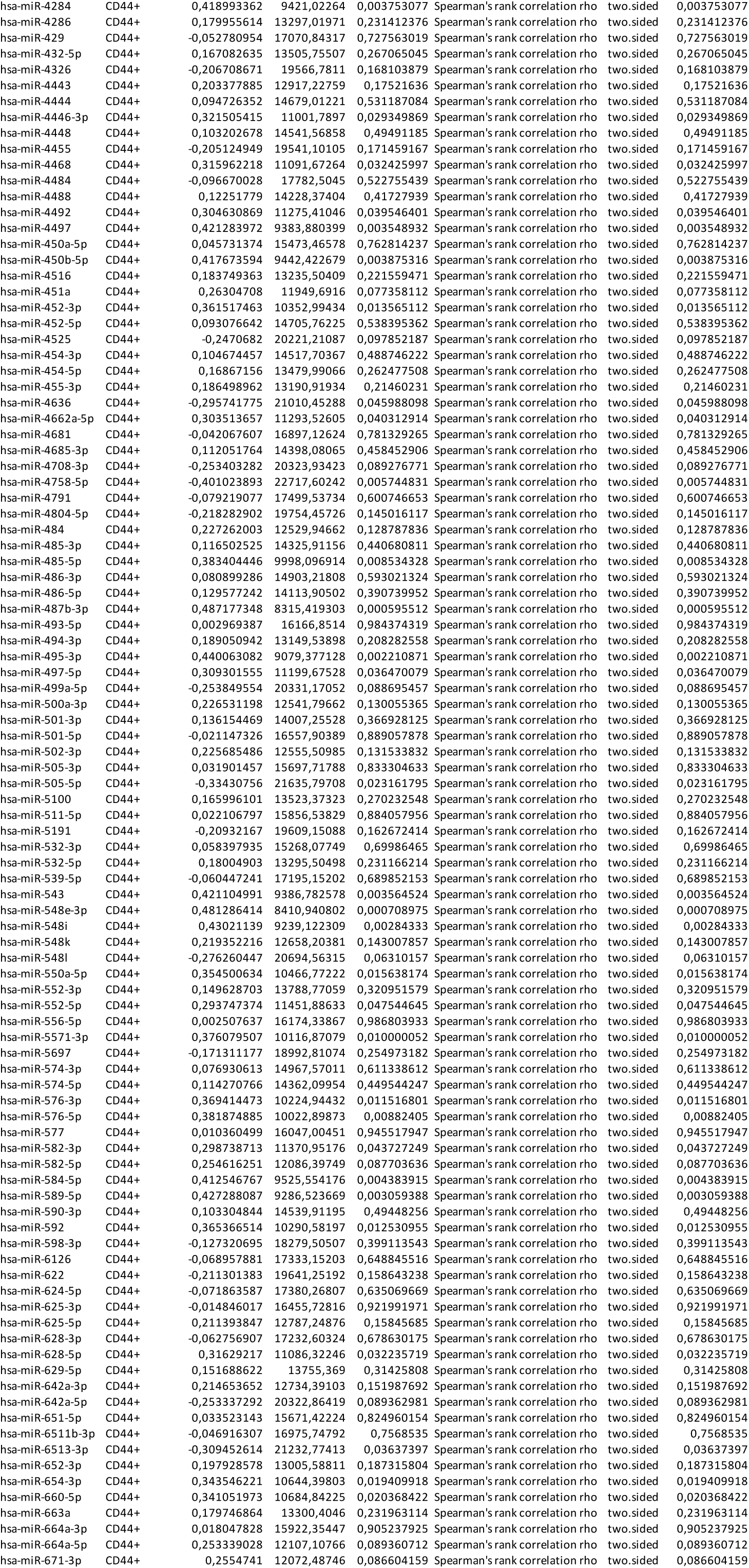
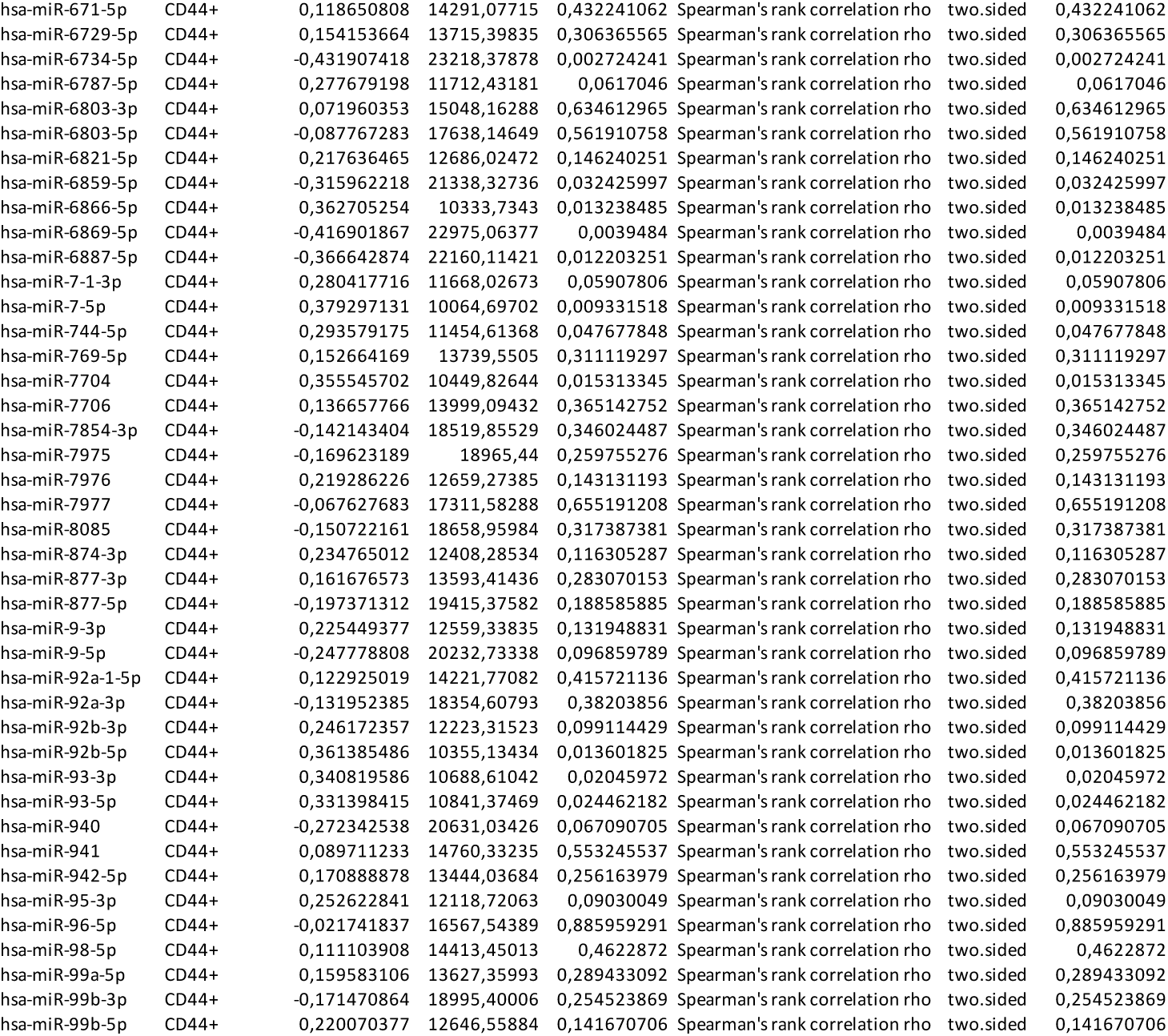
The list of correlations between endoscopic Mayo score and normalized expression of miRNAs expressed in crypt-bottom (CD44+) colonic epithelial cell population.

**Supplementary Table S7.**
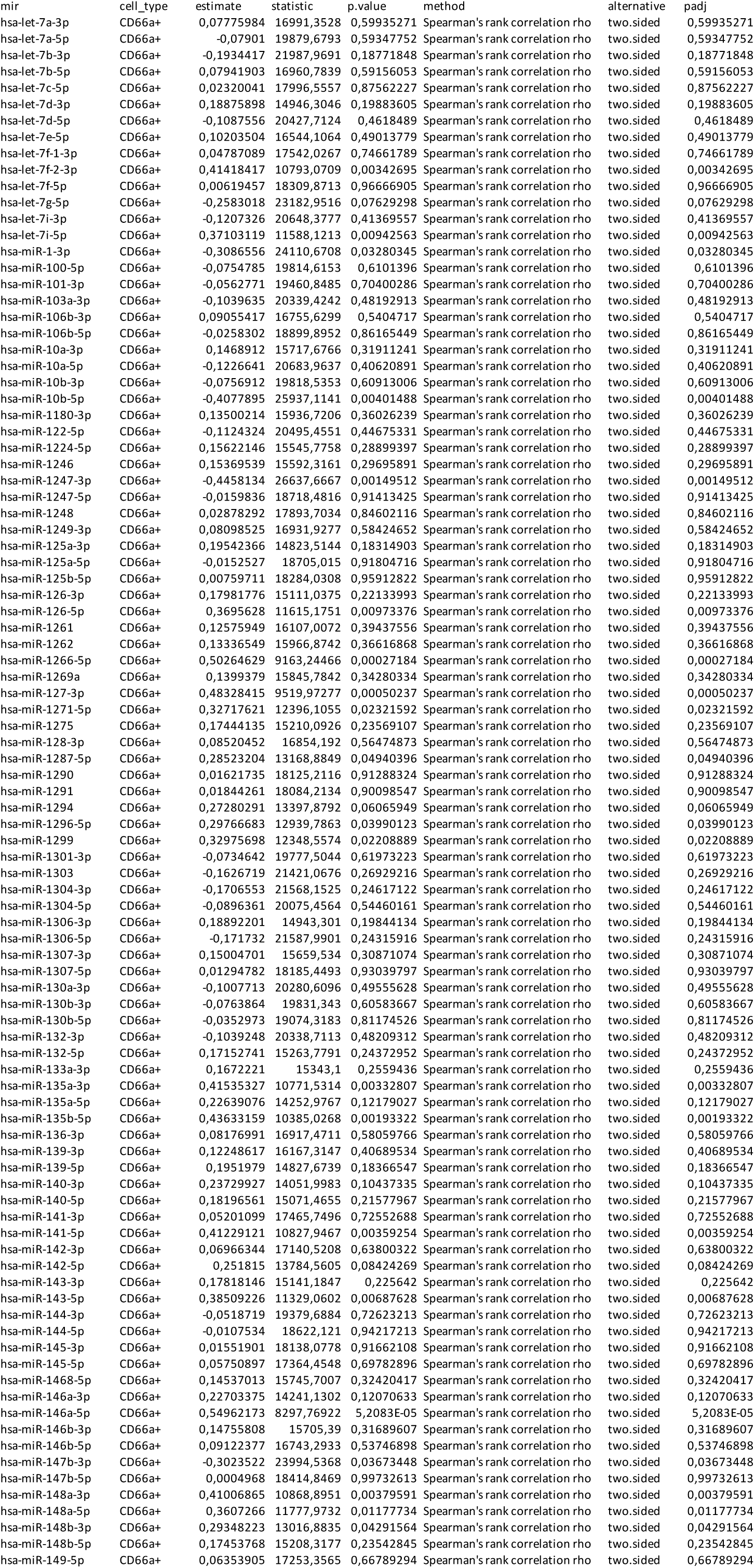
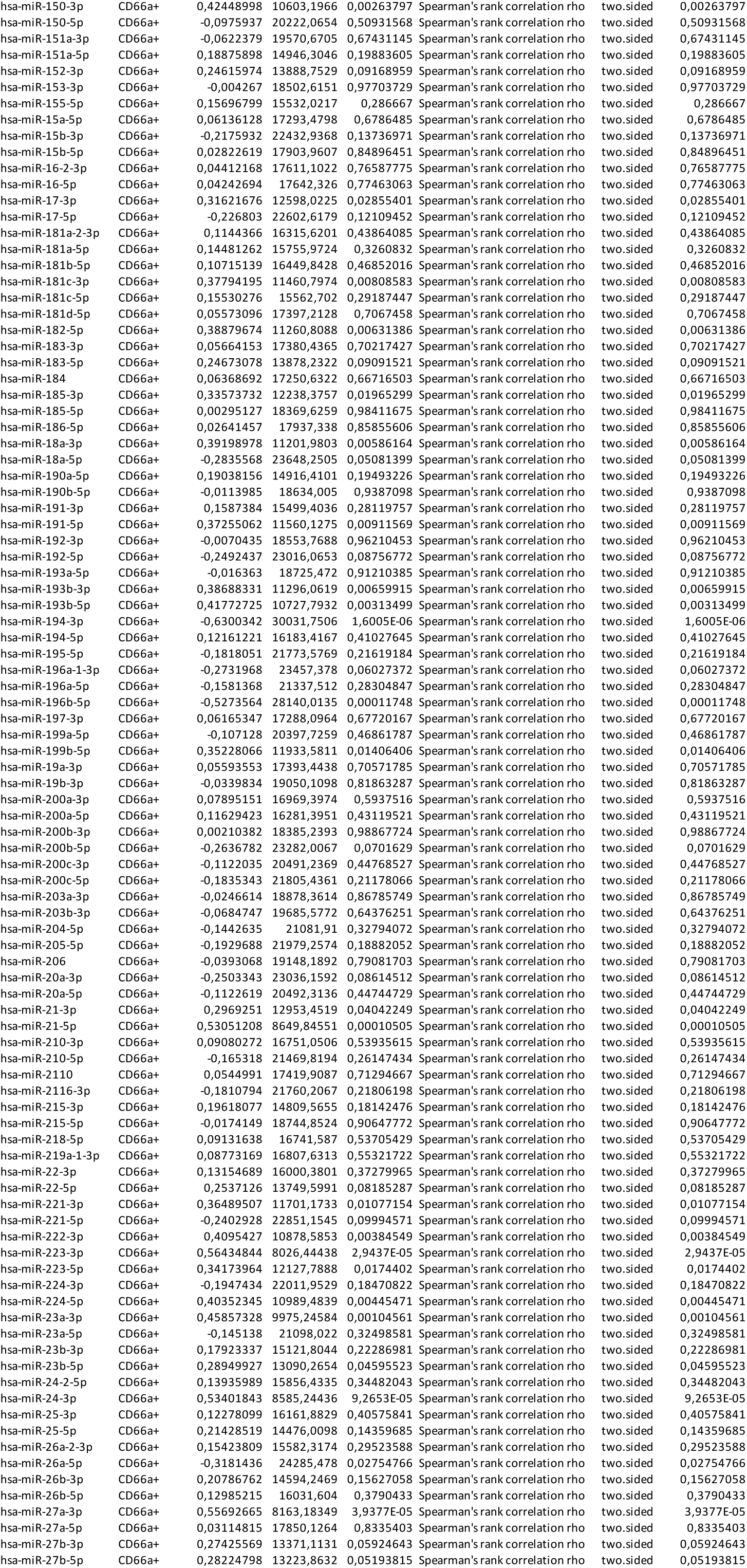
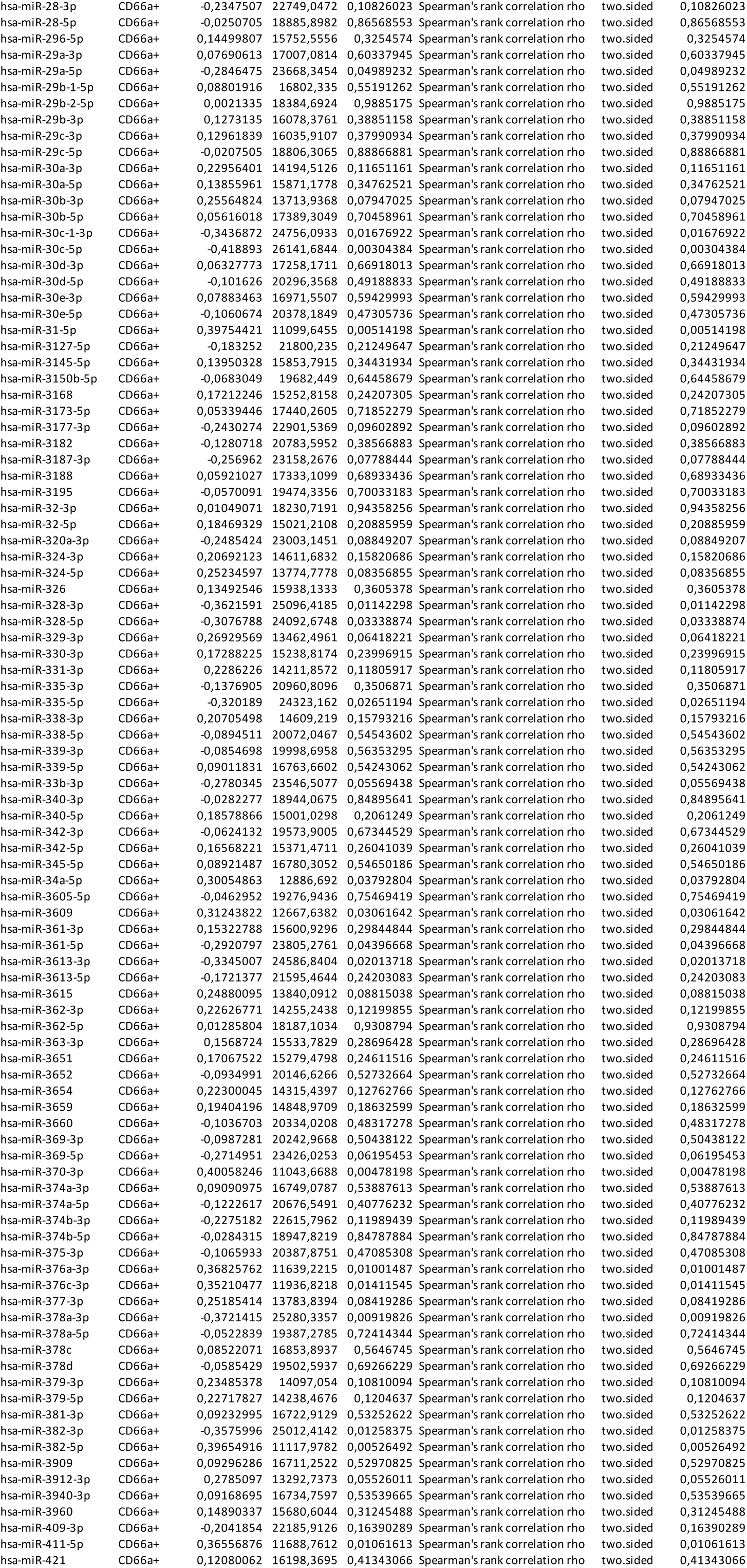
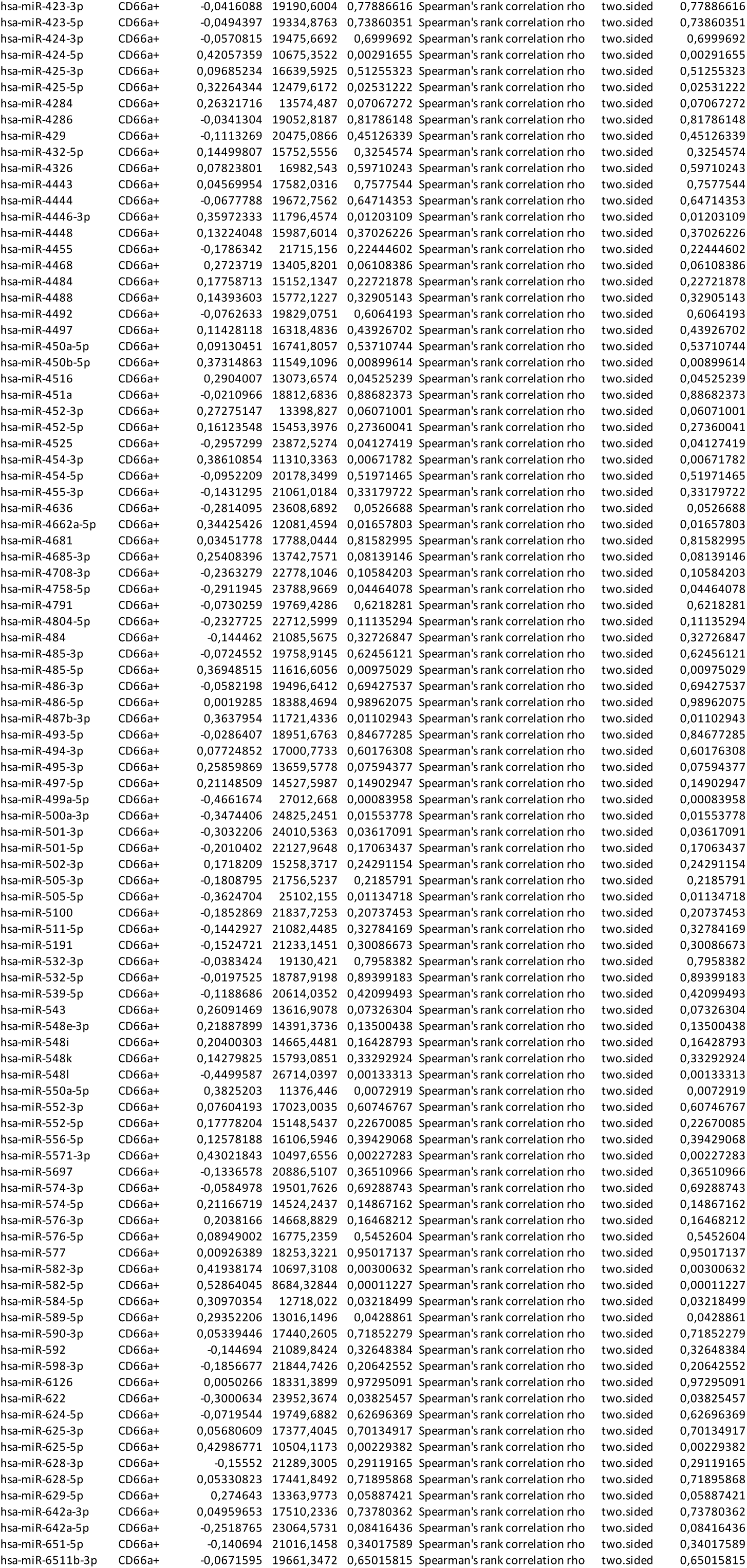
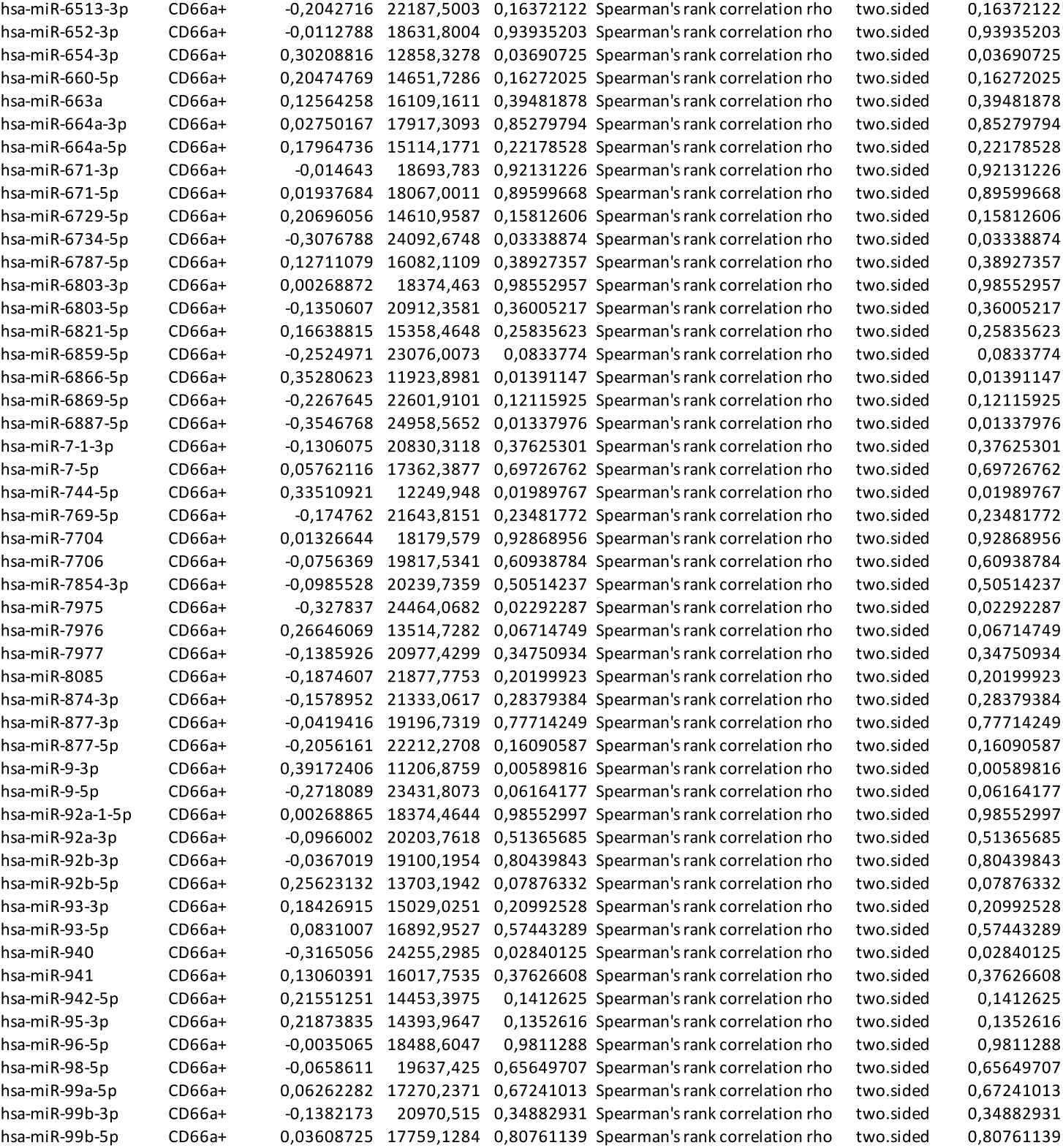
The list of correlations between endoscopic Mayo score and normalized expression of miRNAs expressed in crypt-top (CD66a+) colonic epithelial cell population.

## Notes

### Competing Interest Statement

The authors have declared no competing interest.

### Funding Statement

This work was funded by the Research Council of Lithuania and European Crohn's and Colitis Organisation (grant numbers S-MIP-20-56 and ECCO Grant 2016, respectively).

### Author Declarations

Kaunas Regional Biomedical Research Ethics Committee gave ethical approval for this work (No. BE-2-31, 22-03-2018).

### Summary of Updates

Abstract, introduction, methods and materials, results and discussion updated, including additional WGCNA analysis part.

## References

1. Antoni L., Nuding S., Wehkamp J., Stange EF. Intestinal barrier in inflammatory bowel disease. World J Gastroenterol 2014;20(5):1165–79.

2. Nyström EEL., Martinez-Abad B., Arike L., Birchenough GMH., Nonnecke EB., Castillo PA., et al. An intercrypt subpopulation of goblet cells is essential for colonic mucus barrier function. Science 2021;372(6539).

3. Cichon C., Sabharwal H., Rüter C., Schmidt MA. MicroRNAs regulate tight junction proteins and modulate epithelial/endothelial barrier functions. Tissue Barriers 2014;2(4).

4. Ye D., Guo S., Alsadi R., Ma TY. MicroRNA regulation of intestinal epithelial tight junction permeability. Gastroenterology 2011;141(4):1323–33.

5. Schroeder KW., Tremaine WJ., Ilstrup DM. Coated oral 5-aminosalicylic acid therapy for mildly to moderately active ulcerative colitis. A randomized study. N Engl J Med 1987;317(26):1625–9.

6. Magro F., Gionchetti P., Eliakim R., Ardizzone S., Armuzzi A., Barreiro-de Acosta M., et al. Third European Evidence-based Consensus on Diagnosis and Management of Ulcerative Colitis. Part 1: Definitions, Diagnosis, Extra-intestinal Manifestations, Pregnancy, Cancer Surveillance, Surgery, and Ileo-anal Pouch Disorders. J Crohns Colitis 2017;11(6):649–70.

7. Dalerba P., Kalisky T., Sahoo D., Rajendran PS., Rothenberg ME., Leyrat AA., et al. Single-cell dissection of transcriptional heterogeneity in human colon tumors. Nat Biotechnol 2011;29(12):1120–7.

8. Ewels PA., Peltzer A., Fillinger S., Patel H., Alneberg J., Wilm A., et al. The nf-core framework for community-curated bioinformatics pipelines. Nat Biotechnol 2020:276–8.

9. Pantano L., Estivill X., Martí E. A non-biased framework for the annotation and classification of the non-miRNA small RNA transcriptome. Bioinformatics 2011;27(22):3202–3.

10. Langmead B., Trapnell C., Pop M., Salzberg SL. Ultrafast and memory-efficient alignment of short DNA sequences to the human genome. Genome Biol 2009;10(3):1–10.

11. Kozomara A., Birgaoanu M., Griffiths-Jones S. miRBase: from microRNA sequences to function. Nucleic Acids Res 2019;47(D1):D155–62.

12. Desvignes T., Loher P., Eilbeck K., Ma J., Urgese G., Fromm B., et al. Unification of miRNA and isomiR research: the mirGFF3 format and the mirtop API. Bioinformatics 2020;36(3):698–703.

13. Love MI., Huber W., Anders S. Moderated estimation of fold change and dispersion for RNA-seq data with DESeq2. Genome Biol 2014;15(12):1–21.

14. Ritchie ME., Phipson B., Wu D., Hu Y., Law CW., Shi W., et al. limma powers differential expression analyses for RNA-sequencing and microarray studies. Nucleic Acids Res 2015;43(7):e47.

15. R Foundation for Statistical Computing. R Core Team. R: A language and environment for statistical computing 2021:https://www.r-project.org/.

16. Wickham H. ggplot2: Elegant Graphics for Data Analysis 2016.

17. Jassal B., Matthews L., Viteri G., Gong C., Lorente P., Fabregat A., et al. The reactome pathway knowledgebase. Nucleic Acids Res 2020;48(D1):D498–503.

18. Thomas PD., Ebert D., Muruganujan A., Mushayahama T., Albou LP., Mi H. PANTHER: Making genome-scale phylogenetics accessible to all. Protein Sci 2022;31(1):8–22.

19. Xiao F., Zuo Z., Cai G., Kang S., Gao X., Li T. miRecords: an integrated resource for microRNA-target interactions. Nucleic Acids Res 2009;37(Database issue):D105–10.

20. Hsu S-D., Lin F-M., Wu W-Y., Liang C., Huang W-C., Chan W-L., et al. miRTarBase: a database curates experimentally validated microRNA-target interactions. Nucleic Acids Res 2011;39(Database issue):D163–9.

21. Karagkouni D., Paraskevopoulou MD., Chatzopoulos S., Vlachos IS., Tastsoglou S., Kanellos I., et al. DIANA-TarBase v8: a decade-long collection of experimentally supported miRNA-gene interactions. Nucleic Acids Res 2018;46(D1):D239–45.

22. Ru Y., Kechris KJ., Tabakoff B., Hoffman P., Radcliffe RA., Bowler R., et al. The multiMiR R package and database: integration of microRNA-target interactions along with their disease and drug associations. Nucleic Acids Res 2014;42(17):e133.

23. Yu G., He Q-Y. ReactomePA: an R/Bioconductor package for reactome pathway analysis and visualization. Mol Biosyst 2016;12(2):477–9.

24. Wu T., Hu E., Xu S., Chen M., Guo P., Dai Z., et al. clusterProfiler 4.0: A universal enrichment tool for interpreting omics data. Innov 2021;2(3).

25. Parikh K., Antanaviciute A., Fawkner-Corbett D., Jagielowicz M., Aulicino A., Lagerholm C., et al. Colonic epithelial cell diversity in health and inflammatory bowel disease. Nature 2019;567(7746):49–55.

26. Russo PST., Ferreira GR., Cardozo LE., Bürger MC., Arias-Carrasco R., Maruyama SR., et al. CEMiTool: a Bioconductor package for performing comprehensive modular co-expression analyses. BMC Bioinformatics 2018;19(1):56.

27. Langfelder P., Horvath S. WGCNA: an R package for weighted correlation network analysis. BMC Bioinformatics 2008;9(1):559.

28. Robin X., Turck N., Hainard A., Tiberti N., Lisacek F., Sanchez J-C., et al. pROC: an open-source package for R and S+ to analyze and compare ROC curves. BMC Bioinformatics 2011;12(1):77.

29. Heller F., Florian P., Bojarski C., Richter J., Christ M., Hillenbrand B., et al. Interleukin-13 is the key effector Th2 cytokine in ulcerative colitis that affects epithelial tight junctions, apoptosis, and cell restitution. Gastroenterology 2005;129(2):550–64.

30. Yao D., Dong M., Dai C., Wu S. Inflammation and Inflammatory Cytokine Contribute to the Initiation and Development of Ulcerative Colitis and Its Associated Cancer. Inflamm Bowel Dis 2019;25(10):1595–602.

31. MacDonald TT. Epithelial proliferation in response to gastrointestinal inflammation. Ann N Y Acad Sci 1992;664:202–9.

32. Jacenik D., Zielińska M., Mokrowiecka A., Michlewska S., Małecka-Panas E., Kordek R., et al. G protein-coupled estrogen receptor mediates anti-inflammatory action in Crohn’s disease. Sci Rep 2019;9(1):6749.

33. Ungaro R., Mehandru S., Allen PB., Peyrin-Biroulet L., Colombel J-F. Ulcerative colitis. Lancet (London, England) 2017;389(10080):1756–70.

34. Wu F., Xing T., Gao X., Liu F. miR-501-3p promotes colorectal cancer progression via activation of Wnt/β-catenin signaling. Int J Oncol 2019;55(3):671–83.

35. Akao Y., Nakagawa Y., Naoe T. let-7 microRNA functions as a potential growth suppressor in human colon cancer cells. Biol Pharm Bull 2006;29(5):903–6.

36. Martínez C., Rodiño-Janeiro BK., Lobo B., Stanifer ML., Klaus B., Granzow M., et al. miR-16 and miR-125b are involved in barrier function dysregulation through the modulation of claudin-2 and cingulin expression in the jejunum in IBS with diarrhoea. Gut 2017;66(9):1537–8.

37. Sun T-Y., Li Y-Q., Zhao F-Q., Sun H-M., Gao Y., Wu B., et al. MiR-1-3p and MiR-124-3p Synergistically Damage the Intestinal Barrier in the Ageing Colon. J Crohns Colitis 2022;16(4):656–67.

38. Tian Y., Xu J., Li Y., Zhao R., Du S., Lv C., et al. MicroRNA-31 Reduces Inflammatory Signaling and Promotes Regeneration in Colon Epithelium, and Delivery of Mimics in Microspheres Reduces Colitis in Mice. Gastroenterology 2019;156(8):2281–2296.e6.

39. Masi L., Capobianco I., Magrì C., Marafini I., Petito V., Scaldaferri F. MicroRNAs as Innovative Biomarkers for Inflammatory Bowel Disease and Prediction of Colorectal Cancer. Int J Mol Sci 2022.

40. Su C., Huang D-P., Liu J-W., Liu W-Y., Cao Y-O. miR-27a-3p regulates proliferation and apoptosis of colon cancer cells by potentially targeting BTG1. Oncol Lett 2019;18(3):2825–34.

41. Zhou J., Liu J., Gao Y., Shen L., Li S., Chen S. miRNA-Based Potential Biomarkers and New Molecular Insights in Ulcerative Colitis. Front Pharmacol 2021;12:707776.

42. Béres NJ., Szabó D., Kocsis D., Szűcs D., Kiss Z., Müller KE., et al. Role of Altered Expression of miR-146a, miR-155, and miR-122 in Pediatric Patients with Inflammatory Bowel Disease. Inflamm Bowel Dis 2016;22(2):327–35.

43. Neudecker V., Haneklaus M., Jensen O., Khailova L., Masterson JC., Tye H., et al. Myeloid-derived miR-223 regulates intestinal inflammation via repression of the NLRP3 inflammasome. J Exp Med 2017;214(6):1737–52.

44. Gwiggner M., Martinez-Nunez RT., Whiteoak SR., Bondanese VP., Claridge A., Collins JE., et al. MicroRNA-31 and MicroRNA-155 Are Overexpressed in Ulcerative Colitis and Regulate IL-13 Signaling by Targeting Interleukin 13 Receptor α-1. Genes (Basel) 2018.

45. Xia F., Bo W., Ding J., Yu Y., Wang J. MiR-222-3p Aggravates the Inflammatory Response by Targeting SOCS1 to Activate STAT3 Signaling in Ulcerative Colitis. Turkish J Gastroenterol Off J Turkish Soc Gastroenterol 2022;33(11):934–44.

46. Wang H., Chao K., Ng SC., Bai AH., Yu Q., Yu J., et al. Pro-inflammatory miR-223 mediates the cross-talk between the IL23 pathway and the intestinal barrier in inflammatory bowel disease. Genome Biol 2016;17:58.

47. Allaire JM., Crowley SM., Law HT., Chang S-Y., Ko H-J., Vallance BA. The Intestinal Epithelium: Central Coordinator of Mucosal Immunity. Trends Immunol 2018;39(9):677–96.

48. Chirshev E., Oberg KC., Ioffe YJ., Unternaehrer JJ. Let-7 as biomarker, prognostic indicator, and therapy for precision medicine in cancer. Clin Transl Med 2019;8(1):24.

49. Saatian B., Rezaee F., Desando S., Emo J., Chapman T., Knowlden S., et al. Interleukin-4 and interleukin-13 cause barrier dysfunction in human airway epithelial cells. Tissue Barriers 2013;1(2):e24333.

50. Rosen MJ., Frey MR., Washington MK., Chaturvedi R., Kuhnhein LA., Matta P., et al. STAT6 activation in ulcerative colitis: a new target for prevention of IL-13-induced colon epithelial cell dysfunction. Inflamm Bowel Dis 2011;17(11):2224–34.

51. Kadivar K., Ruchelli ED., Markowitz JE., Defelice ML., Strogatz ML., Kanzaria MM., et al. Intestinal interleukin-13 in pediatric inflammatory bowel disease patients. Inflamm Bowel Dis 2004;10(5):593–8.

52. Giuffrida P., Caprioli F., Facciotti F., Di Sabatino A. The role of interleukin-13 in chronic inflammatory intestinal disorders. Autoimmun Rev 2019;18(5):549–55.

53. Karmele EP., Pasricha TS., Ramalingam TR., Thompson RW., Gieseck RL 3rd., Knilans KJ., et al. Anti-IL-13Rα2 therapy promotes recovery in a murine model of inflammatory bowel disease. Mucosal Immunol 2019;12(5):1174–86.

54. Junttila IS. Tuning the Cytokine Responses: An Update on Interleukin (IL)-4 and IL-13 Receptor Complexes. Front Immunol 2018;9:888.

55. Min M., Yang J., Yang Y-S., Liu Y., Liu L-M., Xu Y. Expression of Transcription Factor FOXO3a is Decreased in Patients with Ulcerative Colitis. Chin Med J (Engl) 2015;128(20):2759–63.

56. Snoeks L., Weber CR., Wasland K., Turner JR., Vainder C., Qi W., et al. Tumor suppressor FOXO3 participates in the regulation of intestinal inflammation. Lab Invest 2009;89(9):1053–62.

57. Doran E., Choy DF., Shikotra A., Butler CA., O’Rourke DM., Johnston JA., et al. Reduced epithelial suppressor of cytokine signalling 1 in severe eosinophilic asthma. Eur Respir J 2016;48(3):715–25.

58. Krndija D., El Marjou F., Guirao B., Richon S., Leroy O., Bellaiche Y., et al. Active cell migration is critical for steady-state epithelial turnover in the gut. Science 2019;365(6454):705–10.

59. Martini E., Krug SM., Siegmund B., Neurath MF., Becker C. Mend Your Fences: The Epithelial Barrier and its Relationship With Mucosal Immunity in Inflammatory Bowel Disease. Cell Mol Gastroenterol Hepatol 2017;4(1):33–46.

60. Godard P., van Eyll J. Pathway analysis from lists of microRNAs: common pitfalls and alternative strategy. Nucleic Acids Res 2015;43(7):3490–7.

